# Immune system and blood-brain barrier-wide biomarker analyses provide causal evidence for autoimmunity in dementia

**DOI:** 10.1101/2022.02.17.22271136

**Authors:** Joni V. Lindbohm, Nina Mars, Pyry N. Sipilä, Archana Singh-Manoux, Heiko Runz, FinnGen, Gill Livingston, Sudha Seshadri, Ramnik Xavier, Aroon D. Hingorani, Samuli Ripatti, Mika Kivimäki

**Author notes:** Correspondence to Dr Joni Lindbohm, Clinicum, Department of Public Health, University of Helsinki, P.O. Box 41, FI-00014 Helsinki, Finland; E-mail address. Authors contributed equally (Samuli Ripatti, Mika Kivimäki).

## Abstract

Immune system and blood brain barrier (BBB) dysfunction are implicated in the development of Alzheimer’s disease and other dementias, but their causal role remains unknown. We performed Mendelian randomization (MR) for over 43,643 immune system and BBB-related biomarkers and identified 126 potential causal risk factors for dementias. A phenome-wide analysis using MR-based polygenic risk score in FinnGen study (N=339,233) for these risk factors revealed a common genetic background for dementias and autoimmune diseases which was supported by further HLA analyses. Pathway analyses linked the 126 proteins to amyloid-β, tau and α-synuclein pathways, increased inflammatory responses, and altered self-tolerance mechanisms. In inverse-probability-weighted analyses simulating randomized controlled drug trials in observational data, anti-inflammatory and immunosuppressive medications were associated with reduced dementia risk (p<0.01 for methotrexate and TNF-α inhibitors). These converging results from different research lines suggest that autoimmunity is a modifiable component in diseases causing dementia.

## Background

Dysregulation of peripheral and central immune systems and chronic neuroinflammation occur in diseases, such as severe infections and autoimmune diseases that relate to increased risk of dementia.^1–3^ In central immune system, neuroinflammation can closely relate to sustained activation of microglia, the resident immune cells in the central nervous system (CNS), and lead to neurodegeneration and increased deposition of plaques and tangles.^1–3^ When CNS encounters infections, trauma, or toxins, microglia are key in the acute immune response and in this phase are neuroprotective. When the microglia activation continues, this leads to chronic production of pro-inflammatory products including reactive oxygen species, nitric oxide, and cytokines.^1–3^ This in turn may exacerbate disease-specific pathology, such as amyloid β and tau protein in Alzheimer’s disease, and neurodegeneration.^1^

In addition, the peripheral immune system and blood brain barrier are thought to be important in the development of dementias.^1, 4^ Because of the crosstalk between the central and peripheral immune system and BBB, the causal factors linking inflammation to dementias may originate from any of these systems. Chronic peripheral inflammation, from low-grade systemic inflammation^5, 6^ and infections,^7^ can disrupt BBB so that it allows neurotoxic and pro-inflammatory plasma components and pathogens to enter CNS.^4^ Dysfunctional BBB may also promote peripherally driven neuroinflammation by increasing endothelial adhesion molecules and chemokines and migration of peripheral leukocytes to CNS.^4^ These processes expose CNS to prolonged neuroinflammation and subsequent neurodegeneration,^1^ although the role of centrally and peripherally driven CNS autoimmunity in dementias remains unclear.^8–11^ In addition, there is limited data to confirm the causal nature of these hypothesised mechanisms.^1–3^

Mendelian randomization (MR), using large GWAS libraries for unconfounded genetic proxies for biomarkers, and triangulation, combining multiple research methods with different sources of strengths and biases to identify convergent evidence, improve validity of causal estimates and success rate in drug development.^12, 13^ Although a range of studies on the immune system and BBB have been conducted,^1–4^ to the best of our knowledge, there are currently no published large-scale, immune system and BBB-wide MR analyses, confirmed by triangulation to identify novel drug targets and risk factors for dementias.

In this study, we analyse biomarkers of immune system and BBB (**Figure 1**) and use MR to study potential causal associations with five outcomes (Alzheimer’s disease, Parkinson’s diseases, frontotemporal dementia, vascular dementia, and poor cognitive performance) and triangulate the results. We identify several novel associations between inflammatory and immune checkpoint-related biomarkers and dementias. Using single nucleotide polymorphisms (SNPs) strongly associated with these biomarkers, we create MR-based polygenic risk scores (PRS) for the diseases. In phenome-wide association analyses (PheWAS) for the PRSs, we observe shared genetic risk factors with autoimmune diseases and dementias, which are supported by HLA analysis. In line with this, our pathway analyses for risk factors identified in MR show enrichment in inflammatory and self-tolerance related processes, which are in close interaction with amyloid β, tau protein, and α-synuclein. Lastly, using inverse-probability-weighted (IPW) survival analysis to simulate a randomized controlled trial design in observational data with up to 20 years of follow-up, we identify anti-inflammatory medications which may lower the risk of dementias. In summary, we detect several novel risk factors and potential drug targets for dementia-causing diseases in a triangulation framework and propose that dementias may have an autoimmune component which is modifiable with anti-inflammatory medication. These findings are set out in **Figure 2**.

**Figure 1.**
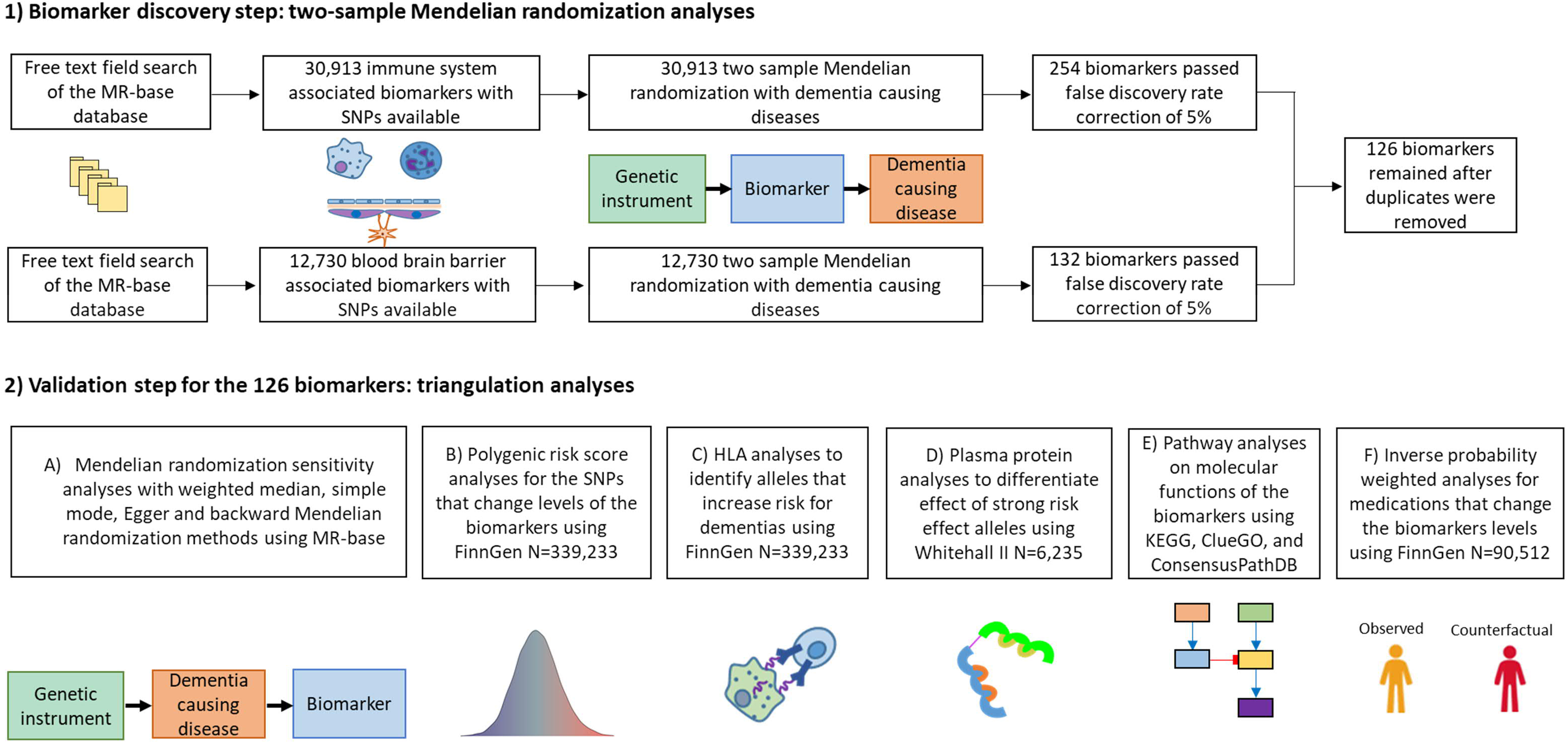
Flowchart of the study design. HLA, human leukocyte antigen; KEGG, Kyoto Encyclopedia of Genes and Genomes; PRS, polygenic risk score; SNP, single nucleotide polymorphism.

**Figure 2.**
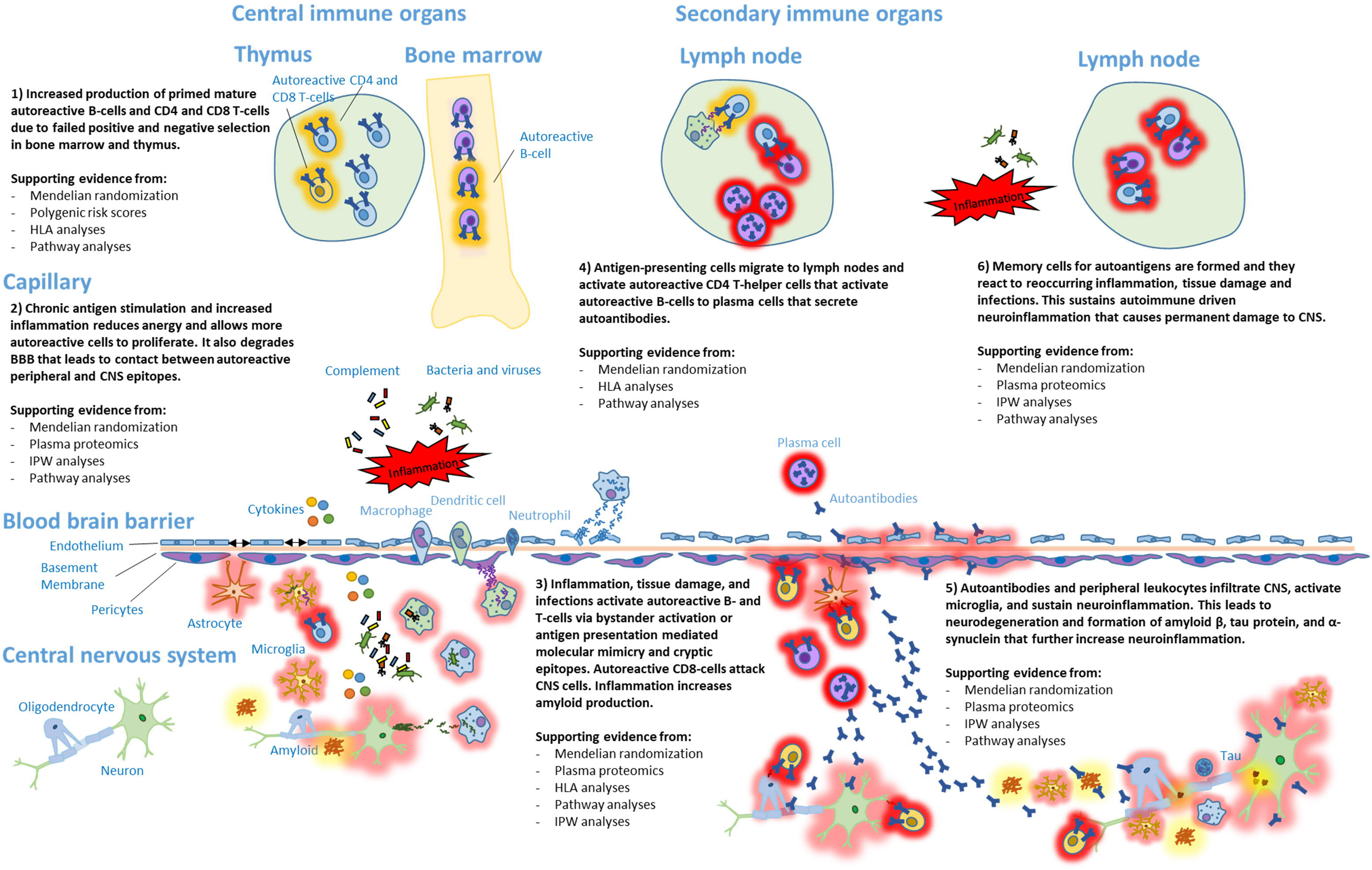
The autoimmune hypothesis of diseases causing dementias, the supporting evidence from current analyses are listed at each mechanism. BBB, blood brain barrier; CD, cluster of differentiation; CNS, central nervoussystem; HLA, Human Leukocyte Antigen; cluster of differentiation; IPW, inverse probability weighted;

## Results

### Mendelian randomization and proteomics analyses

In the discovery step, we used MR to identify potential causal risk factors for dementias. The MR-base search provided 30,913 components for the immune system and 12,730 components for BBB; 254 immune system components and 132 BBB components passed the false discovery rate (FDR) of 5% (**Figure 1**). After removal of duplicates, 126 unique biomarkers were associated (p-value <0.000502) with neurodegenerative diseases in MR analyses using Wald ratio (when only 1 or 2 SNPs was available) or inverse variance-weighted method (when 3 or more SNPs were available) (**Figure 3A and B**). MR sensitivity analyses showed no strong evidence of reverse causality, and evidence of horizontal pleiotropy for only two outcomes. (**STable 1 and SFigures 1A and B**). The 126 biomarkers (FDR <5%) showed few off-target associations, mainly with type 1 diabetes and LDL-C. (**SFigure 2**).

**Figure 3A.**
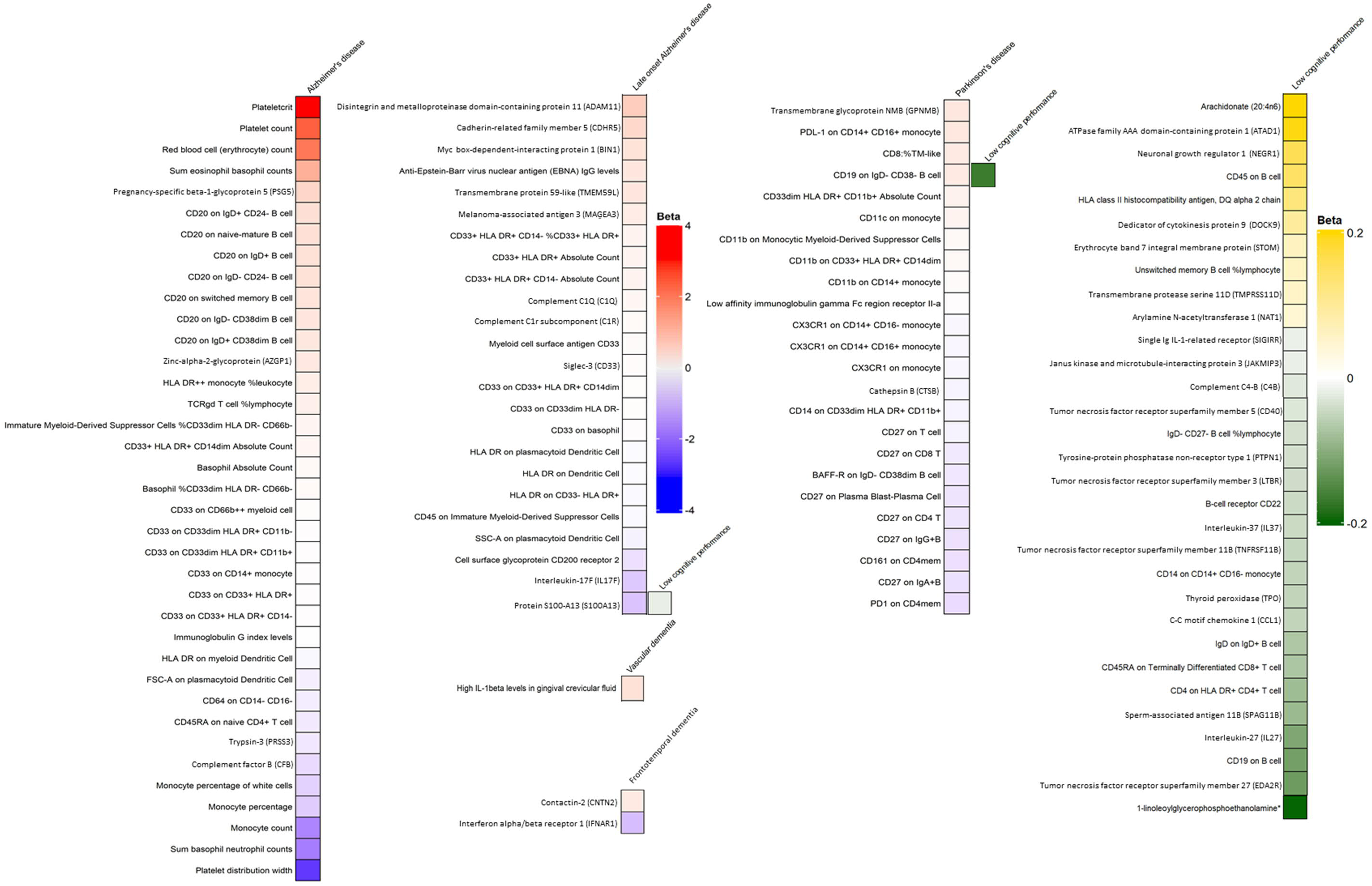
Wald ratio or inverse variance weighted Mendelian randomization estimates for outcome-specific biomarkers.

**Figure 3B.**
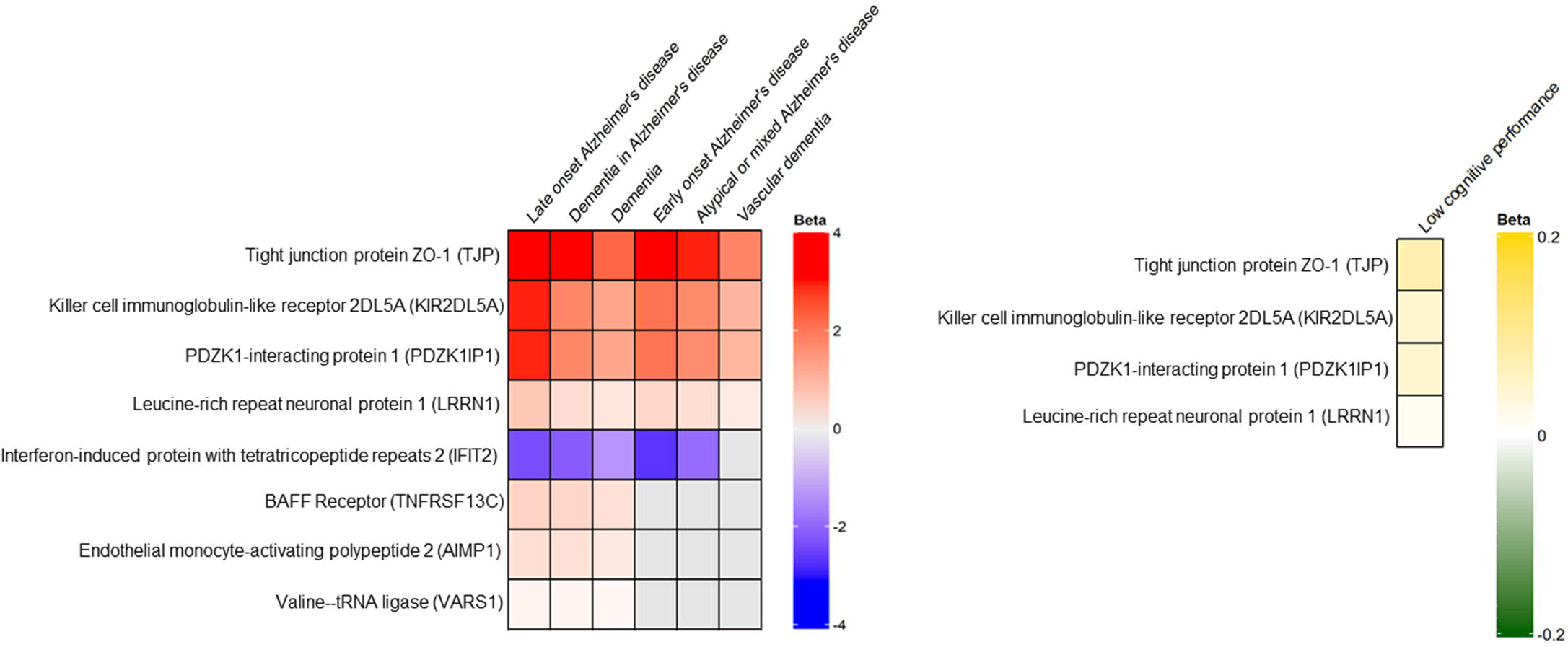
Wald ratio or inverse variance weighted Mendelian randomization estimate for biomarkers that associated with at least three outcomes. These biomarkers localized around *APOE* gene. Biomarkers outcome pairs with no common SNPs available are on grey colour.

While most biomarkers were disease-specific, 8 were associated with Alzheimer’s disease, vascular dementia, and poor cognitive performance. The SNPs for the latter biomarkers were centred within 500 kilobases (kb) from *APOE* gene, one of the strongest genetic risk factors for late-onset Alzheimer’s disease (**SFile 1**). To differentiate the effects of these biomarkers from *APOE*, we studied dementia outcomes and plasma proteins measured within the Whitehall II cohort (N=6,235). Out of the 8 proteins, 2 were associated with all-cause dementia in a 20-year follow-up and 1 (IFIT2) of these remained after adjustment of *APOE* status (**SFigure 3**; data on dementia subtypes were not available).

The 118 outcome-specific biomarkers did not show similar enrichment around high-risk genes for Alzheimer’s disease such as *APP*, *PSEN1*, *PSEN2*, *ADAM10*, *TREM2*, *PLD3*, and *UNC5C*. Instead, they were characterized by inflammatory, chemokine, complement, and adhesion processes (C1Q, C1R, C4B, CCL1, CDHR5, GPNMB, IL1β, IL17, IL27, IL37, LTBR, PTP1B, SIGIRR), antigen presenting and immune checkpoints (HLA-DR, HLA-DQ, BAFFR, C1R, C1Q, CD11, CD19, CD20, CD33, CD40, CX3CR1, PD-1, and PDL-1), and BBB tight junction related biomarkers (TJP, AIMP1, BIN1).

### Polygenic risk score and HLA analysis

To study the diseases that share common genetic background with dementias, we created MR-based polygenetic risk score (PRS) using the SNPs of the 126 biomarkers for dementias and then run a phenome-wide association analysis (PheWAS) (**Figure 4**). After linkage disequilibrium (LD) pruning and SNP matching in FinnGen (N=339,233), 92 SNPs were available for a PRS for Alzheimer’s disease (the number of SNPs for other outcomes was insufficient, ≤25). The PRS was strongly and positively associated with risk of all types of dementia-causing diseases (except Parkinson’s disease) and autoimmune disorders, such as rheumatic diseases, type 1 diabetes and its complications. The PRS was also associated with increased risk of thyroiditis, coronary heart disease, hyperlipidaemia, and the likelihood of having medications for Crohn’s disease (indicating disease severity), but reduced risk of cancers. We studied whether HLA-types that predispose to autoimmune diseases are also associated with dementias. These analyses showed 21 directionally consistent HLA association with dementias and at least one autoimmune disease (**Figure 5A-E**).

**Figure 4.**
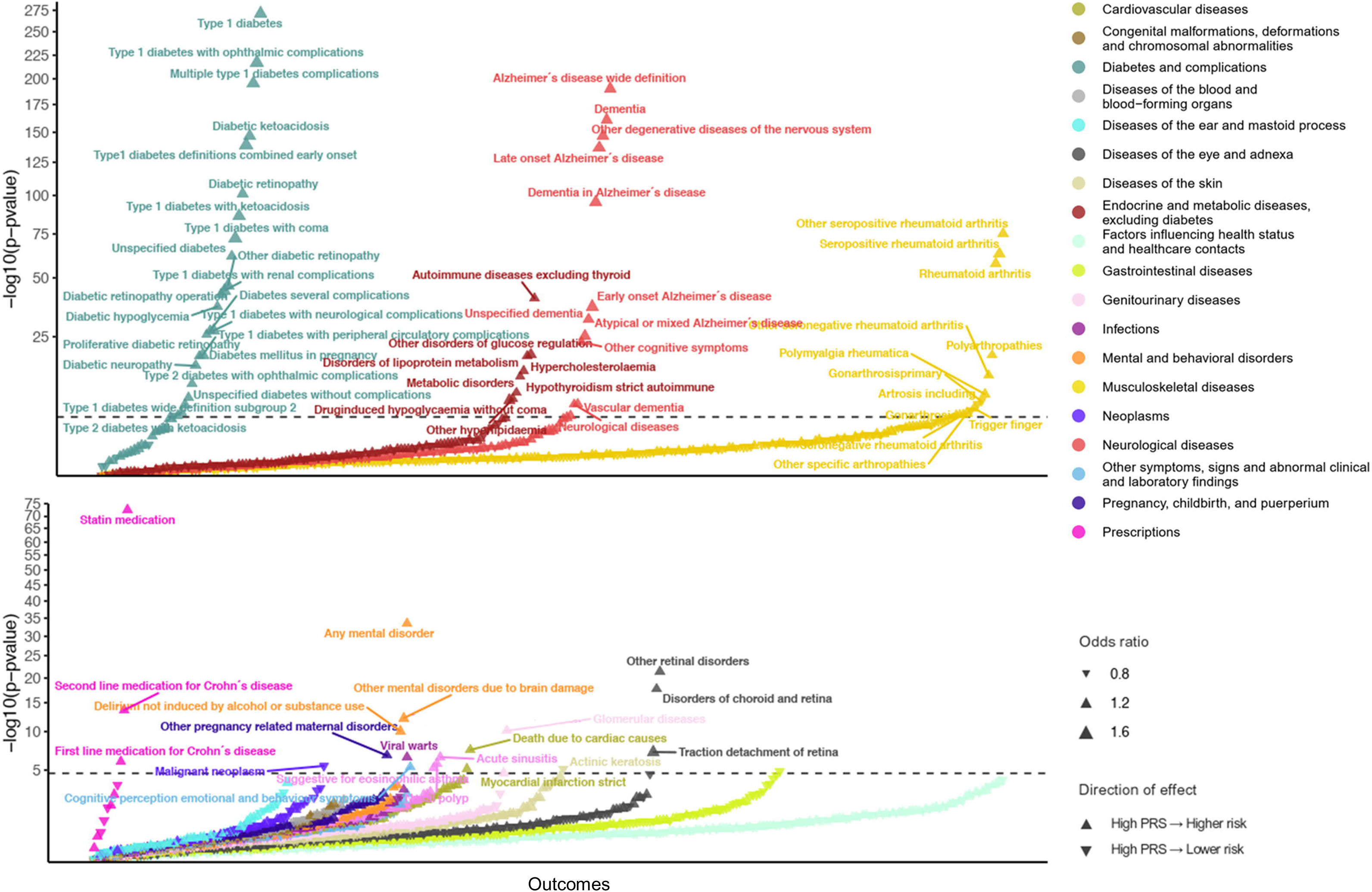
Phenome-wide association analyses for a Mendelian randomization-based polygenic risk score constructed only from SNPs that associated with levels of causal Alzheimer’s disease biomarkers in Mendelian randomization Wald ratio or inverse variance weighted analyses. Upward pointing triangle means risk increasing and downward risk decreasing. Larger triangle indicates larger effect size.

**Figure 5A-E.**
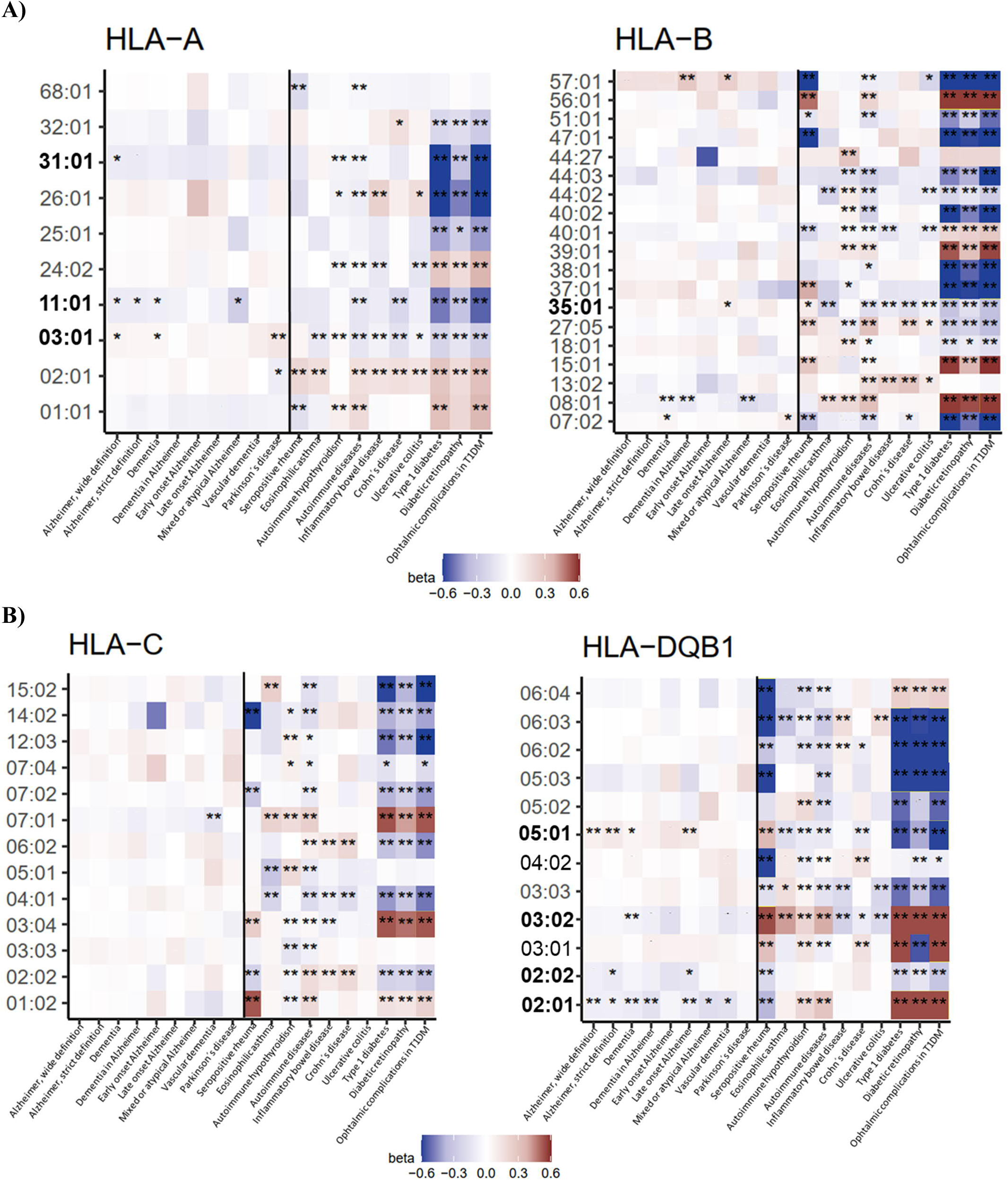
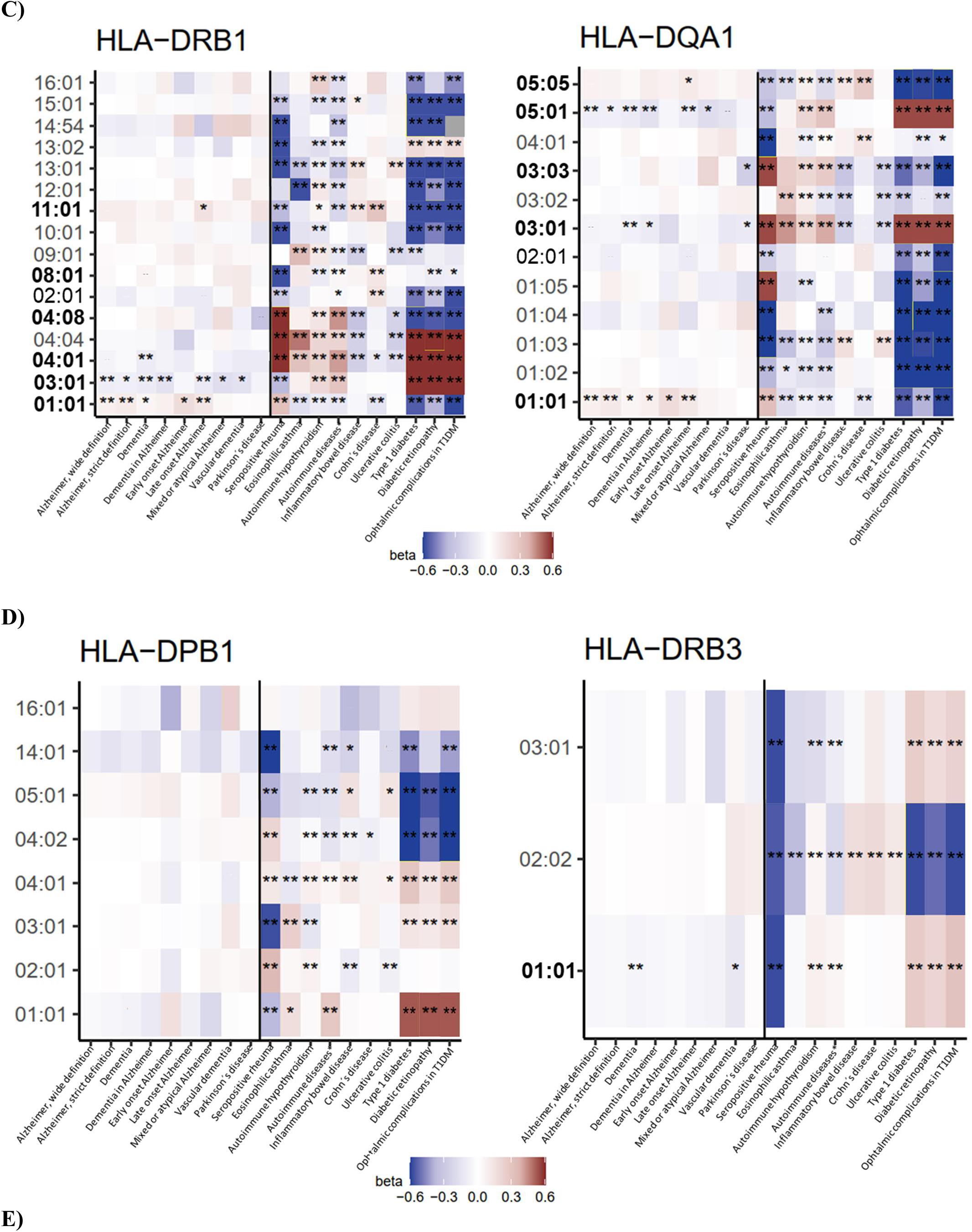
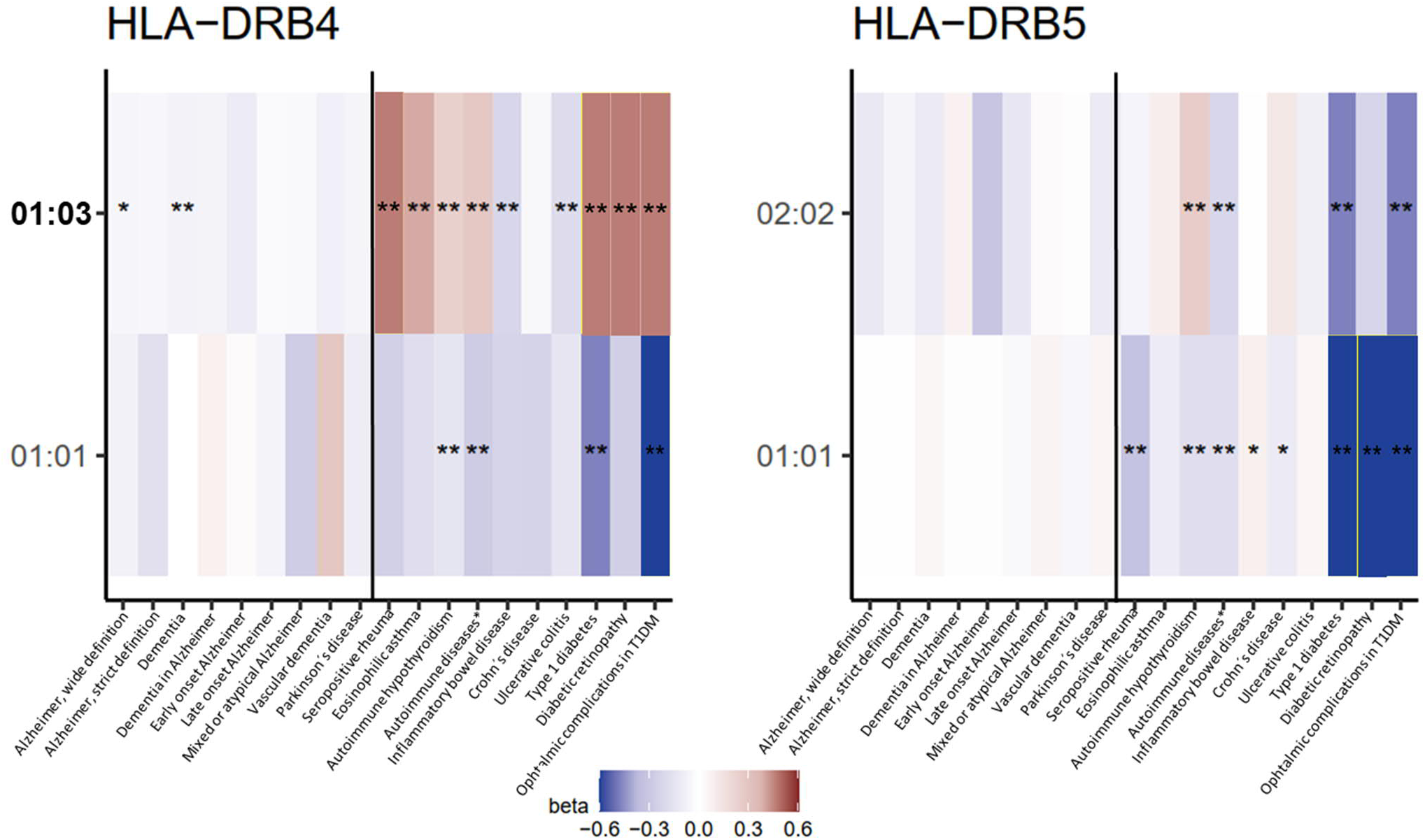
Association between different HLA-allele subclasses (y-axis) and dementia causing diseases, autoimmune diseases and their complications (x-axis). The HLA-types with at least one directionally consistent association with autoimmune diseases are bolded. Left side of the panel describes dementias and right side autoimmune diseases and their complications. *, false discovery rate p < 0.02; **, p < 0.01.

### Pathway analyses

To identify pathways the 126 biomarkers are involved in, we performed analyses based on KEGG, ClueGO, and ConsensusPathDB databases. The findings provided further support for the autoimmune component in dementias. KEGG and ClueGO analyses showed that the identified biomarkers increased MHC-II-mediated antigen presentation in several processes, HLA-DR expression across all hematopoietic cell lines, neuronal adhesion molecule CNTN2, and PD-L1 in T-cell-antigen interactions that reduces self-tolerance. The biomarkers also decreased barrier protecting IL-17F, self-tolerance increasing PDCD1 in T-cell-antigen interactions, and antiviral complement factor B and IFNAR1 (**SFigures 4A-H and 5**). ConsensusPathDB analyses showed that all of the 42 plasma proteins included in the 126 biomarkers had physical interactions with or were in a biological pathway within zero to two molecule distance from amyloid precursor protein, tau protein, or α-synuclein (**SFigure 6A-R**).

### Inverse probability-weighted analyses

To evaluate the autoimmune hypothesis in relation to modifiability and drug repurposing, we used inverse probability-weighted (IPW) analyses in FinnGen and studied whether commonly used anti-inflammatory and immunosuppressive medications are likely to reduce the risk of dementias (**Table 1**). We validated the IPW protocol with a positive control, by replicating the randomized controlled trial (RCT) effect between statin medication and myocardial infarction and used anti-inflammatory medications as negative control.^14–17^ Supporting the autoimmune hypothesis, risk of dementia-causing diseases was reduced with methotrexate and TNF-α inhibitors. Methotrexate appeared to decrease risk only in those with above-average MR-based autoimmune PRS (50% cut-off). To identify rare medications not detectable in FinnGen with sufficient power, we additionally searched the OpenTargets database. This search identified 64 drugs that target 18 of the 126 proteins, suggesting that these may have potential for repurposing to treatment of dementias (**SFile 2**).

**Table 1.**
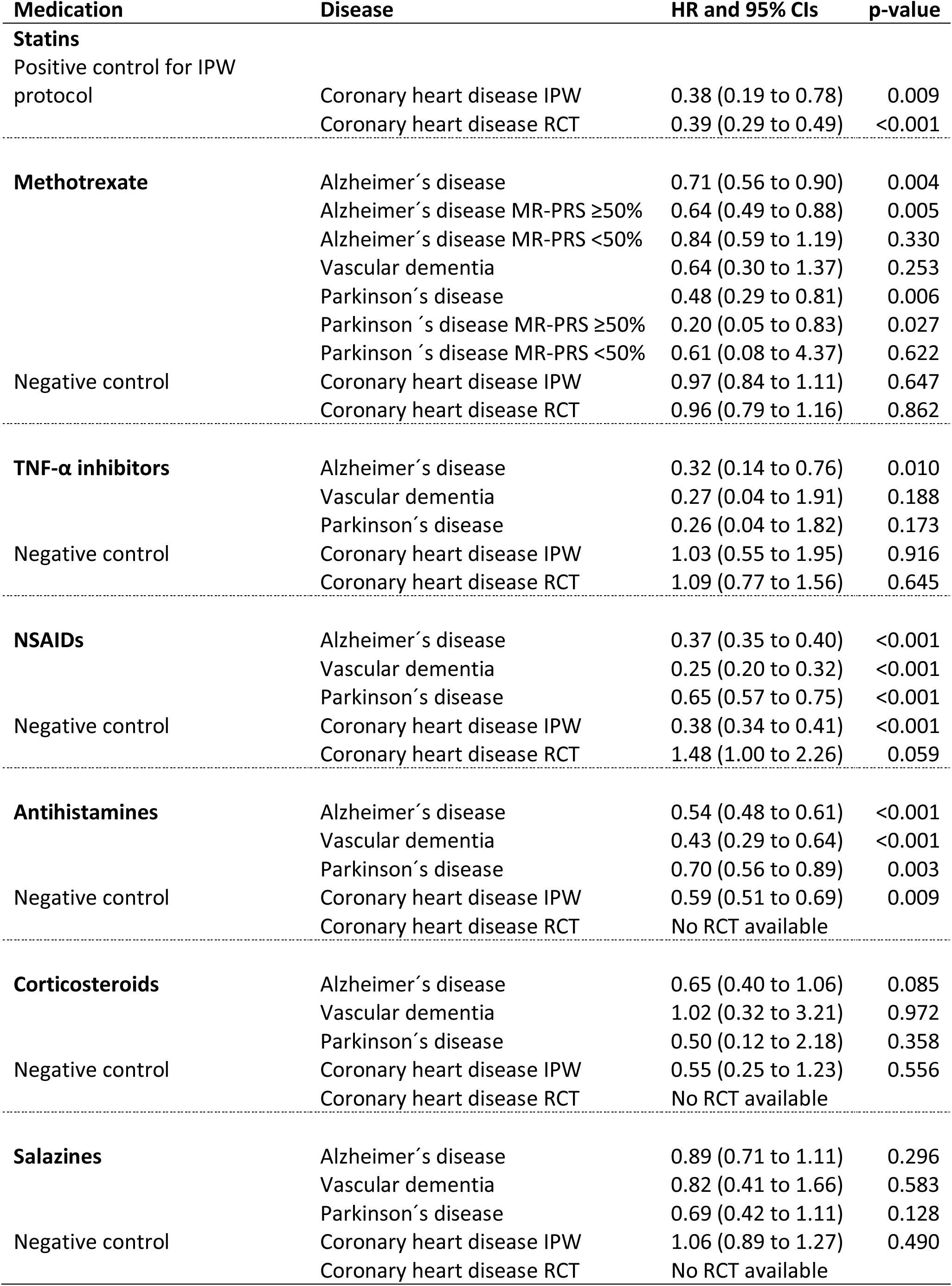
Associations between dementias and anti-inflammatory medications from inverse probability weighted (IPW) analyses in the FinnGen study. The positive control IPW analyses validate the analysis protocol by replicating association between statin medication and coronary heart disease. The negative control IPW analyses validate the associations between each anti-inflammatory medication and dementias by replicating the null finding between anti-inflammatory medications and coronary heart disease. The baseline variables in IPW analyses were birth year, sex, ten principal components and time-varying variables statin, ACE-blocker, AT-blocker, renin-blocker, calcium channel blocker, any diuretic, insulin, metformin, other diabetes drug, depression medication, antipsychotic, and anticoagulant use as well as time varying any cancer, myocardial infarction, atrial fibrillation, heart failure, venous thromboembolism, ischemic stroke, intracerebral haemorrhage, subarachnoid haemorrhage, obesity, sleep apnoea, and chronic obstructive pulmonary diseases, with informative censoring included. Because of limited number of participants, TNF-α inhibitors users in dementias include also baseline users, other analyses include only medication-free individuals at baseline. The estimates for randomized control trials are from references 14 to 17. Secondary prevention randomized control trial was used for methotrexate because no primary prevention trial was available. MR-PRS, Mendelian randomization based polygenic risk score for Alzheimer’s disease; NSAID, non-steroidal anti-inflammatory drugs; TNF-α, tumor necrosis factor α; IPW, inverse probability weighted analyses; RCT, randomized control trials.

## Discussion

Using triangulation across Mendelian randomization, plasma proteomics, polygenic risk scores, HLA type, pathway, and IPW analyses, we identified causal support for 126 risk factors and potential drug targets for dementia-causing diseases. Of these 118 were novel MR associations. Most of the identified risk factors and targets were outcome specific, and included inflammatory, self-tolerance, and/or BBB tight junction related biomarkers. A phenome-wide analysis of PRS from SNPs associated with the identified risk factors showed that Alzheimer’s disease share genetic background with autoimmune diseases, such as rheumatoid arthritis and type 1 diabetes. The autoimmune hypothesis was also supported by HLA analyses showing 21 HLA-type associations common to dementias and autoimmune diseases. Pathway analyses linked the 126 biomarkers to autoimmunity via antigen presentation and reduced self-tolerance and showed that all 42 plasma proteins of the 126 biomarkers are closely related to α-synuclein, amyloid precursor, and tau protein pathways that characterize Alzheimer’s and Parkinson’s diseases and dementia with Levy bodies. The autoimmune hypothesis was further supported by IPW analyses mimicking randomized controlled drug trial in observational data. They suggested anti-inflammatory medication use may potentially reduce the risk of these illnesses.

Several lines of earlier and current evidence support the hypothesis that diseases causing dementia have an autoimmune component (**Figure 2**). Epidemiological studies have shown higher rates of cognitive impairment, Alzheimer’s diseases, Parkinson’s disease, vascular, and frontotemporal dementia in rheumatic disease patients.^18–22^ Diabetes is a recognized risk factor for Alzheimer’s disease^23^ and individuals with type 1 diabetes have higher rate of cognitive decline, Alzheimer’s disease, and vascular dementia.^24, 25^ Thyroiditis, which often has an autoimmune origin, increases risk of dementias.^10, 26^ In agreement with these findings, we showed that MR-based Alzheimer’s disease PRS increased risk of all these autoimmune diseases. In addition, the PRS was associated with reduced risk of cancers, which is a well-described beneficial side effect of reduced self-tolerance commonly harnessed in cancers medications.^27^

Furthermore, risk factors, course of disease, and clinical picture in dementias and autoimmune diseases have several similarities. Alzheimer’s and autoimmune diseases are more common in women^9, 28, 29^ and the diseases share inflammatory risk factors such as smoking and obesity.^9, 23, 30, 31^ Infections increase risk for autoimmune diseases,^32^ rate of cognitive decline,^1^ and dementias even 20 years before the diagnosis,^7^ and can acutely trigger rheumatoid arthritis, type 1 diabetes and the formation of autoantibodies against CNS that lead to tissue degeneration.^8, 9, 11^ Interestingly, many inflammation-related biomarkers in our analyses influenced pathways closely related to amyloid precursor protein, tau protein, or α-synuclein suggesting that they may contribute to proteinopathies that characterize of Alzheimer’s and Parkinson’s disease and dementia with Levy bodies. This process has some similarities with AA form of amyloidosis in rheumatic arthritis in which uncontrolled inflammation accumulates amyloid to a range of tissues^33^ and with islet amyloidosis in diabetes that leads to tissue degeneration.^34^ Two MR studies suggested inflammatory risk factor may not be causally linked to dementias.^35, 36^ However, those studies did not focus on processes that control self-tolerance and they may have used inflammatory markers that are only responses to these processes and thus prone to produce null finding or reverse causation in MR analyses.

### Common cellular mechanisms between autoimmune disease and dementias

At cellular level, loss of central tolerance in central immune organs and subsequent escape of autoreactive B- and T-cells to periphery^37, 38^ are thought to predispose to autoimmune diseases together with certain HLA alleles.^9, 39, 40^ Conversion to disease may occur after complex interplay between inflammation, infections, and tissue damage, with subsequent failure of peripheral self-tolerance mechanisms such as ignorance, anergy, suppression, inhibition, and antigen presentation.^37, 38^ Our results link all of these mechanism to the development of dementias. BBB normally controls tightly the access of toxins, pathogens, and peripheral molecules and cells to CNS.^4^ This keeps peripheral lymphocytes “ignorant” of CNS structures and prevents activation of autoreactive cells against CNS.^38^ However, chronic antigen exposure and activation of innate and adaptive immune systems impair BBB by releasing pro-inflammatory cytokines, prostaglandins, and histamine leading to endothelial damage and upregulation of adhesion molecules.^4, 41^ This exposes CNS to autoreactive peripheral lymphocytes that lose their ignorance.^38^ The prolonged inflammation also leads to reduced anergy, which allows more potentially self-reactive T-lymphocytes to proliferate.^38^

Our MR analyses support these processes. We showed that higher plasma levels of the integral part of BBB, the tight junction component (TJP1^42^), and proteins degrading it (AIMP1^43^ and BIN1^44^), increased the risk of dementias. Moreover, pro-inflammatory C1Q, C1R, CD20, CD11, CDHR5, GPNMB, and IL1β^45–49^ elevated risk of dementias, whereas anti-inflammatory C4B, IL27, IL37, PTP1B, and SIGGIRR^50–54^ and barrier protecting IL-17F^55^ reduced the risk. In suppression and inhibition, regulatory T-cells suppress, or molecules on B- and T-cells inhibit activation of lymphocytes at peripheral checkpoints.^38^ Our MR analyses identified several checkpoint regulators including BAFFR, C1R, C1Q, CD11, CD19, CD20, CD22, CD33, CD40, CX3CR1, LTBR, PD-1, and PDL-1^37, 48, 56–64^ as potential causal risk factors for poor cognitive performance and dementias, uncovering the importance of checkpoint control and autoreactive lymphocytes in dementias.

HLA analyses were also consistent with the autoimmunity hypothesis. After the increase in BBB permeability, peripherally driven neuroinflammation activates central neuroinflammatory responses in astrocytes and microglia.^1, 4^ The chronic neuroinflammation, pathogens, and tissue damage initiate antigen presentation in HLA receptors on antigen-presenting cells in CNS that can activate the autoreactive B- and T-cells and initiate self-sustaining autoimmunity^11, 40^ against CNS structures. Supporting this we identified 21 shared HLA-types between autoimmune diseases and dementias. Similarly, MR and pathway analyses supported causal status of HLA-DR and HLA-DQ cell types in dementias.

The autoreactive cells can be activated via bystander activation, or HLA-mediated molecular mimicry and epitope spreading.^8–10, 38^ In bystander activation, sustained or excessive immune response can bypass normal T-cell antigen presentation to B-cells and activate autoreactive B-cell to plasma cell that produces autoantibodies. In molecular mimicry, autoreactive T- and B-cells first target foreign antigens, and then cross-react with similar self-antigens expressed by host cells. In epitope spreading, cryptic epitopes (parts of dying cells, and intrinsic or foreign DNA or RNA) normally invisible to immune system become visible after tissue damage, which activates autoreactive B- and T cells on these epitopes.

Our results also provide support for these mechanisms. In MR analyses, we observed that higher levels of naïve and memory CD20 B-cells, the main drug target for B-cell induced autoimmune diseases,^65^ increases risk for Alzheimer’s diseases. Similarly, higher levels of CD19, a co-receptor on B-cells that can lead to autoimmunity when hyperactivated,^66^ increased Parkinson’s diseases risk. We also observed that higher levels of IgG against Epstein-Barr virus’ nuclear antigen, a marker of active infection that can mimic CNS structures,^67, 68^ elevated Alzheimer’s diseases risk. In addition, IFIT2 that intercepts foreign RNA and protects against central viral infections,^69^ reduced the risk of all dementia causing diseases except Parkinson’s diseases. Moreover, the pathway analyses suggested that many proteins (COPS6, SOCS3, PSMB5, TOLLIP, TRIM21, PRPF40A, NOC4L, RPS6, IK, KRR1, and MTO1)^70–80^ involved in intracellular pathways from DNA, RNA, and protein processing to antigen presentation may be linked with amyloid β, tau protein, and α- synuclein.

The identified 8 proteins with SNPs located near *APOE* gene were associated with increased BBB permeability, inflammation, neurotoxicity, and lipid transport disturbances,^42, 56, 69, 81–84^ which are all processes linked to *APOE* in Alzheimer’s disease.^85^ Our results suggest that the 8 proteins may be linked to the effects of *APOE*. Another novel result was that higher levels of CD33, one of the top GWAS hits for Alzheimer’s diseases^86^ and target receptor on microglia for an ongoing trial (NCT03822208) including mild to moderate Alzheimer’s diseases patients, increased the risk of disease even when expressed on peripheral leukocytes.

### Anti-inflammatory medications and dementias

Our IPW analyses on use of anti-inflammatory medications, including records of filled drug prescriptions in initially dementia-free individuals at baseline followed up to 20 years supported the modifiability of dysfunctional autoimmune processes. We replicated protective associations between TNF-α inhibitors,^87^ methotrexate.^88^ and Alzheimer’s disease. As novel findings we showed that methotrexate may be protective only for Alzheimer’s and Parkinson’s disease among those with high MR-based PRS. These findings are plausible on a cellular level for BBB degradation and neuroinflammation. TNF-α is a gatekeeper of inflammatory responses and endothelial permeability.^89^ Methotrexate, in turn, modulates a range of inflammatory and immune responses in nearly every cell types forming the foundations for treatment of rheumatoid arthritis.^9^ NSAIDs and antihistamines appeared to decrease risk for all dementias but the validation using a negative control outcome (coronary heart disease) suggested that the IPW model for these medications was insufficient to replicate trial design.

Randomized controlled trials of TNF-α inhibitors in Alzheimer’s disease patients have not shown benefits in 1 to 2 year follow-up.^90, 91^ However, in autoimmune diseases, early initiation of anti-inflammatory medication is of paramount importance suggesting that the trials of medication for people with established dementia may be too late.^9^

### Strengths and limitations

Our triangulation approach has several strengths. By using a broad set of immune system and BBB biomarkers we were able to study these systems comprehensively without restrictions to specific predefined hypotheses. Our findings are based on MR, a promising approach to examine causality,^12^ and on analyses using plasma proteomics, PRS, HLA, pathway, and IPW analyses in two independent cohorts and a range of databases. Plasma protein analyses allowed us to adjust effect estimates for *APOE* which was not possible in Mendelian randomization analyses for some proteins. PRS and HLA analyses used data from the FinnGen study with a sample size of 340,000 providing sufficient statistical power for the identification of shared novel risk alleles between dementias and autoimmune disease. We used ConsensusPathDB, which is one of the most comprehensive collection of databases on molecular pathways and interactions.^92^ The medication analyses used IPW survival analysis that is suggested to provide more reliable causal estimates than traditional survival analyses of observational data.^93^

Our study has also several limitations. Although we explored over 40,000 biomarkers related to the immune system and BBB, we may have missed some biomarkers due to limitations in search protocol or lack of SNPs. Our plasma protein analyses were limited by lack of data on dementia subtypes. The PRS and PheWAS analyses were done with samples with European ancestry and may not apply to non-European ancestries. Due to limited number of SNPs included in PRS, we may have missed important immune system and BBB related associations in our phenome-wide analyses. Nonetheless, the Alzheimer’s MR-based PRS associated strongly with all types of dementia (except Parkinson’s disease) and a range of outcomes related to type 1 diabetes and rheumatic diseases suggesting that at least these findings are robust. The IPW analyses on medications may include some bias due to a limited number of covariates for simulation of randomized control trial. However, major bias is unlikely for the main findings since the analysis protocol was validated by a positive- and negative-control analyses.

## Conclusions

In summary, this study provides novel insights into autoimmunity, BBB, and inflammatory dysfunctions that may contribute to the development of dementias. As the inflammatory components are potentially modifiable with anti-inflammatory medications, the present results also raise hypotheses on several novel drug targets for dementia-causing diseases which need to be validated in future experimental studies. Randomized controlled trials on early initiation of these medications for high-risk individuals are warranted while antigen-specific prevention strategies may offer new avenues in search of treatment for dementias.

## Methods

### Mendelian randomization

The SNPs for biomarkers and outcomes were searched from MR-base database.^94^ Immune system and BBB search terms were identified using identifiers of cell types, receptors, proteins, metabolites, and genes. The identifiers were searched from several publications^3, 4, 6, 23, 36, 38, 40, 95, 96^ and from Uniprot^97^ with search terms “immune” and “blood brain barrier”. Outcomes included all cohorts with following diseases: all types of Alzheimer’s disease, Parkinson’s disease, vascular dementia, frontotemporal dementia, dementia in general, and progression of dementia. (**SMethods**). Poor cognitive performance was chosen as an intermediate outcome. Two sample MR was used to analyse associations between biomarkers and outcomes.^12^ First analyses estimated effects with Wald ratio or inverse variance-weighted analyses.^94^ For biomarker-outcome pairs that passed FDR of 5% and shared over two common SNPs we performed sensitivity analyses with weighted median, simple mode, MR Egger, and backward Mendelian randomization^12^ with TwoSampleMR and MRInstruments R packages.

To assess off-target effects of the observed causal biomarkers, we performed a phenome-wide MR analysis for each biomarker, using GWAS summary statistics for 210 UK Biobank endpoints conducted by Neale lab and with recognized dementia risk factors.^23, 98^

### Plasma protein analyses

Plasma protein measurements in Whitehall II study were available for 6,235 individuals of whom 310 developed dementia and the methods have been describe before (**SMethods**).^6, 99, 100^ In brief, the plasma proteins were measured using the SomaScan version 4.0 and 4.1 assays.^6, 101, 102^ The assays were validated against external reference population and protein-specific conversion coefficients were used to balance the technical differences between versions 4.0 and 4.1. Using DNA extracted from whole blood, a standard PCR assay determined *APOE* genotype using the salting out method.^103, 104^ In analyses, proteins were transformed to a normal distribution by inverse rank-based normal transformation. The analyses used plasma samples measured in 1995/1997 and the follow-up ended at onset of dementia, death, or 1^st^ of October 2019, which ever occurred first. Cox regression models adjusted for age, sex, and *APOE* estimated associations between the proteins and dementia.^105^ The proportionality assumption in all Cox models was assessed with Schoenfeld residuals and with log-log plots.^105^ We used statistical software R (3.6.0 and 4.1.0) for these analyses.

### Polygenic risk score and IPW analyses

FinnGen Data Freeze 8 contains 339,233 individuals, and is a collection of prospective epidemiological and disease-based cohorts, and hospital biobank samples and links genotypes by the unique national personal identification numbers to nationwide health registries, including the national hospital discharge (available from 1968), death (1969–), cancer (1953–) and medication reimbursement (1964–) and purchase (1995–) registries. Genotyping and imputation is described in **SMethods**.

PRS was constructed from SNPs that were associated with the 126 MR identified causal biomarkers for dementias. The SNPs were LD-pruned with clumping cut-off R^2^ = 0.01 in 500kb window with R package TwoSampleMR. The available SNPs were searched from FinnGen genotypes and the final scores were generated with PLINK v2.00aLM3 by calculating the SNP-biomarker beta weighted sum of risk alleles for each SNP. The PRS was scaled to zero mean and unit variance, and we analysed its phenome-wide associations across 2401 disease endpoints. The association between PRS and endpoints were studied with logistic regression, adjusting for birth year, sex, and ten first principal components of ancestry.

The IPW analyses^93^ were done in FinnGen and included participants aged over 45 without any dementia-causing disease at baseline (N = 90,512). ATC codes for anti-inflammatory medication use were obtained from linkage to national medication purchase registry (data starts from 1995) and in each analyses we included only new medication users to simulate trial design.^106^ A model including a range of baseline and time-varying variables and informative censoring estimated the weights (**SMethods**). The analyses used time-varying Cox models and assumptions were assessed similar to plasma proteins. The follow-up started in 1997 and ended at onset of dementia, death, or 31^st^ of December 2019, which ever occurred first. We used R (4.1.2) and ipw package for these analyses.

### HLA analyses

The analyses were done using HLA alleles, imputed with high accuracy using a Finnish-specific reference panel, which has been previously described in detail.^107^ After filtering based on a HLA carrier frequency of ≥0.01 and posterior probability of ≥0.6, we assessed the association between HLA alleles and dementias and autoimmune diseases using logistic regression adjusted for birth year, sex, and ten first principal components of ancestry.

### Pathway analyses

KEGG pathway analysis was done with Generally Applicable Gene-set Enrichment (GAGE),^108^ using MR betas and p-values as input for expression ratios. GO term enrichment analyses were done with ClueGO version 2.5.8^109^ in Cytoscape version 3.7.2^110^ (**SMethods**). The hypergeometric test used all the 76 plasma proteins or surface receptors of the 126 biomarkers as input and all immune system and BBB related proteins from Uniprot^97^ as background, with correction for 5% FDR. The shortest interaction path analyses were done with ConsensusPathDB,^92^ a web based analysis tool containing several biomedical databases.

### Open Targets analyses

Medications that change the levels of the 126 biomarkers were searched from Open Targets database using Uniprot protein and Entrez gene symbols.

### Ethics statement

In the Whitehall II study, research ethics approvals were renewed at each wave; the most recent approval was obtained from the University College London Hospital Committee on the Ethics of Human Research (reference number 85/0938). Written, informed consent from participants was obtained at each contact.

Patients and control subjects in FinnGen provided informed consent for biobank research, based on the Finnish Biobank Act. Alternatively, separate research cohorts, collected prior the Finnish Biobank Act came into effect (in September 2013) and start of FinnGen (August 2017), were collected based on study-specific consents and later transferred to the Finnish biobanks after approval by Fimea (Finnish Medicines Agency), the National Supervisory Authority for Welfare and Health. Recruitment protocols followed the biobank protocols approved by Fimea. The Coordinating Ethics Committee of the Hospital District of Helsinki and Uusimaa (HUS) statement number for the FinnGen study is Nr HUS/990/2017.

The FinnGen study is approved by Finnish Institute for Health and Welfare (permit numbers: THL/2031/6.02.00/2017, THL/1101/5.05.00/2017, THL/341/6.02.00/2018, THL/2222/6.02.00/2018, THL/283/6.02.00/2019, THL/1721/5.05.00/2019 and THL/1524/5.05.00/2020), Digital and population data service agency (permit numbers: VRK43431/2017-3, VRK/6909/2018-3, VRK/4415/2019-3), the Social Insurance Institution (permit numbers: KELA 58/522/2017, KELA 131/522/2018, KELA 70/522/2019, KELA 98/522/2019, KELA 134/522/2019, KELA 138/522/2019, KELA 2/522/2020, KELA 16/522/2020), Findata permit numbers THL/2364/14.02/2020, THL/4055/14.06.00/2020,,THL/3433/14.06.00/2020, THL/4432/14.06/2020, THL/5189/14.06/2020, THL/5894/14.06.00/2020, THL/6619/14.06.00/2020, THL/209/14.06.00/2021, THL/688/14.06.00/2021, THL/1284/14.06.00/2021, THL/1965/14.06.00/2021, THL/5546/14.02.00/2020, THL/2658/14.06.00/2021, THL/4235/14.06.00/2021 and Statistics Finland (permit numbers: TK-53-1041-17 and TK/143/07.03.00/2020 (earlier TK-53-90-20) TK/1735/07.03.00/2021).

The Biobank Access Decisions for FinnGen samples and data utilized in FinnGen Data Freeze 8 include: THL Biobank BB2017_55, BB2017_111, BB2018_19, BB_2018_34, BB_2018_67, BB2018_71, BB2019_7, BB2019_8, BB2019_26, BB2020_1, Finnish Red Cross Blood Service Biobank 7.12.2017, Helsinki Biobank HUS/359/2017, Auria Biobank AB17-5154 and amendment #1 (August 17 2020), AB20-5926 and amendment #1 (April 23 2020), Biobank Borealis of Northern Finland_2017_1013, Biobank of Eastern Finland 1186/2018 and amendment 22 § /2020, Finnish Clinical Biobank Tampere MH0004 and amendments (21.02.2020 & 06.10.2020), Central Finland Biobank 1-2017, and Terveystalo Biobank STB 2018001.

## Data Availability

Data, protocols, and other metadata of the Whitehall II and FinnGen studies are available according to the data sharing policies of these studies. Pre-existing data access policy for Whitehall II specify that research data requests can be submitted to each steering committee; these will be promptly reviewed for confidentiality or intellectual property restrictions and will not unreasonably be refused. Detailed information on data sharing can be found here: https://www.ucl.ac.uk/epidemiology-health-care/research/epidemiology-and-public-health/research/whitehall-ii/data-sharing Individual-level patient or protein data may further be restricted by consent, confidentiality, or privacy laws/considerations. The FinnGen data may be accessed through Finnish Biobanks FinBB portal (www.finbb.fi).

https://www.ucl.ac.uk/epidemiology-health-care/research/epidemiology-and-public-health/research/whitehall-ii

## Acknowledgments and Funding Sources

We want to acknowledge the participants and investigators of FinnGen study. The FinnGen project is funded by two grants from Business Finland (HUS 4685/31/2016 and UH 4386/31/2016) and the following industry partners: AbbVie Inc., AstraZeneca UK Ltd, Biogen MA Inc., Bristol Myers Squibb (and Celgene Corporation & Celgene International II Sàrl), Genentech Inc., Merck Sharp & Dohme Corp, Pfizer Inc., GlaxoSmithKline Intellectual Property Development Ltd., Sanofi US Services Inc., Maze Therapeutics Inc., Janssen Biotech Inc, Novartis AG, and Boehringer Ingelheim. Following biobanks are acknowledged for delivering biobank samples to FinnGen: Auria Biobank (www.auria.fi/biopankki), THL Biobank (www.thl.fi/biobank), Helsinki Biobank (www.helsinginbiopankki.fi), Biobank Borealis of Northern Finland (https://www.ppshp.fi/Tutkimus-ja-opetus/Biopankki/Pages/Biobank-Borealis-briefly-in-English.aspx), Finnish Clinical Biobank Tampere (www.tays.fi/en-US/Research_and_development/Finnish_Clinical_Biobank_Tampere), Biobank of Eastern Finland (www.ita-suomenbiopankki.fi/en), Central Finland Biobank (www.ksshp.fi/fi-FI/Potilaalle/Biopankki), Finnish Red Cross Blood Service Biobank (www.veripalvelu.fi/verenluovutus/biopankkitoiminta) and Terveystalo Biobank (www.terveystalo.com/fi/Yritystietoa/Terveystalo-Biopankki/Biopankki/). All Finnish Biobanks are members of BBMRI.fi infrastructure (www.bbmri.fi). Finnish Biobank Cooperative -FINBB (https://finbb.fi/) is the coordinator of BBMRI-ERIC operations in Finland. The Finnish biobank data can be accessed through the Fingenious® services (https://site.fingenious.fi/en/) managed by FINBB.

The study was supported by the Wellcome Trust (221854/Z/20/Z), the UK Medical Research Council (S011676, K013351, R024227), the National Institute on Aging (National Institutes of Health; R01AG056477 and RF1AG062553), the British Heart Foundation (RG/16/11/32334), the Academy of Finland (311492), NordForsk (75021), and SomaLogic, Inc. JVL was supported by Academy of Finland (339568). NM was supported by the Academy of Finland (331671). PNS was supported by the Emil Aaltonen Foundation. MK was supported by the Wellcome Trust (221854/Z/20/Z), UK Medical Research Council (MR/S011676, MR/R024227), US National Institute on Aging (R01AG062553, R01AG056477), Academy of Finland (311492), Helsinki Institute of Life Science (H970), and the Finnish Work Environment Fund (190424). SR was funded by Academy of Finland (285380 and 312062), Sigrid Jusélius Foundation and University of Helsinki HiLIFE Fellow grants 2017-2020.

FinnGen consortium authors are listed in FinnGen Banner Authors file.

## Author Contributions

JVL, NM, SR, and MK generated the hypothesis and designed the study. JVL wrote the first draft of the report, did the primary analyses, with support from NM, and performed literature searches. All authors interpreted the data and critically commented and reviewed the report. JVL and MK had full access to pseudonymised data from the Whitehall II. JVL, NM, and SR had full access to pseudonymised data from the FinnGen study.

## Competing interests

In Whitehall II study part of the proteins were measured as academic–industry partnership project beween UCL and SomaLogic, Inc. which provided expertise in plasma proteins and funded 2240 SOMAscan assays. PNS reports a grant from the Emil Aaltonen foundation, during the conduct of the study. MK reports grants from Wellcome Trust, UK Medical Research Council, US National Institute on Aging, Academy of Finland, Helsinki Institute of Life Science, and the Finnish Work Environment Fund, during the conduct of the study.

## Data sharing

Data, protocols, and other metadata of the Whitehall II and FinnGen studies are available according to the data sharing policies of these studies. Pre-existing data access policy for Whitehall II specify that research data requests can be submitted to each steering committee; these will be promptly reviewed for confidentiality or intellectual property restrictions and will not unreasonably be refused. Detailed information on data sharing can be found here: https://www.ucl.ac.uk/epidemiology-health-care/research/epidemiology-and-public-health/research/whitehall-ii/data-sharing Individual-level patient or protein data may further be restricted by consent, confidentiality, or privacy laws/considerations. The FinnGen data may be accessed through Finnish Biobanks’ FinBB portal (www.finbb.fi).

---

**Supplementary Table 1.** Significant backward Mendelian randomization results with p-value below 0.05. These hazard ratios were not stronger than observed in forward Mendelian randomization analyses in Supplementary Figure 1A and B.

| Exposure | Outcome | Number of SNPs | HR (95% CIs) | p-value |
| --- | --- | --- | --- | --- |
| Alzheimer's disease | FSC-A on plasmacytoid dendritic cell | 20 | 0.85 (0.73 to 0.98) | 0.0275 |
| Late-onset Alzheimer's disease | Protein S100-A13 | 13 | 0.51 (0.41 to 0.63) | <0.001 |

**Supplementary Figure 1A.**
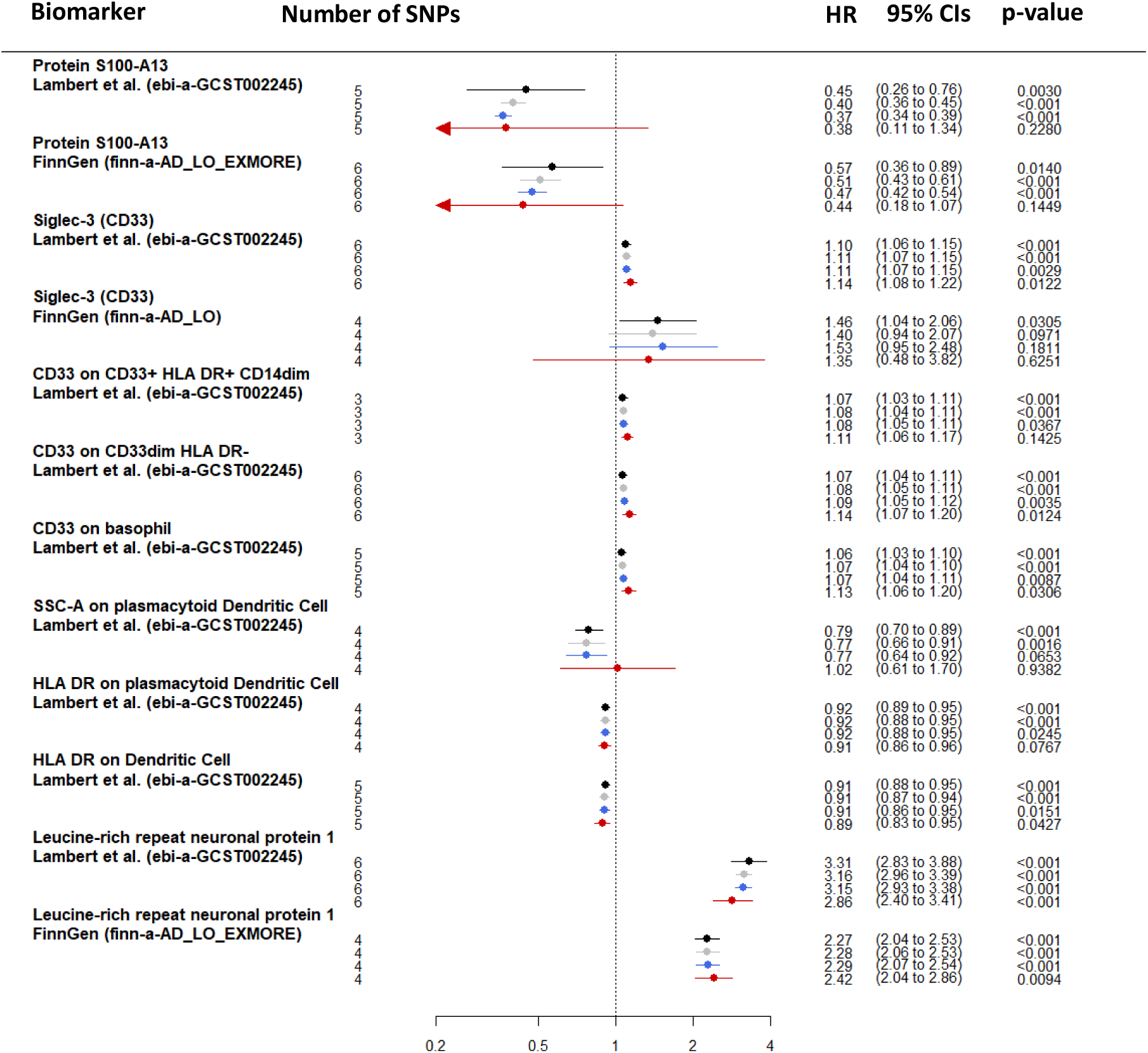
Mendelian randomization sensitivity analyses for one standard deviation change in biomarker levels associated with late onset Alzheimeŕs diseases and with at least 3 SNPs available. The source of outcome and MR-base outcome identifier is described below the biomarker. Black = inverse variance weighted, grey = weighted median, blue = simple mode, red = Egger Mendelian randomization.

**Supplementary Figure 1B.**
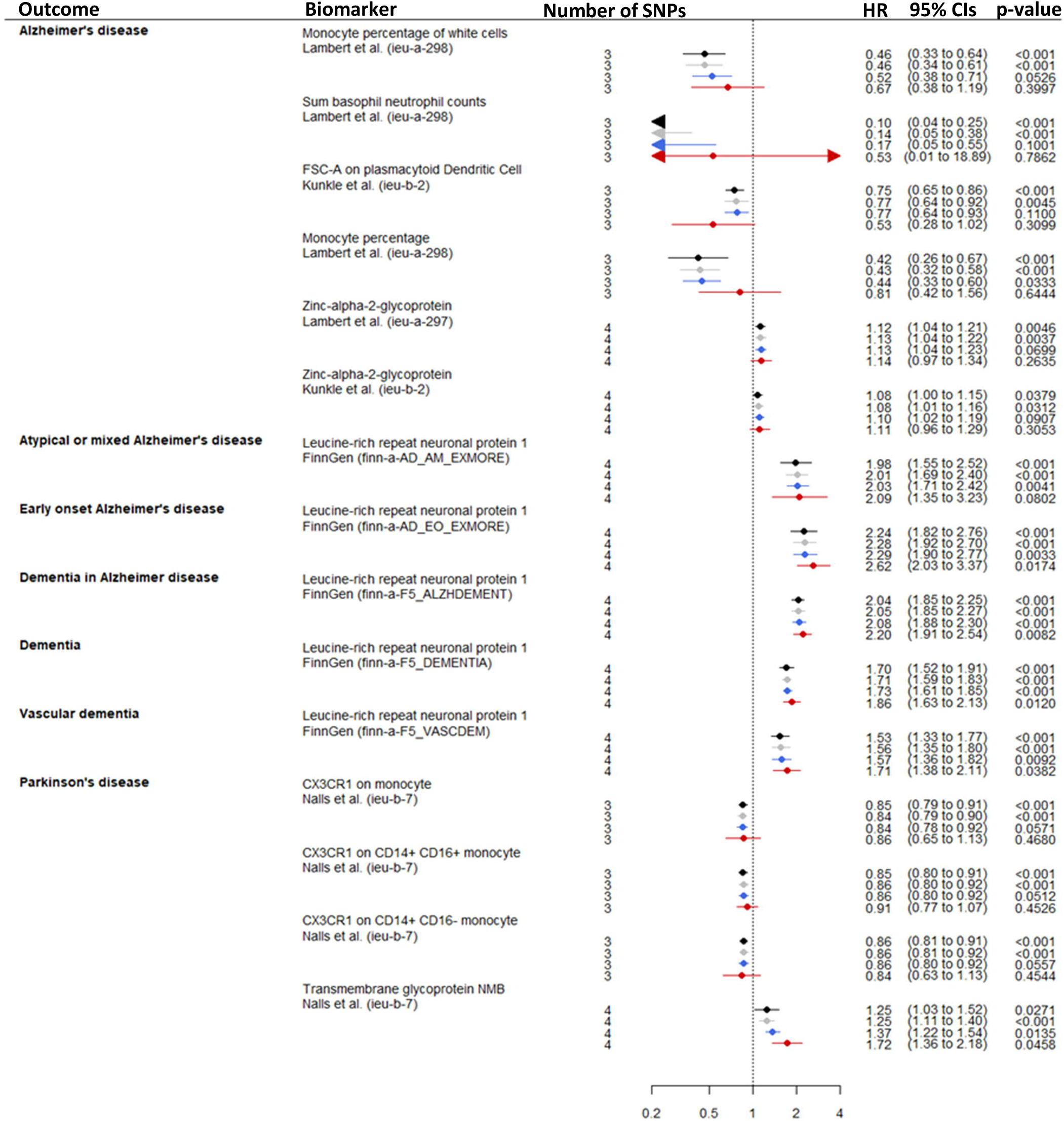
Mendelian randomization sensitivity analyses for one standard deviation change in biomarker levels associated with other dementias than late onset Alzheimeŕs diseases and with at least 3 SNPs available. The source of outcome and MR-base outcome identifier is described below the Biomarker. Black = inverse variance weighted, grey = weighted median, blue = simple mode, red = Egger Mendelian randomization.

**Supplementary Figure 2.**
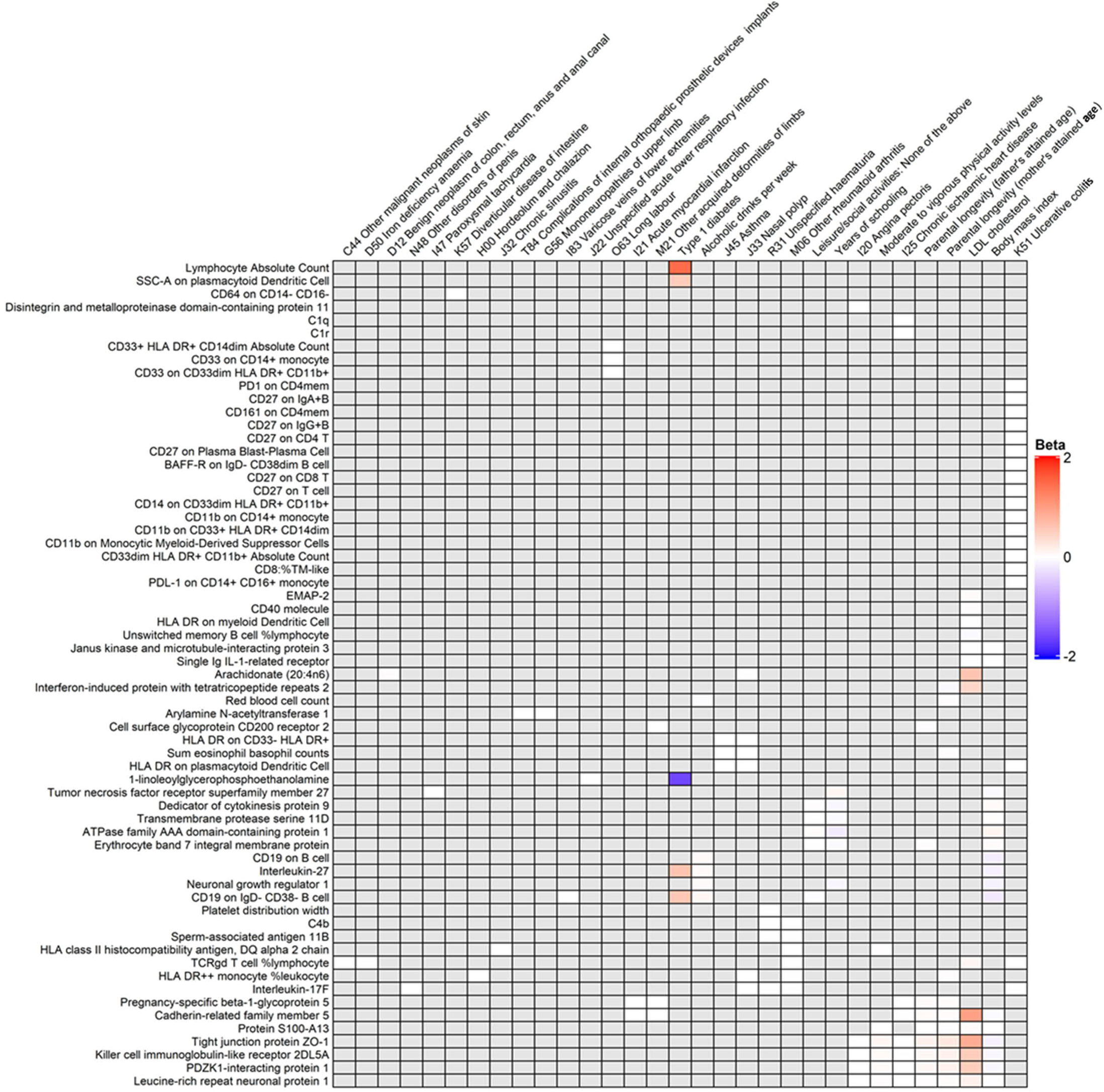
Phenome-wide association analyses for the 126 biomarkers, effect size describes one standard deviation change in biomarker levels. All biomarker and outcomes pairs that passed false discovery rate correction of 5% are presented. Most biomarkers associated with only few outcomes and the grey boxes indicate no association after false discovery rate correction of 5% or lack of common SNPs.

**Supplementary Figure 3.**
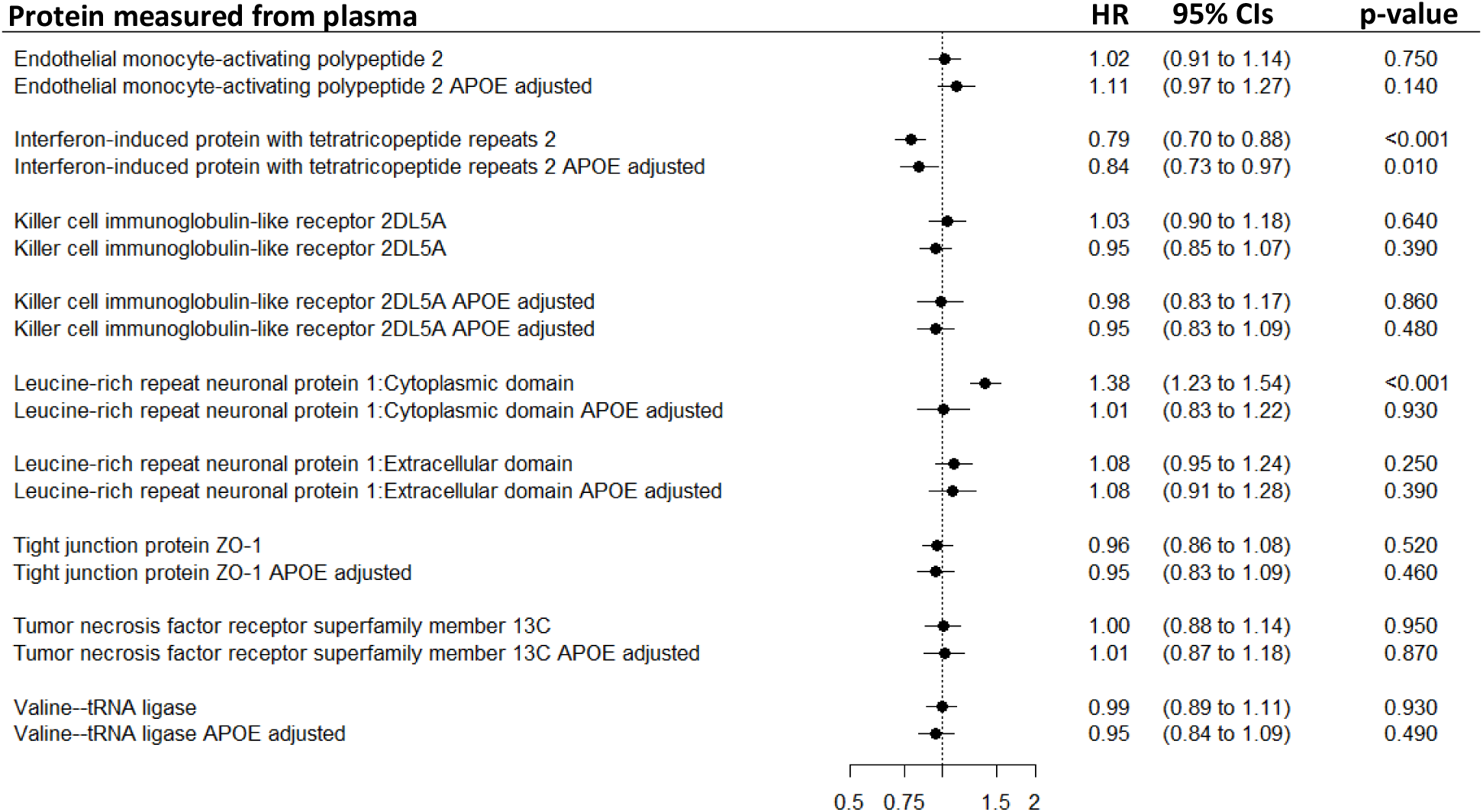
Plasma protein associations with dementia for one standard deviation change in protein levels from the Whitehall II cohort. The analyses included 8 proteins that had SNPs clustered around *APOE* and associated with at least three dementia subtypes in Mendelian randomization analyses. Analyses are first adjusted for age and sex and then additionally for *APOE* status.

**Supplementary Figures 4 A-H.**
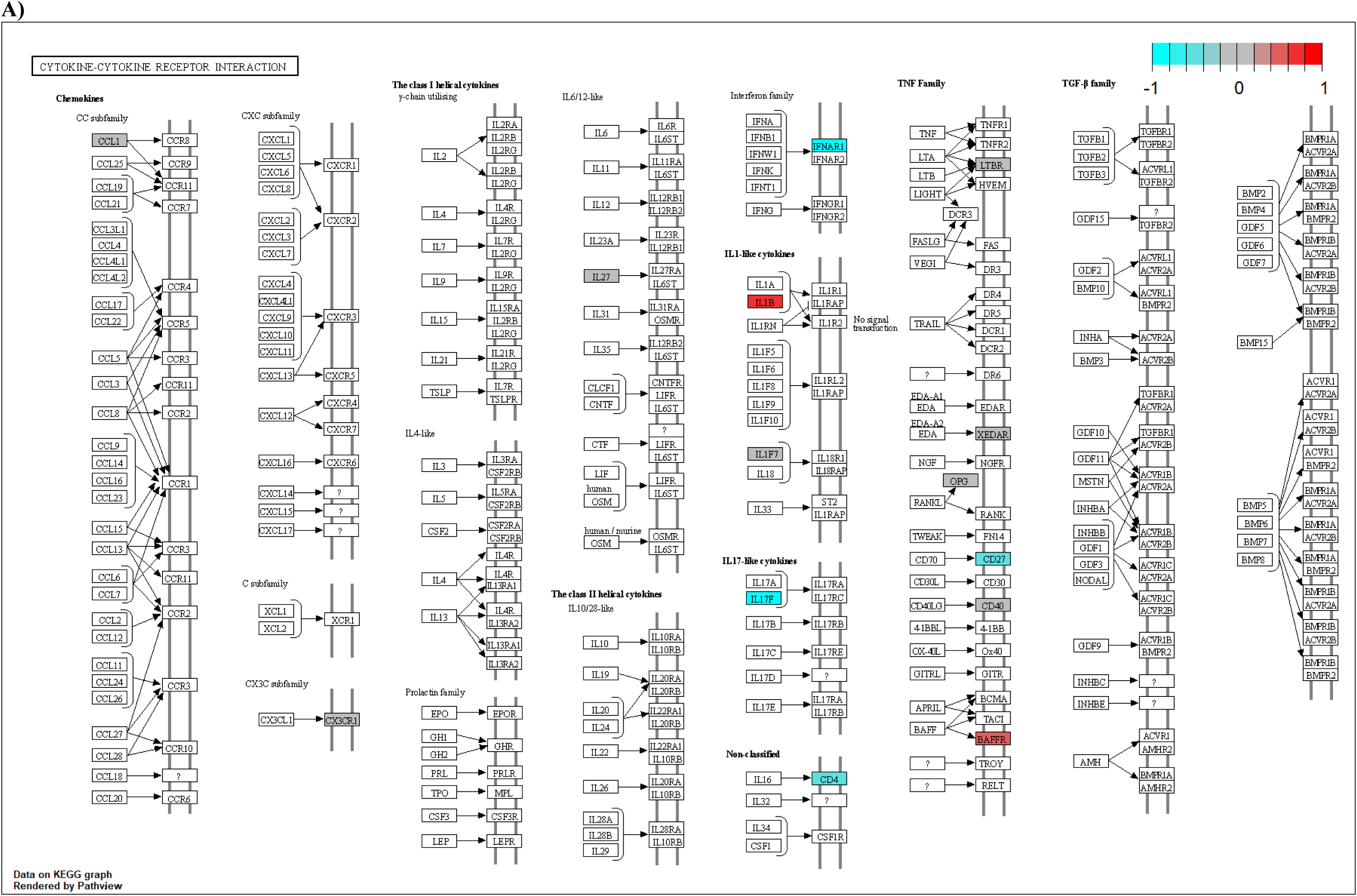
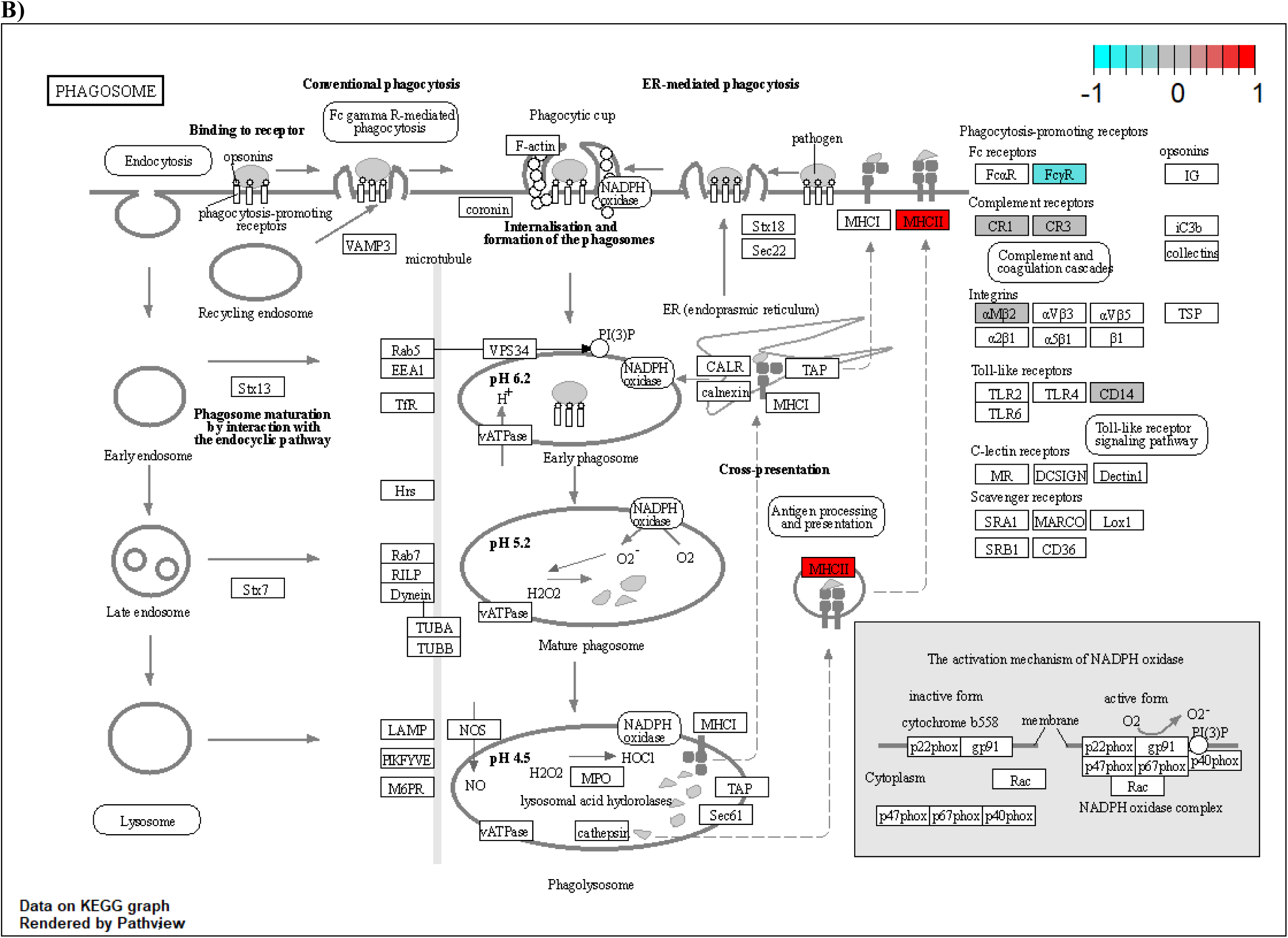
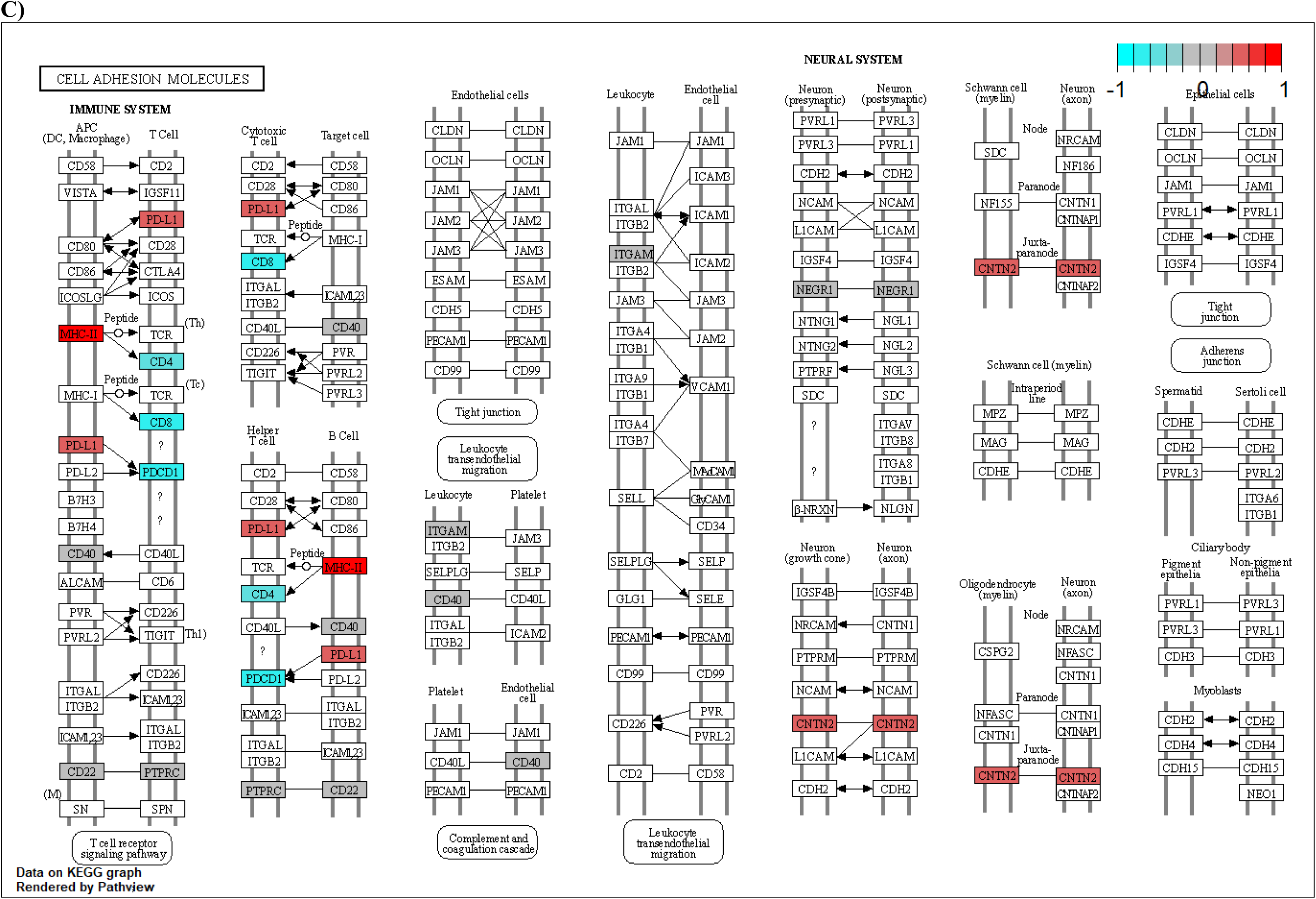
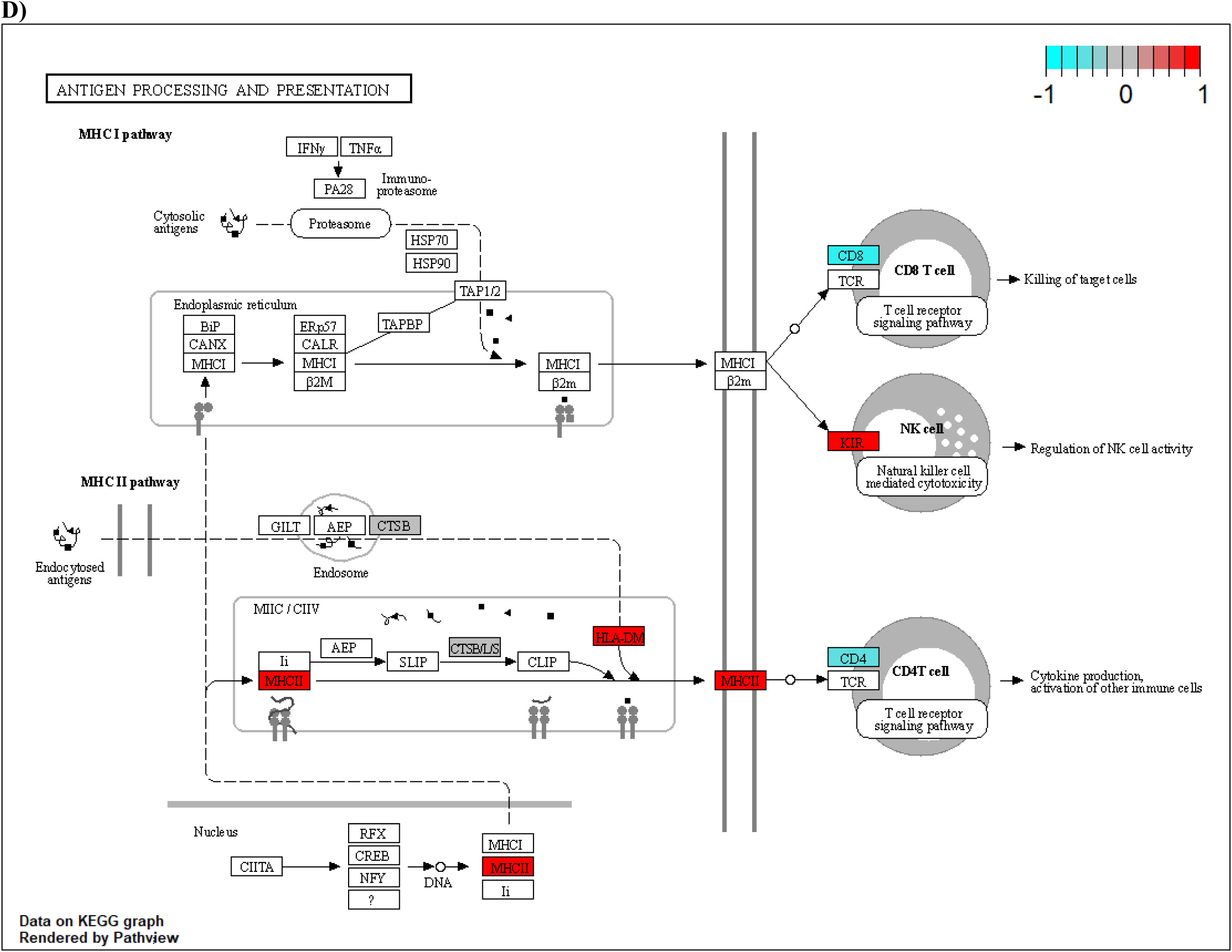
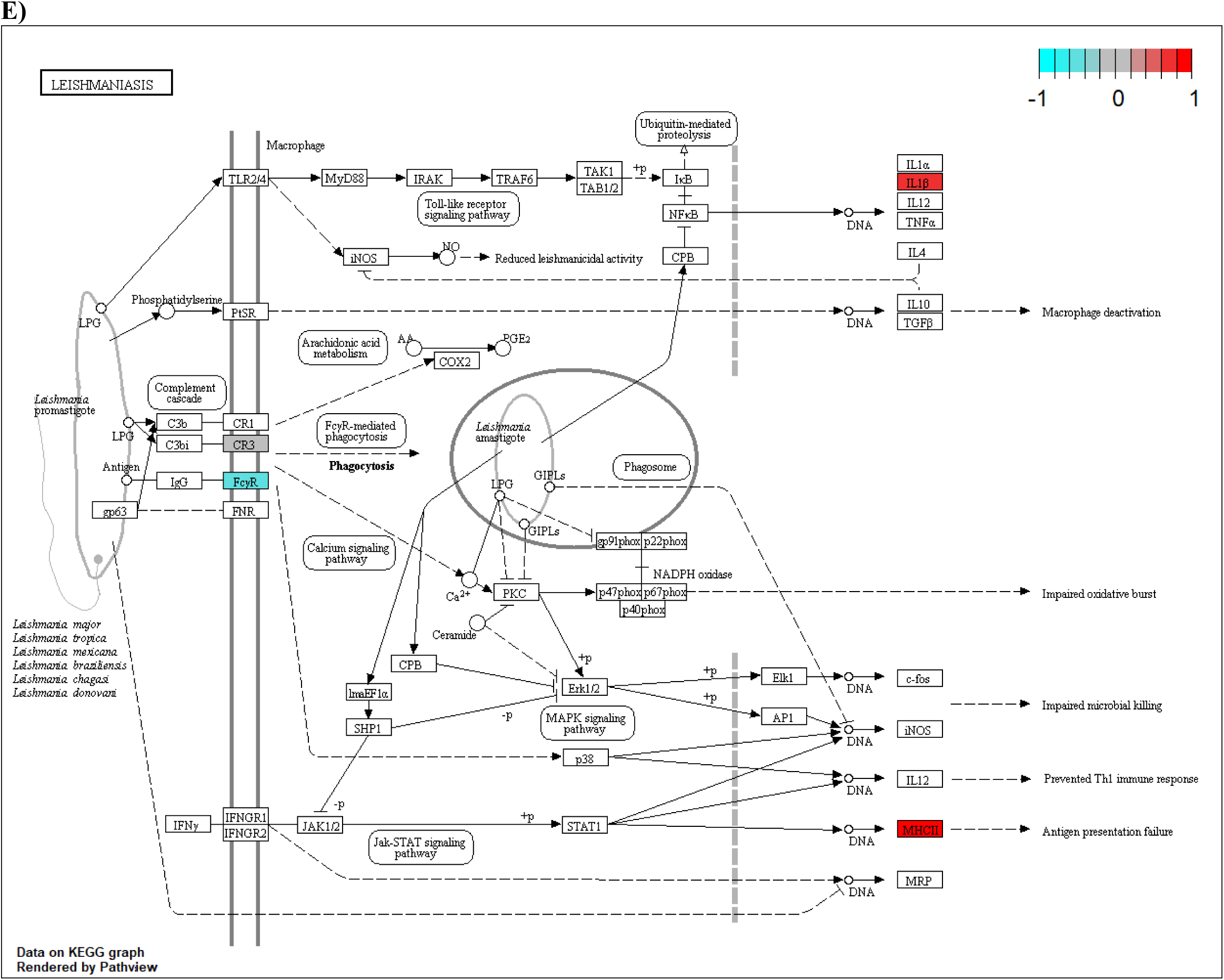
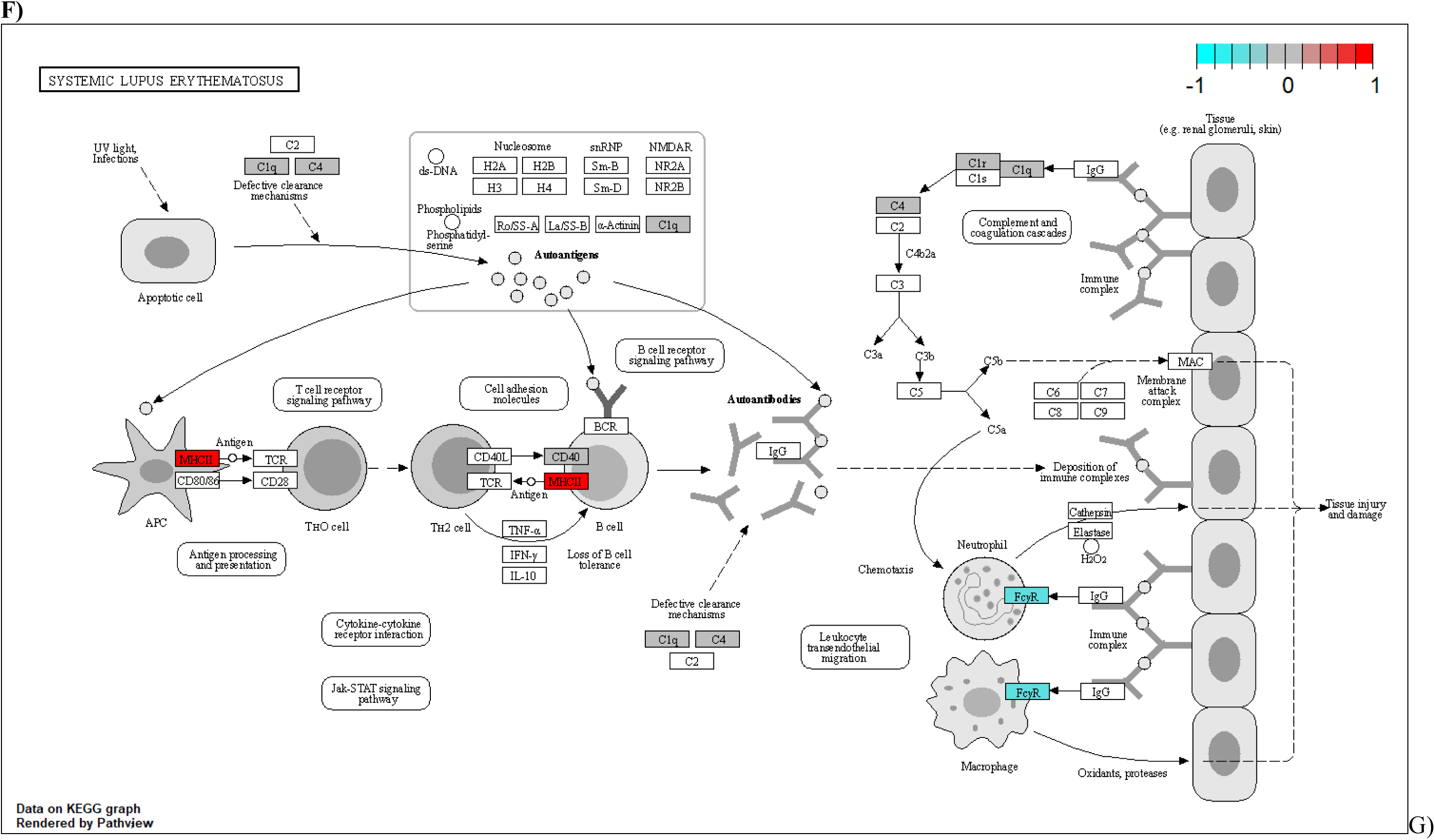
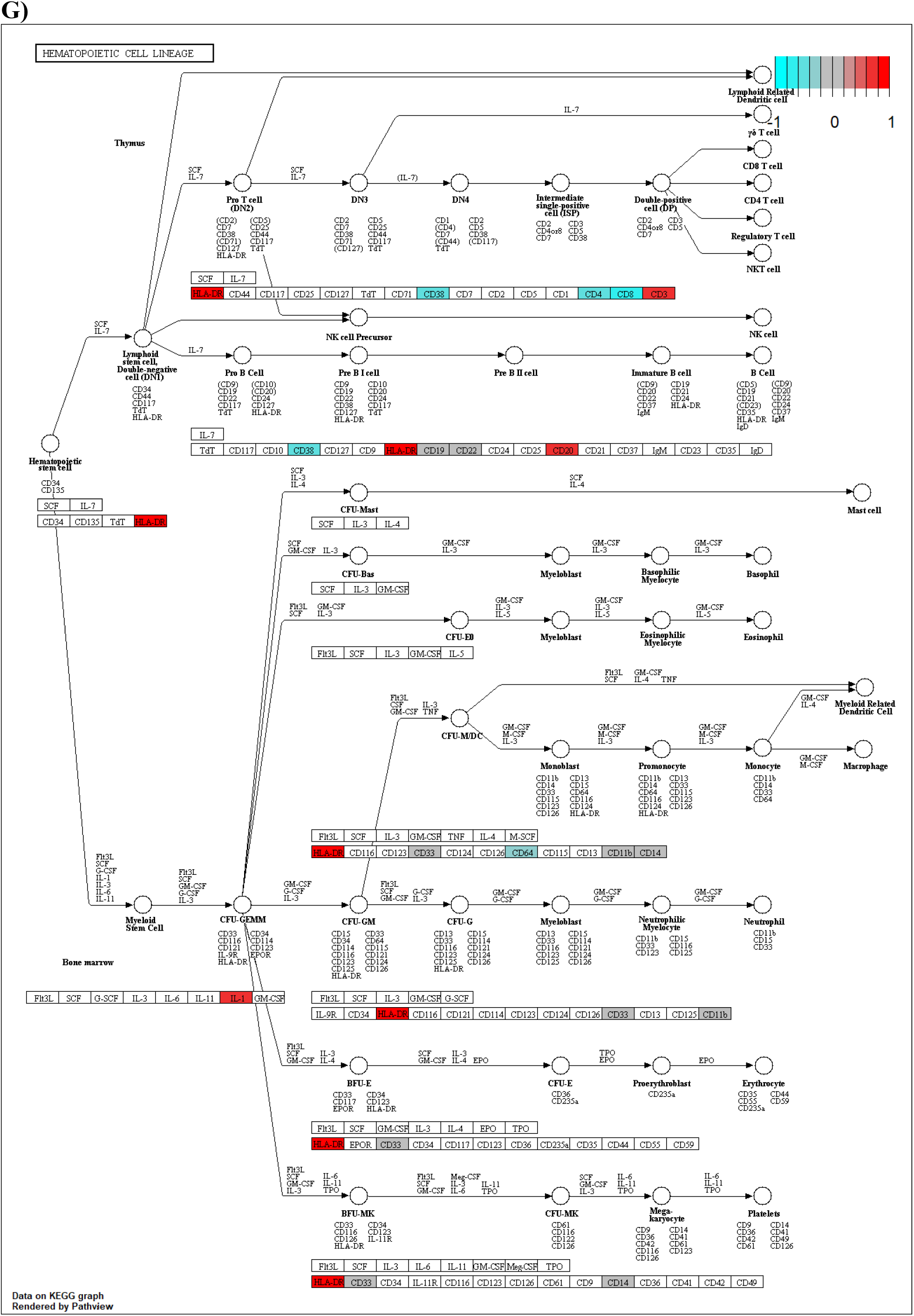
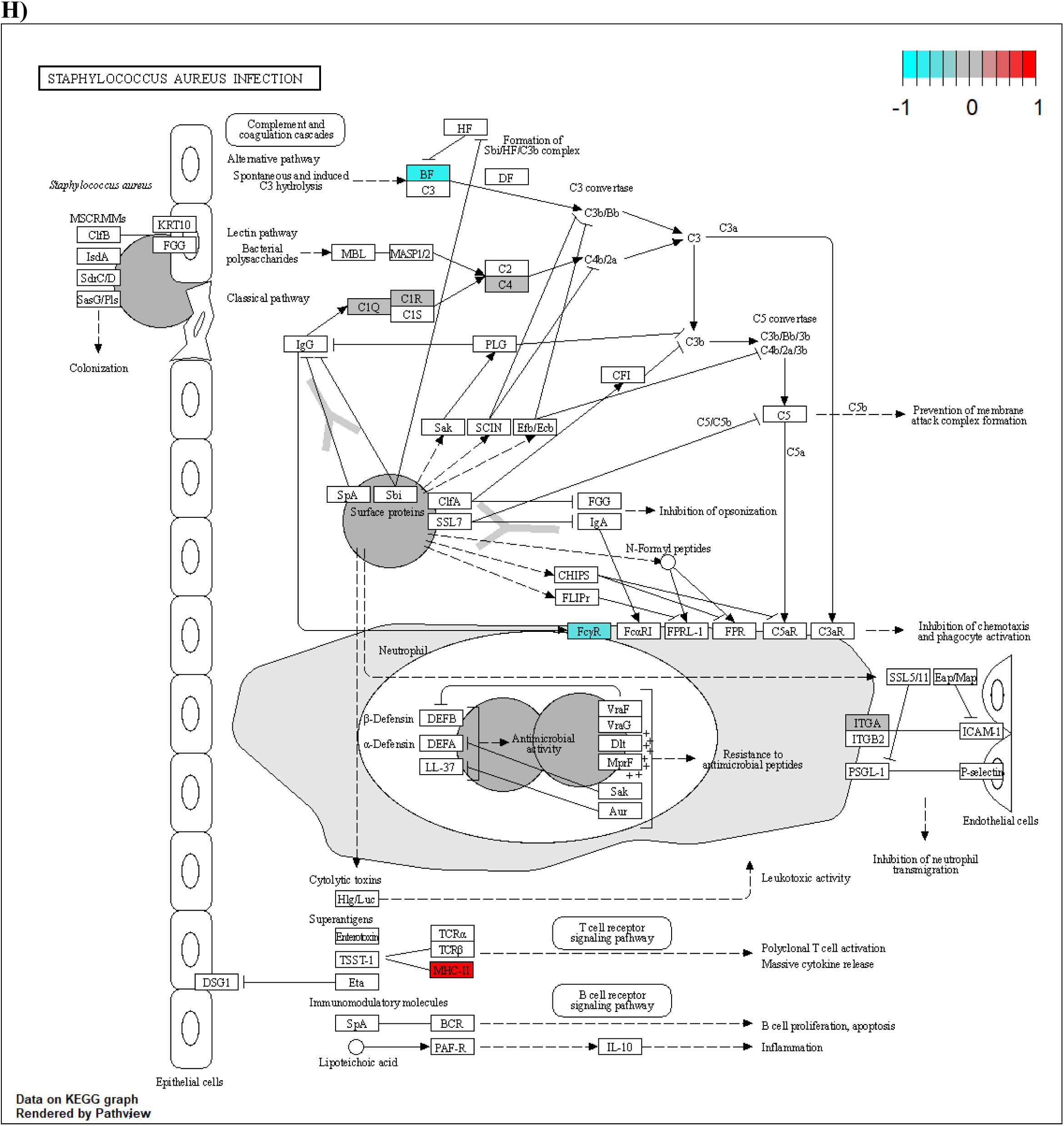
KEGG functional pathway enrichment analyses for all proteins and receptors that associated with dementias in Mendelian randomization analyses. The estimated causal betas were used in place of log fold change.

**Supplementary Figure 5.**
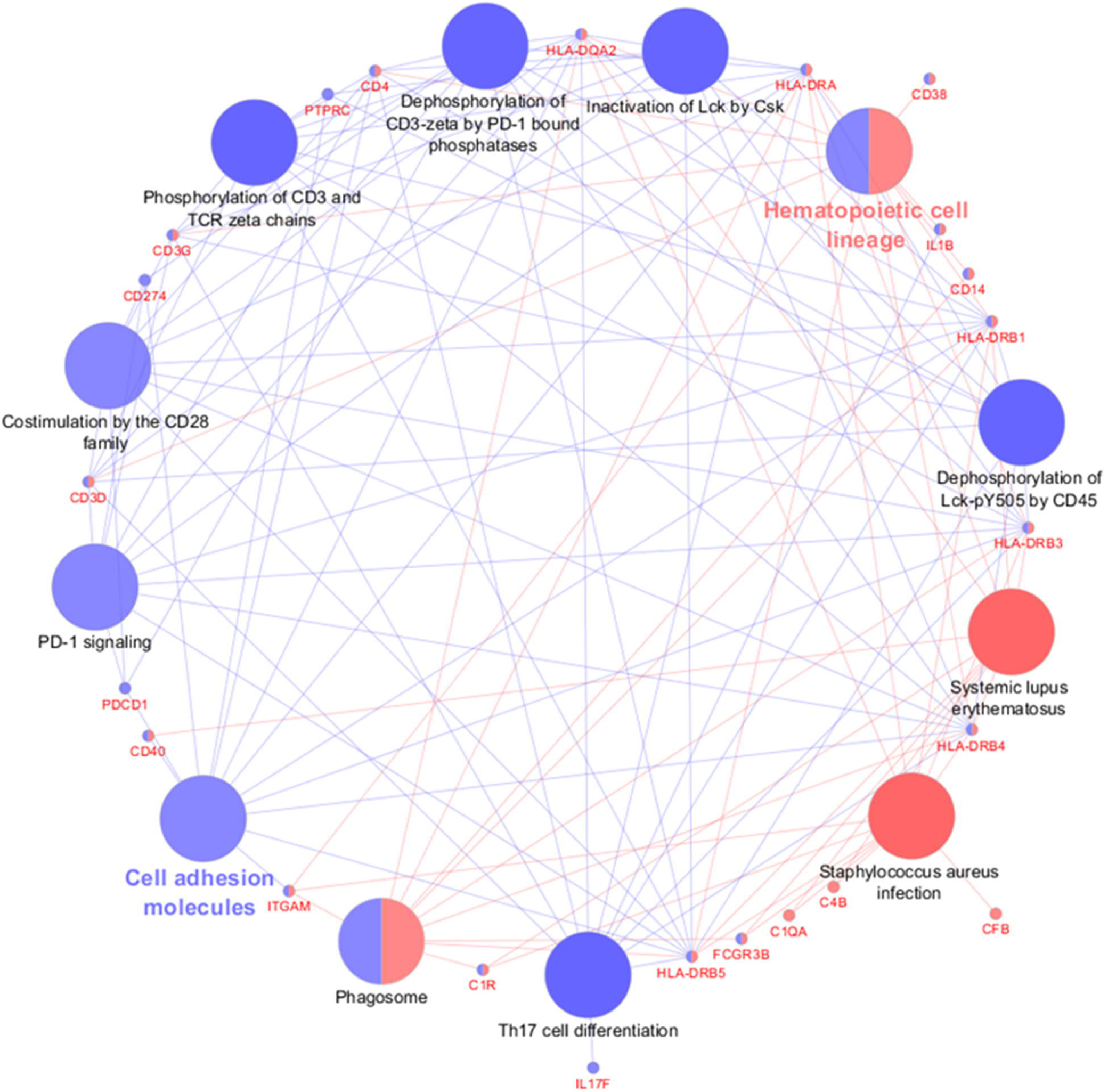
ClueGO enrichment analyses for all the 76 proteins and receptors in the biomarkers that had evidence of causal association in Mendelian randomization analyses. All proteins that associated with Uniprot search terms “immune system” and “blood brain barrier” were used as a background dataset. GO-terms in blue were enriched in cluster where most significant associations were observed for cell adhesion molecules. In the red cluster, the most significant GO-term associations were observed for hematopoietic cell lineage. Big dots describe the GO-terms and small dots the genes associated with these terms.

**Supplementary Figures 6A-R.**
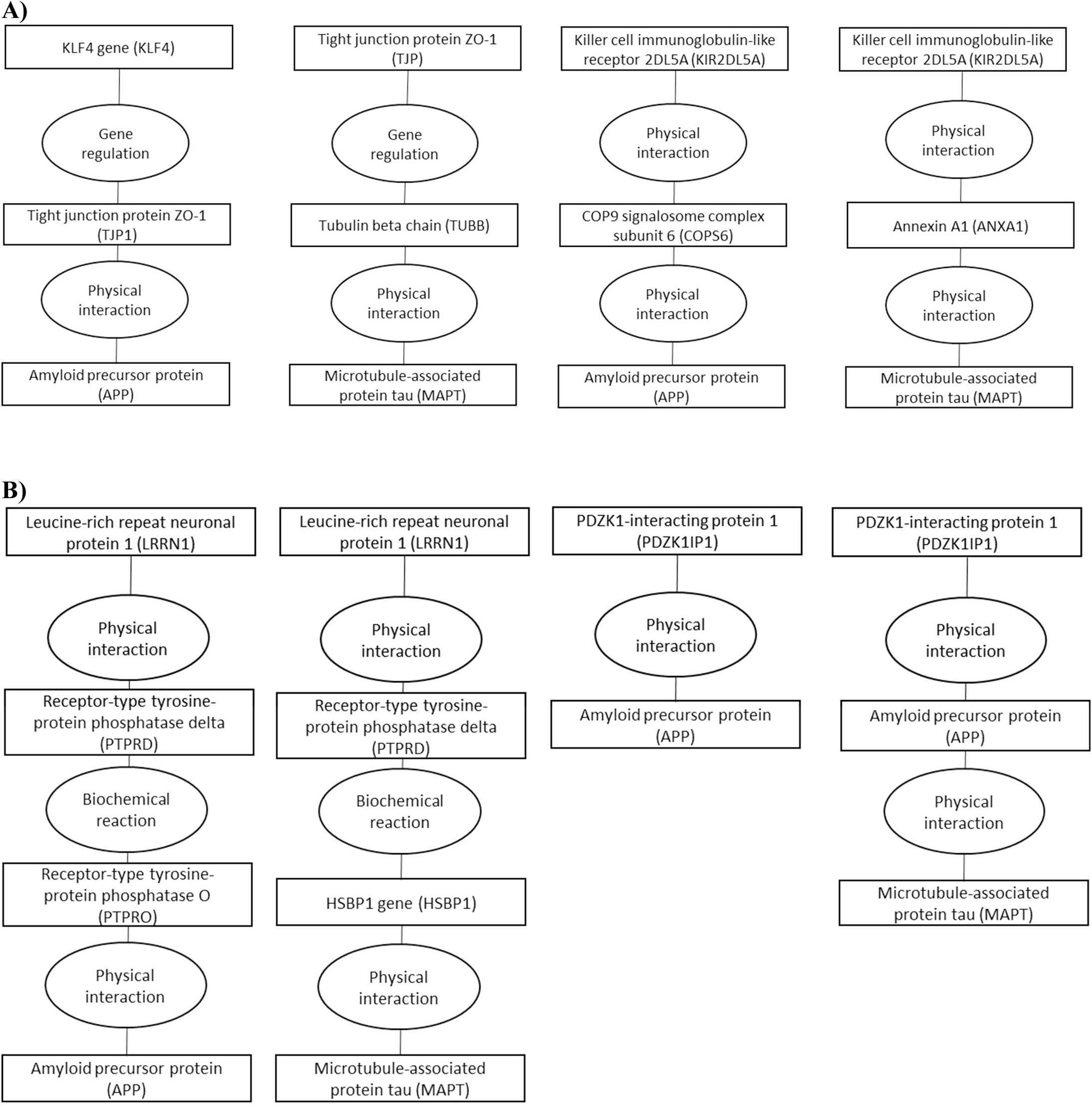
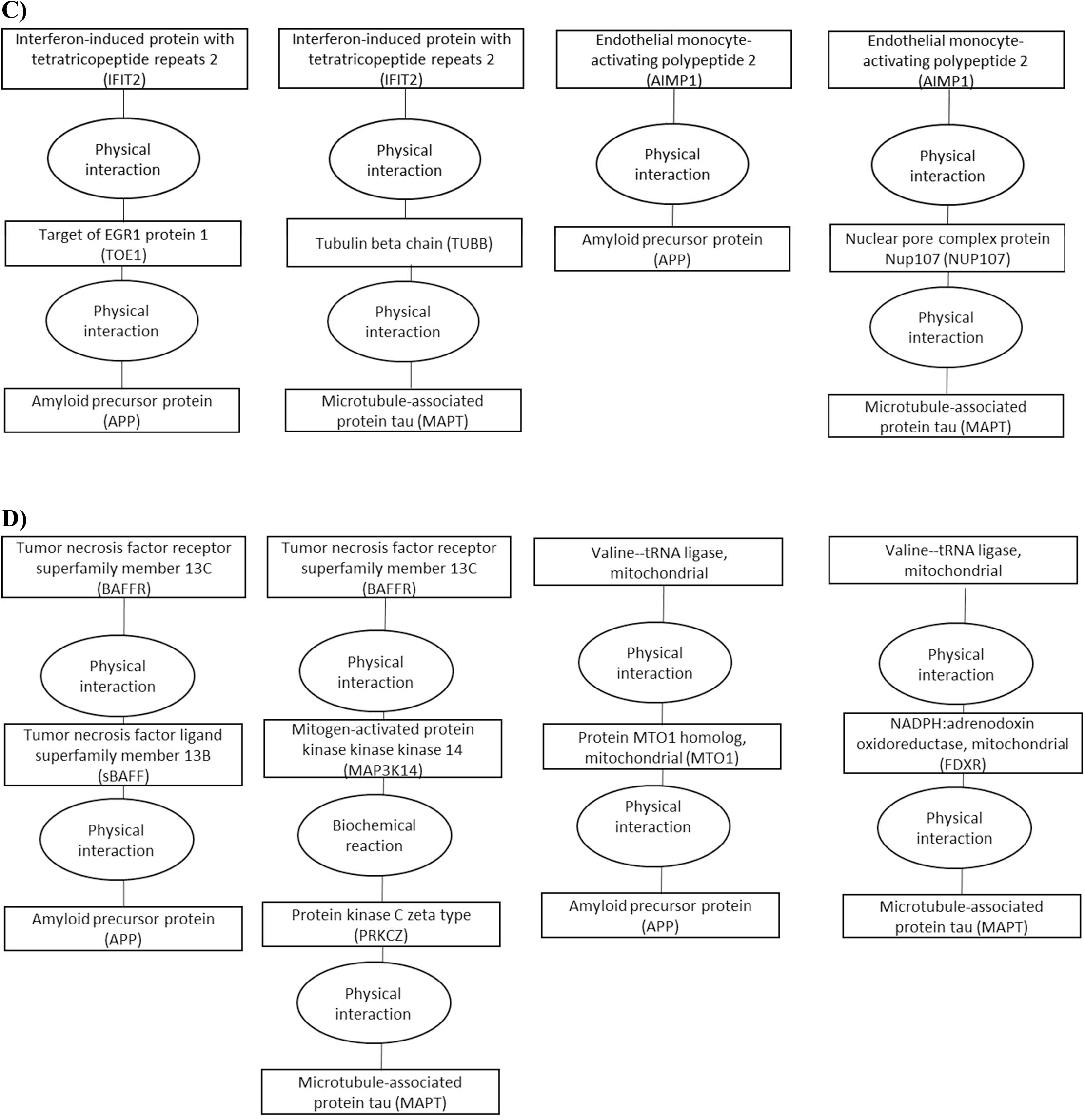
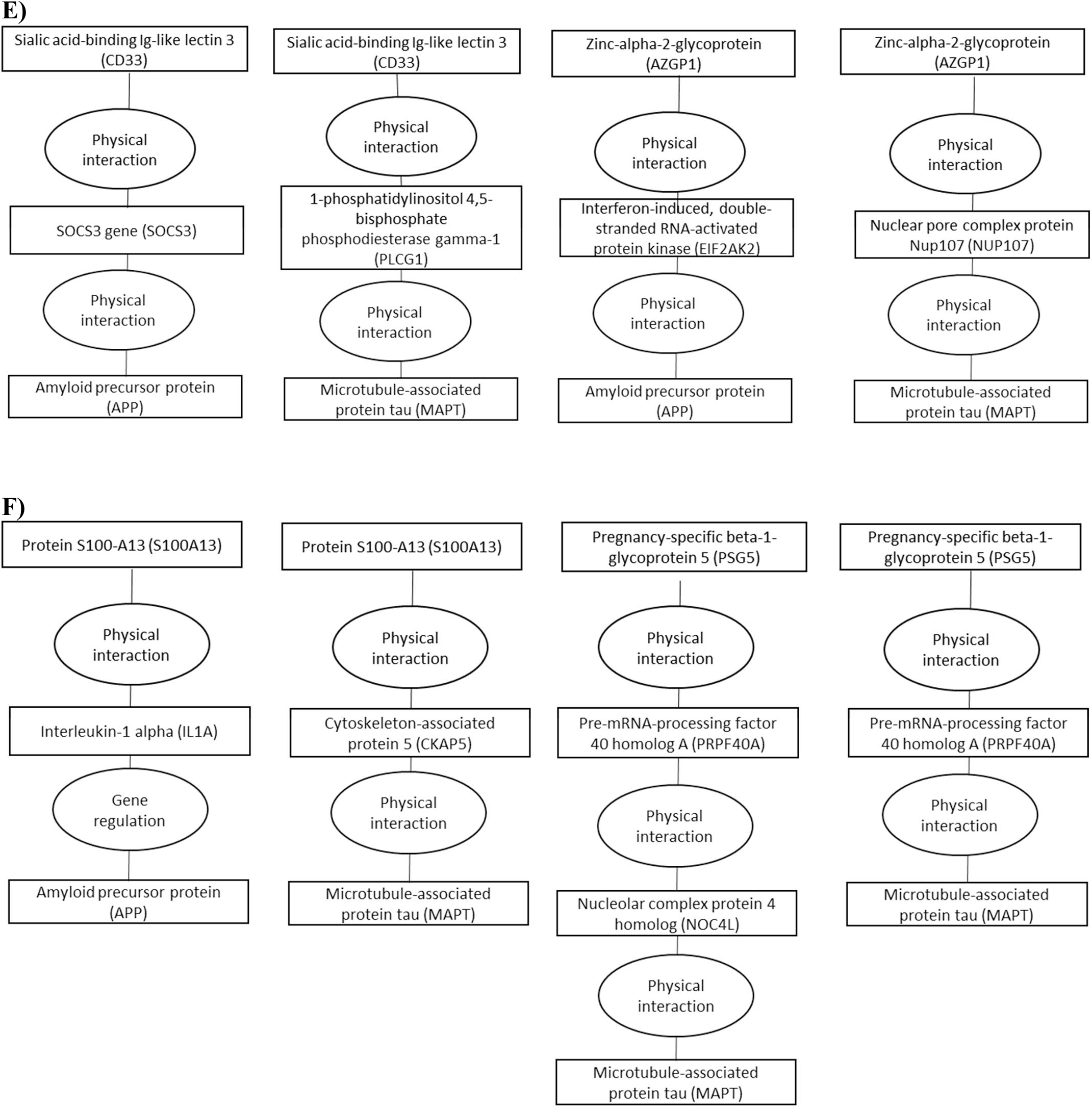
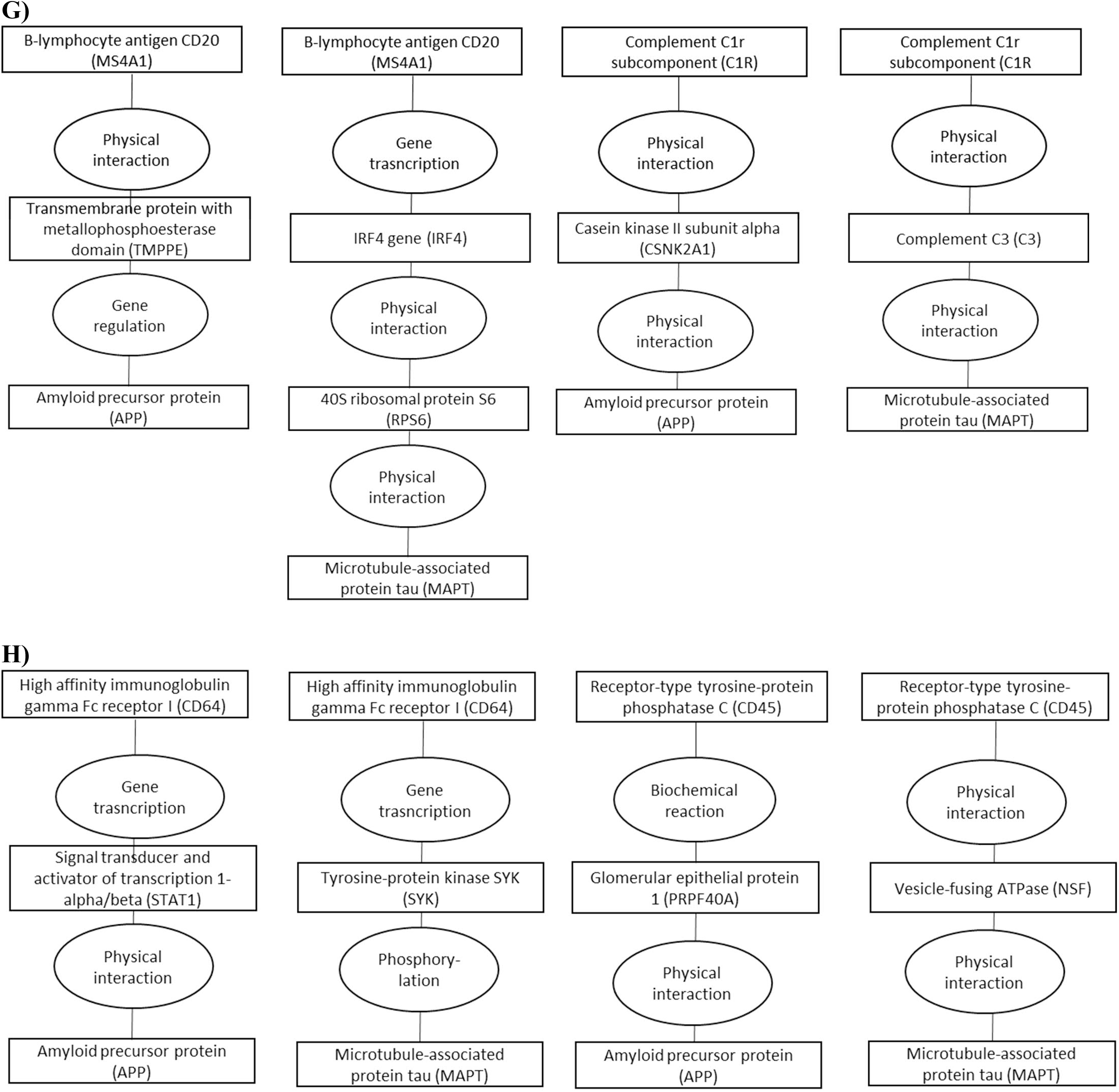
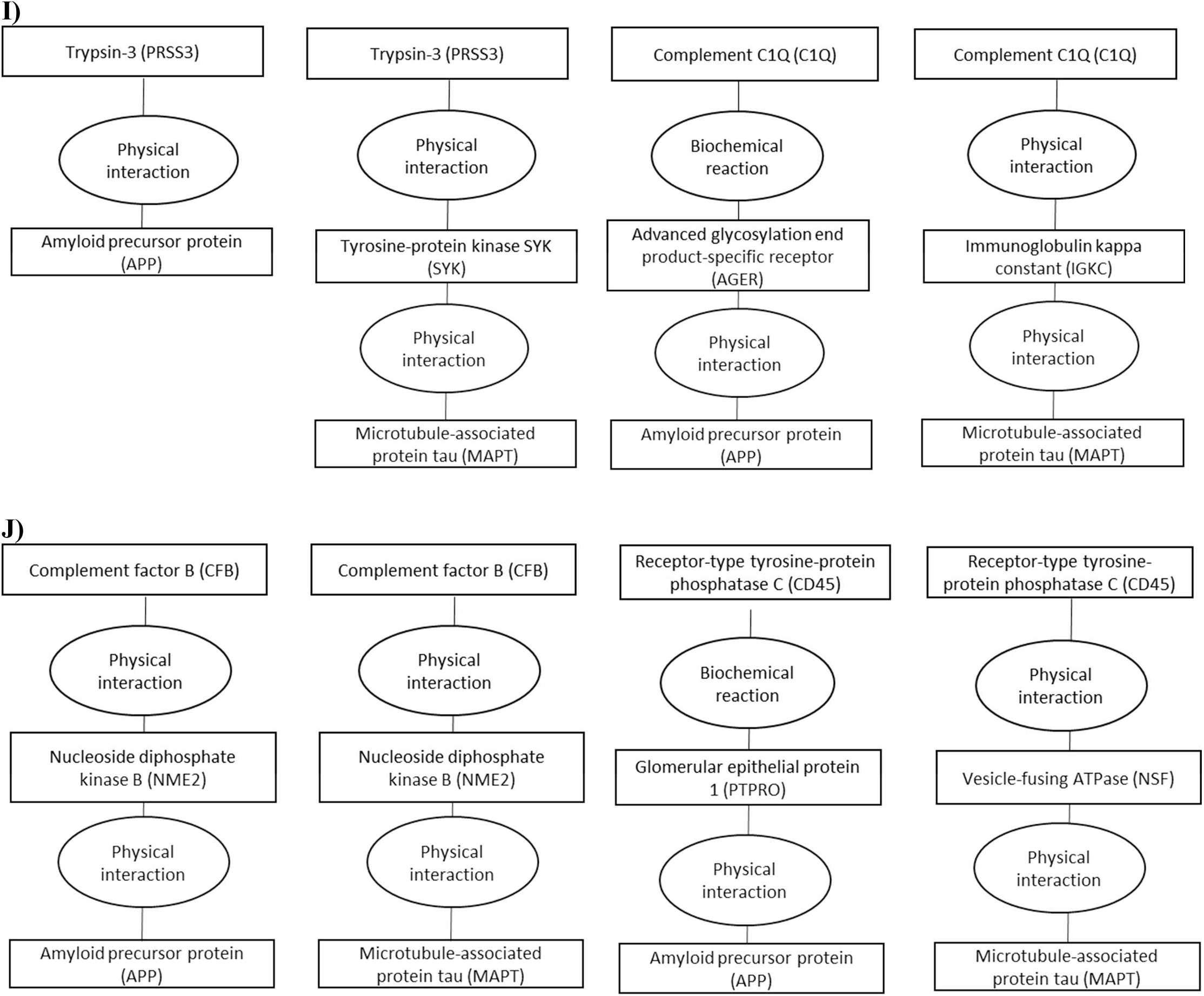
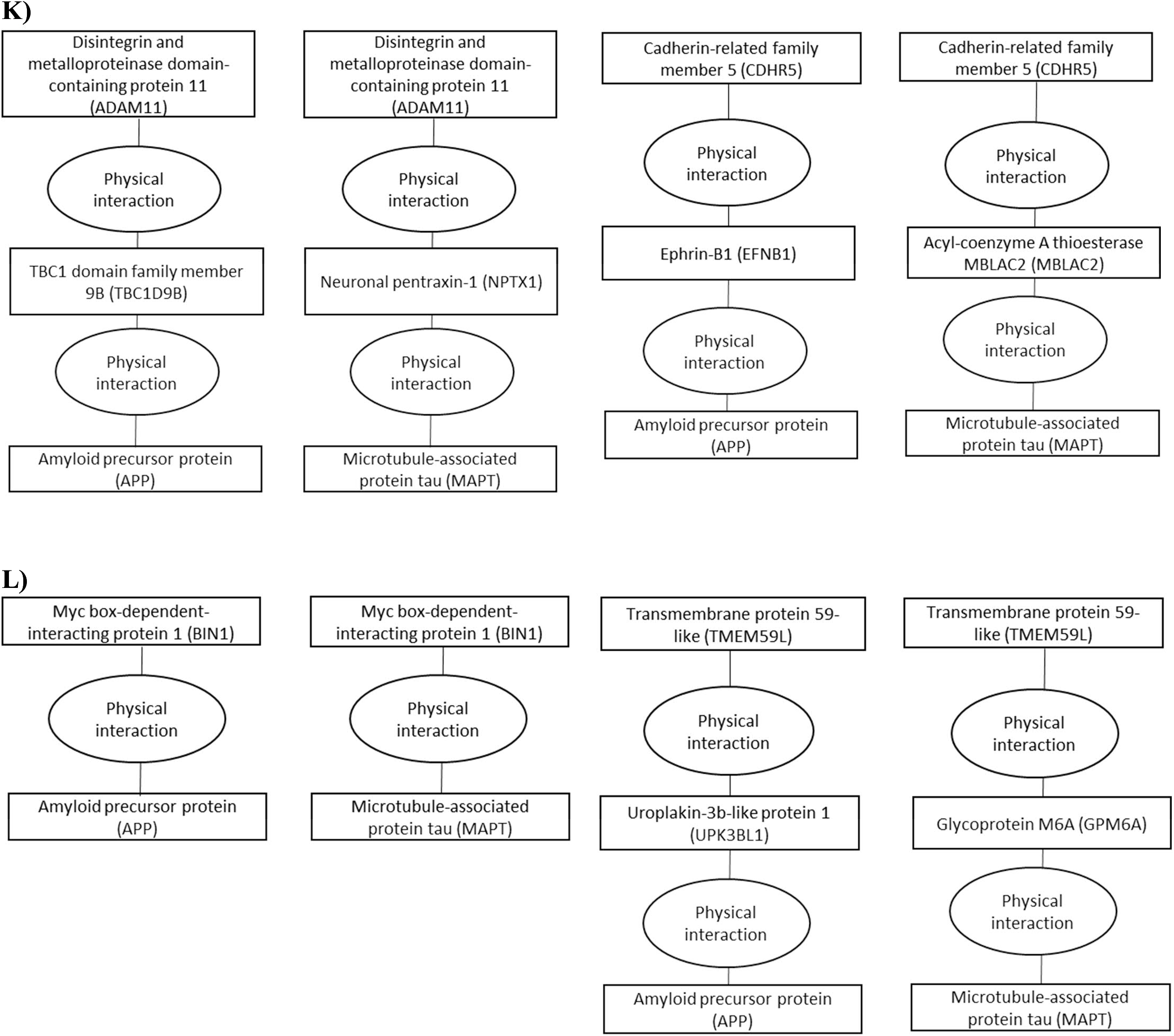
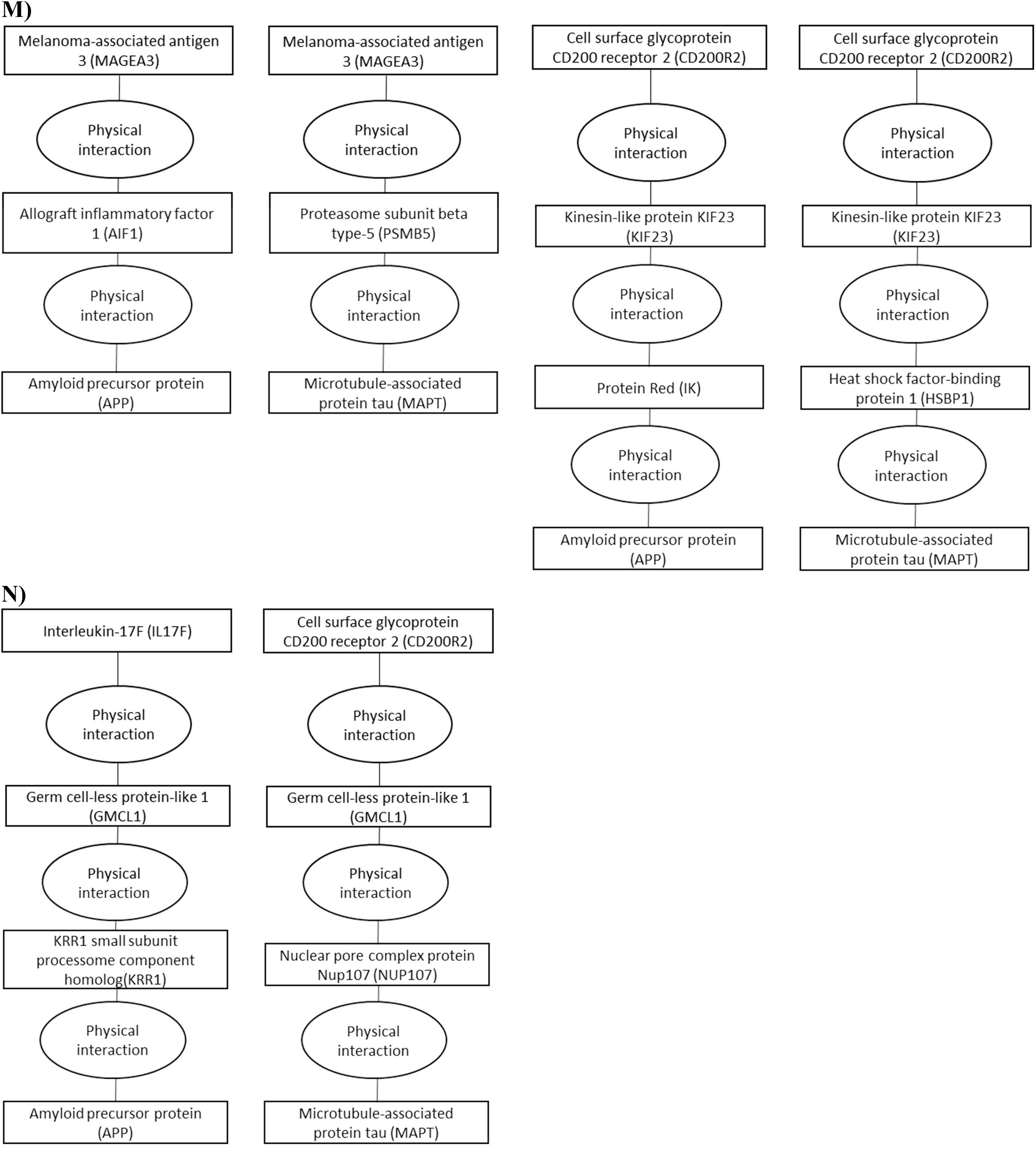
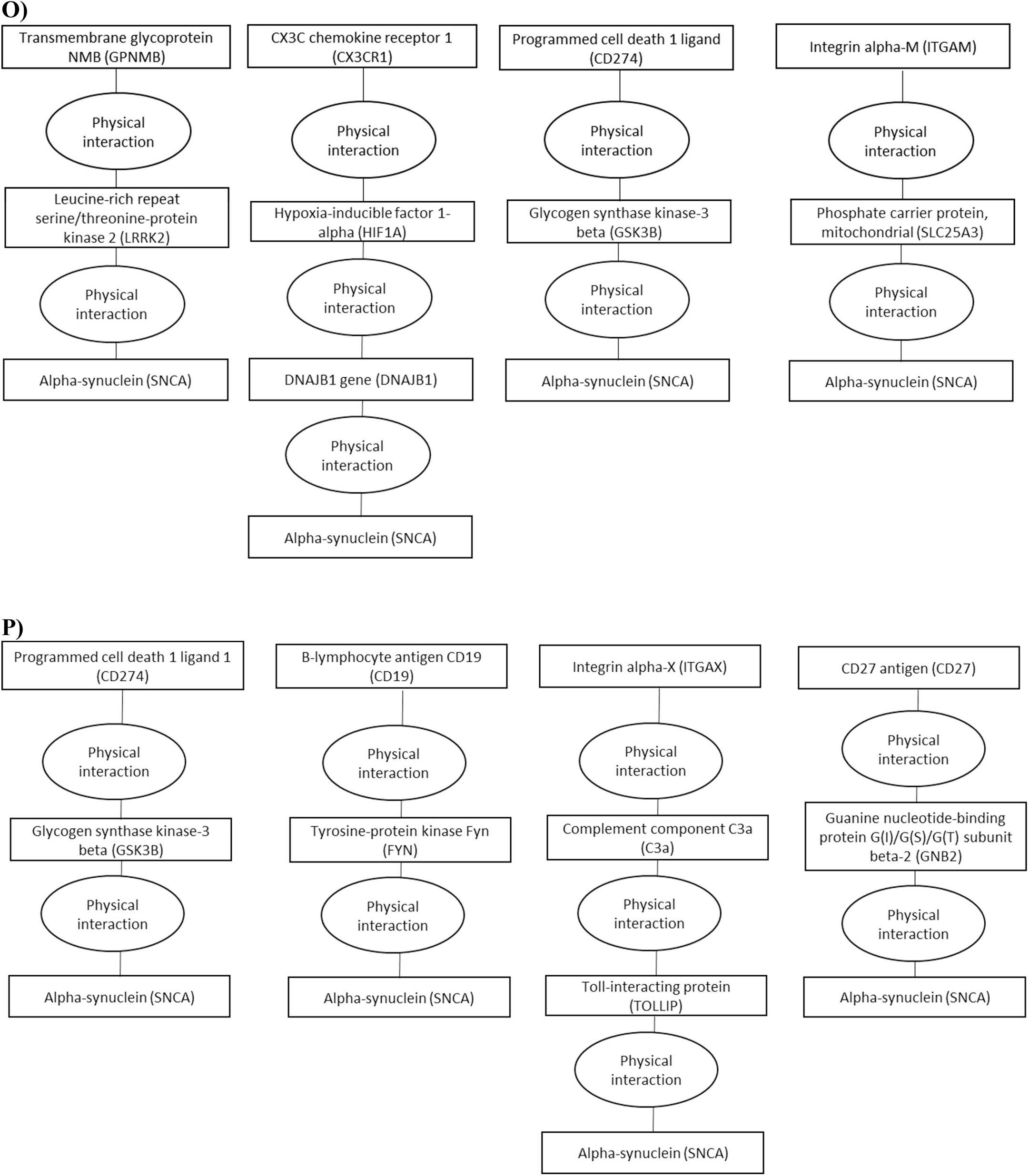
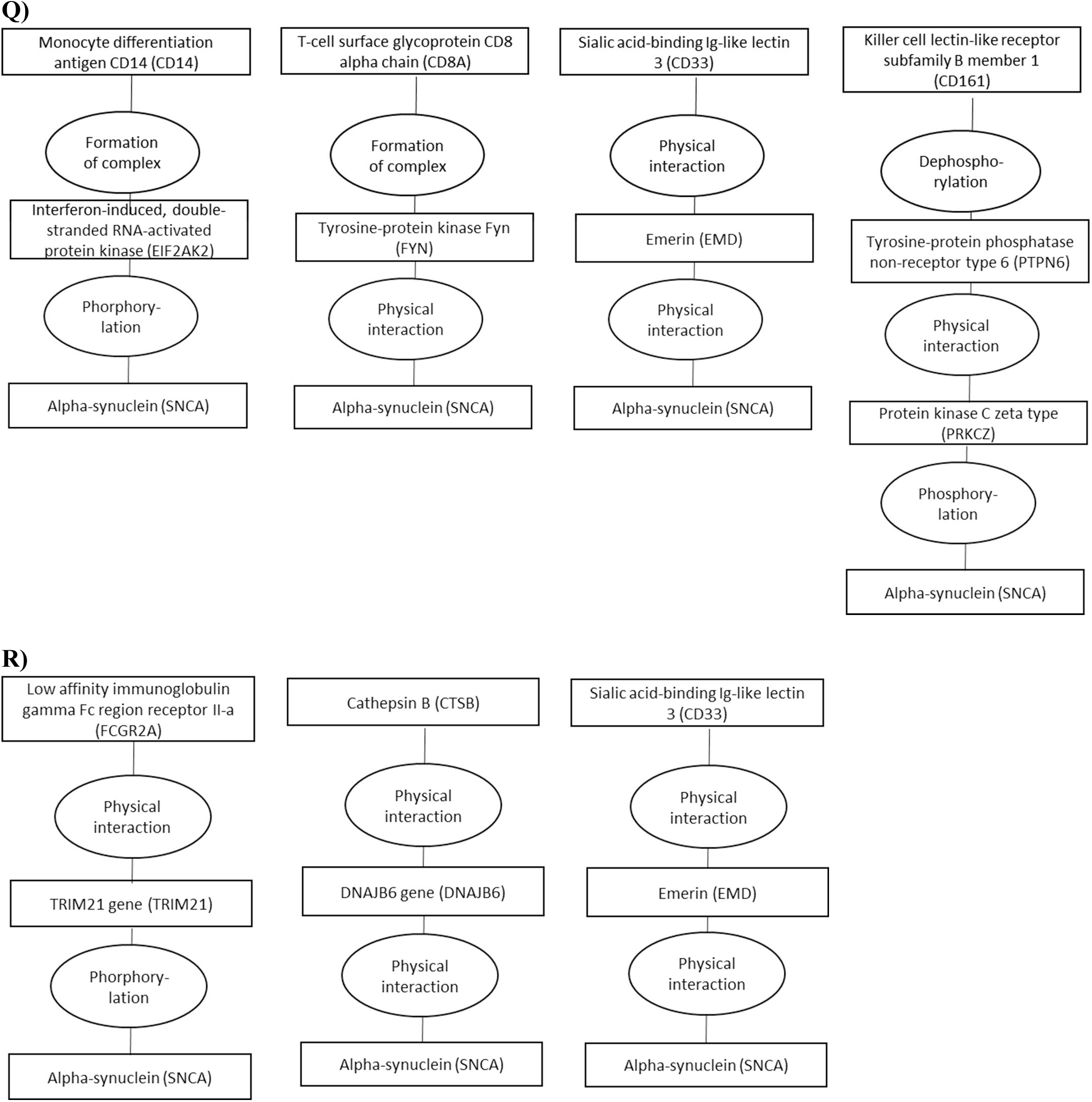
ConsensusPathDB shortest interaction path analyses for the 42 proteins that were causally associated with dementia causing diseases in Mendelian randomization analyses. Figures A-N include biomarkers for amyloid and tau and O-R for α- synuclein pathways.

## Supplementary methods

### Mendelian randomization analyses

The SNPs for biomarkers and outcomes were searched from MR-base database.^1^ Immune system and BBB search terms were identified using identifiers of cell types, receptors, proteins, and genes. The identifiers were searched from several publications^2–10^ and from Uniprot^11^ with search terms “immune” and “blood brain barrier”. Outcomes included all cohorts with following dementia-causing diseases: all types of Alzheimer’s disease, Parkinson’s disease, vascular dementia, frontotemporal dementia, dementia in general, and progression of dementia. Poor cognitive performance was chosen as an intermediate outcome. Two sample Mendelian randomization was used to analyse associations between biomarkers and outcomes.^12^ First analyses estimated effects with Wald ratio or inverse variance-weighted analyses.^1^ For biomarker-outcome pairs that passed FDR of 5% and shared over two common SNPs we performed sensitivity analyses with weighted median, simple mode, MR Egger, and backward Mendelian randomization^12^ with TwoSampleMR and MRInstruments R packages.

To assess off-target effects of the observed causal biomarkers, we performed a phenome-wide Mendelian randomization analyses for each biomarker, using GWAS summary statistics for 210 UK Biobank endpoints conducted by Neale lab and with recognized dementia risk factors.^5, 13^ All analyses used European ancestry as reference, clumping cut-off R2 = 0.01 and a 10,000kb window. LD proxies were searched with R2 = 0.6 and a proxy split size of 500. The biomarkers and outcome alleles were harmonized by inferring from positive strand alleles using allele frequencies for palindromes. We used statistical software R (3.6.0 and 4.1.0) for these analyses. Novelty of our Mendelian randomization findings was evaluated with Pubmed search using following search terms: (Mendelian randomization) AND (dementia OR Alzheim* OR Parkin* OR cognitive decline) AND (Entrez gene symbol OR Uniprot protein identifier) without limitations.

### Plasma proteins and Whitehall II study

Plasma protein measurements in Whitehall II study were available for 6,235 individuals of whom 310 developed dementia. The participants are linked to the National Health Services (NHS) Hospital Episode Statistics (HES) database, and the British National mortality register using individual NHS identification numbers for linkage.^14^ The NHS provides nearly complete health care coverage for all individuals legally resident in the UK. Incident dementia was defined using the WHO International Classification of Diseases, version 10 (ICD-10) codes F00, F01, F03, G30, and G31 and ICD-9 codes 290.0-290.4, 331.0-331.2, 331.82, and 331.9. We also conducted informant interviews and checked participants’ medications at each screening (in 1996-1998, 2011-2013 and 2016-2017) for dementia-related medication. Sensitivity and specificity of dementia assessment based on HES data is 0.78 and 0.92.^15^

The plasma proteins were measured using the SomaScan version 4.0 and 4.1 assays.^4, 16, 17^ The assays were validated against external reference population and protein-specific conversion coefficients were used to balance the technical differences between versions 4.0 and 4.1. The analyses used plasma samples measured in 1995/1997 and stored in 0.25 mL aliquots at −80◦C. Earlier studies describe performance of the SomaScan assay and the modified aptamer binding in detail.^4, 16, 17^ In brief, the assay uses a mix of thousands of slow off-rate modified aptamers (SOMAmers) that bind to proteins in participants’ plasma samples where the specificity is ensured with two-step process analogous to a conventional immunoassay. The specificity of the aptamer reagents is good and has been tested in several ways.^4, 18, 19^ Median intra- and inter-assay coefficients of variation for SomaScan version 4 are ∼5% and assay sensitivity is comparable to that of typical immunoassays, with a median lower limit of detection in the femtomolar range.

In the Whitehall II study, standard self-administered questionnaires provided data on age and sex. Using DNA extracted from whole blood, a standard PCR assay determined *APOE* genotype using the salting out method.^20, 21^ Two blinded independent observers read the genotype and any discrepancies were resolved by repeating the PCR analysis.

In analyses, proteins were transformed to a normal distribution by inverse rank-based normal transformation. The follow-up ended at onset of dementia, death, or 1^st^ of October 2019, which ever occurred first. Age, sex, and *APOE* adjusted Cox regression models estimated associations between the proteins and dementia.^22^ The proportionality assumption in all Cox models was assessed with Schoenfeld residuals and with log-log plots.^22^ We used statistical software R (3.6.0 and 4.1.0) for these analyses.

### Polygenic risk score and IPW analyses

FinnGen Data Freeze 8 contains 339,233 individuals, and represents approximately 7% of Finnish adult population. FinnGen is a collection of prospective epidemiological and disease-based cohorts, and hospital biobank samples and links genotypes by the unique national personal identification numbers to nationwide health registries, including the national hospital discharge (available from 1968), death (1969–), cancer (1953–) and medication reimbursement (1964–) and purchase (1995–) registries. The registry-based follow-up ended on Dec 31, 2019. Alzheimer’s disease was defined with ICD-10 codes under F00 and G30, ICD-9 codes under 3310, ICD-8 code under 29010, and medication purchase code N06D. Vascular dementia was defined with ICD-10 codes under F01 and ICD-9 codes under 4378. Parkinson’s disease was defined with ICD-10 codes under G20, ICD-9 codes under 3320A, ICD-8 code under 34200, and reimbursements code 110.

FinnGen samples were genotyped with Illumina and Affymetrix arrays (Illumina Inc., San Diego, and Thermo Fisher Scientific, Santa Clara, CA, USA), and genotype calls were made with the GenCall or zCall (for Illumina) and the AxiomGT1 algorithm for Affymetrix data. Individuals with ambiguous gender, high genotype missingness (>5%), excess heterozygosity (+-4SD) and non-Finnish ancestry were excluded, as well as all variants with high missingness (>2%), low Hardy– Weinberg equilibrium p-value (<1e-6) and minor allele count (MAC < 3). Array data pre-phasing was carried out with Eagle 2.3.5^23^ with the number of conditioning haplotypes set to 20,000. Genotype imputation was done using the population-specific SISu v3 imputation reference with 3,775 high-coverage (25-30x) whole-genome sequences in Finns, described in detail at https://doi.org/10.17504/protocols.io.xbgfijw.

The PRS were constructed from SNPs that were associated with the Mendelian randomization causal 126 biomarkers for dementias. The SNPs were LD-pruned with clumping cut-off R2 = 0.01 in 500kb window with R package TwoSampleMR. The available SNPs were searched from FinnGen genotypes and the final scores were generated with PLINK v2.00aLM3 by calculating the SNP-biomarker beta weighted sum of risk alleles for each SNP. The PRS was scaled to zero mean and unit variance, and we analysed its phenome-wide associations across 2401 disease endpoints. The association between PRS and endpoints were studied with logistic regression, adjusting for birth year, sex, and ten first principal components of ancestry.

We used IPW analyses to emulate randomized control trials on the effect of preventive anti-inflammatory medication on risk of dementias in FinnGen.^24^ These analyses included participants aged over 45 without any dementia-causing disease at baseline (N = 90,512) ATC codes for anti-inflammatory medication use were searched from medication purchase registry that starts from 1995. To simulate trial design and to avoid immortal time bias, each analyses included only new medication users. In IPW analyses, we assumed that when medication is initiated, it is continued until end of follow-up to simulate intention to treat analyses and thus to provide conservative estimates. The baseline variables in IPW analyses were birth year, sex, ten principal components of ancestry and time-varying variables statin, ACE-blocker, AT-blocker, renin-blocker, calcium channel blocker, any diuretic, insulin, metformin, other diabetes drug, depression medication, antipsychotic, and anticoagulant use as well as time varying any cancer, myocardial infarction, atrial fibrillation, heart failure, venous thromboembolism, ischemic stroke, intracerebral haemorrhage, subarachnoid haemorrhage, obesity, sleep apnoea, and chronic obstructive pulmonary diseases, with informative censoring included. The positive control analyses on statin medication used these same variables but did not include statin as time-varying covariate. We used R (4.1.2) for these analyses.

### HLA analyses

The analyses were done using imputed HLA alleles, imputed with high accuracy using a Finnish-specific reference panel, which has been previously described in detail.^25^ After filtering based on a HLA carrier frequency of ≥0.01 and posterior probability of ≥0.6, we assessed the association between HLA alleles and dementias and autoimmune diseases using logistic regression, adjusting for birth year, sex, and ten first principal components of ancestry.

### Pathway analyses

KEGG pathway analysis was used for studying the effect of the biomarkers on validated pathways with Generally Applicable Gene-set Enrichment (GAGE),^26^ using Mendelian randomization Wald ratios or inverse variance weight betas and p-values as input for expression ratios. GO term enrichment analyses were done with ClueGO version 2.5.8^27^ in Cytoscape version 3.7.2.^28^ The hypergeometric test used all the 76 plasma proteins or surface receptors of the 126 biomarkers as input and all immune system and BBB related proteins from Uniprot^11^ as background, with correction for 5% FDR. The shortest interaction path analyses were done with ConsensusPathDB,^29^ a web based analysis tool containing a range of biomedical databases. ConsensusPathDB was used to decipher the potential common pathways between the biomarkers and amyloid β, tau protein, and α-synuclein.

---

**Supplementary file 1.**
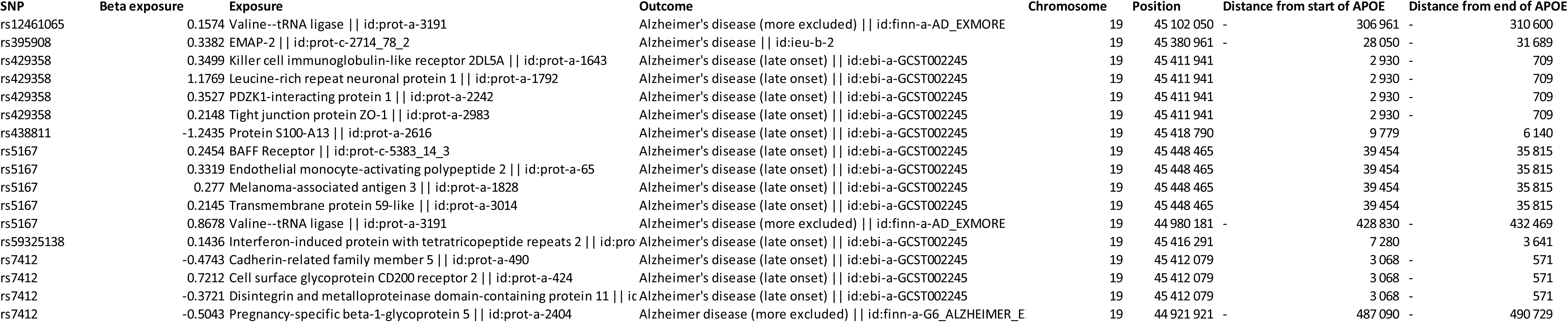
Distance from *APOE* gene.

**Supplementary File 2.**
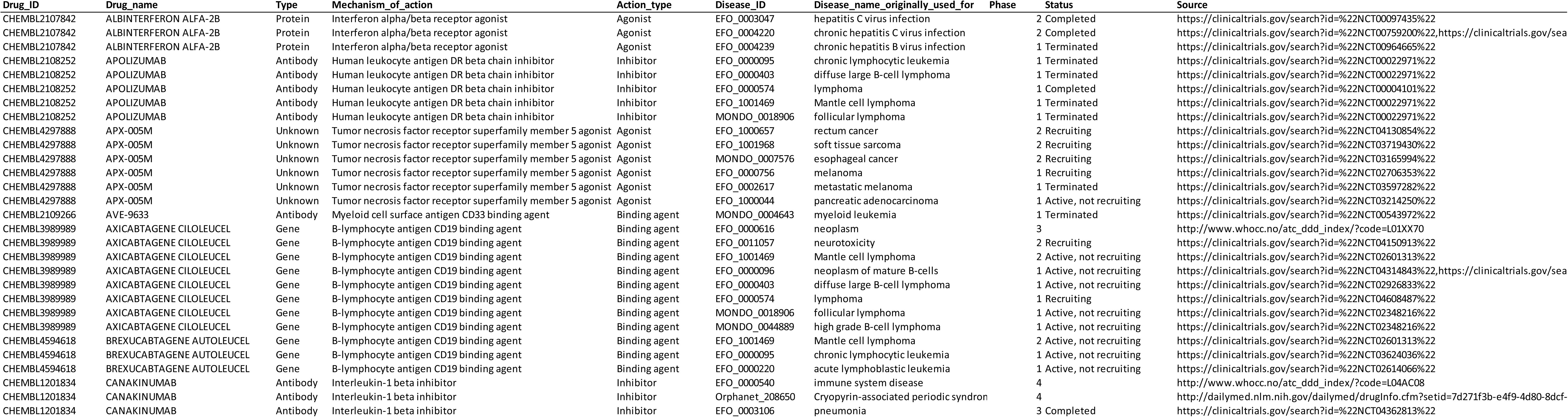
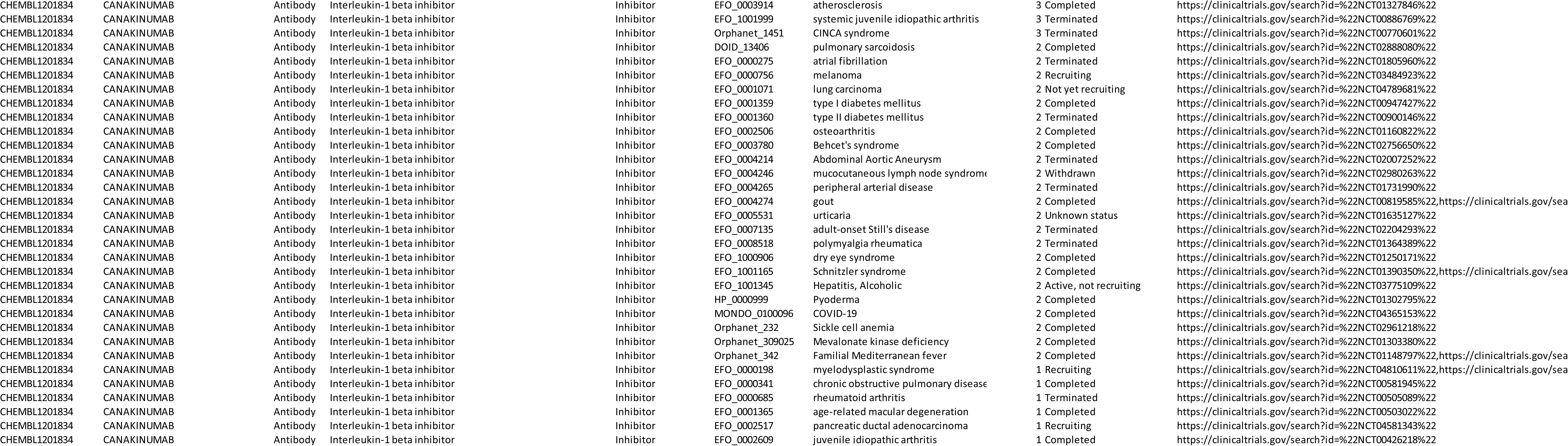
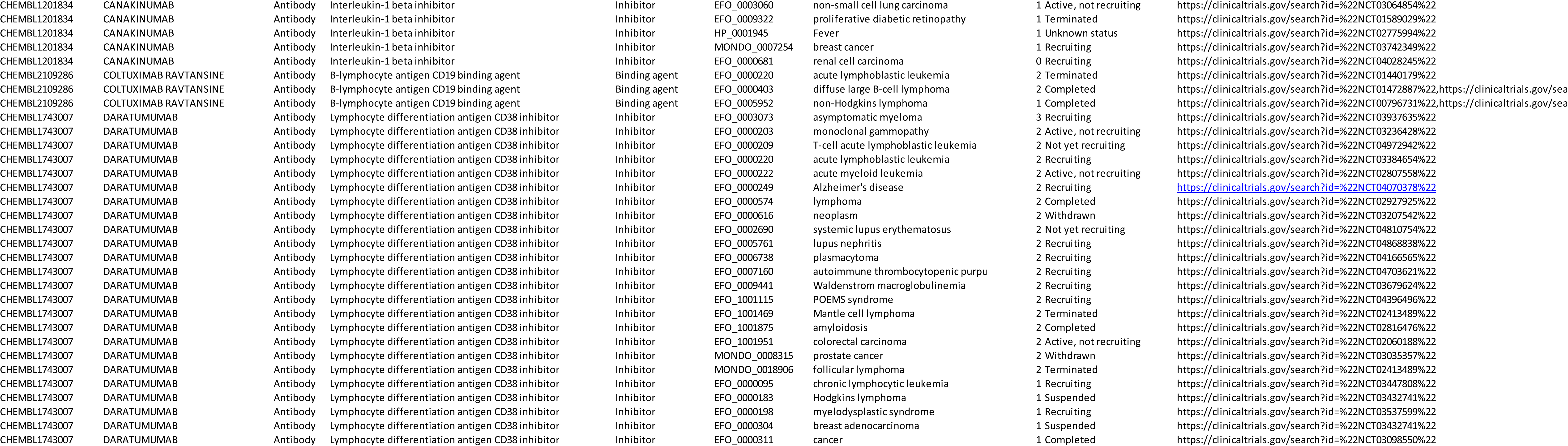
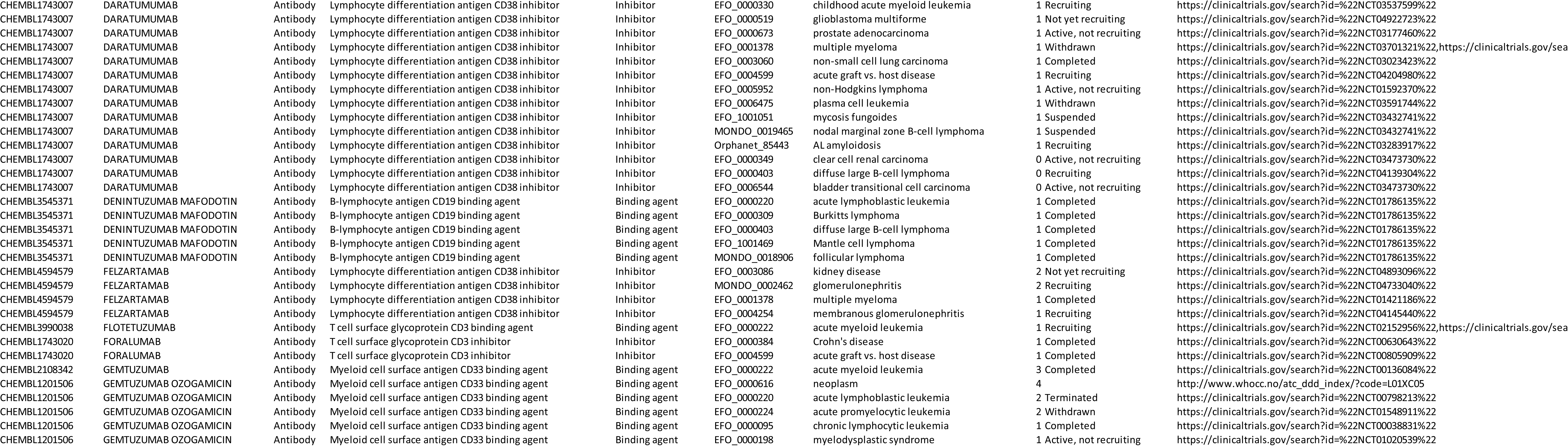
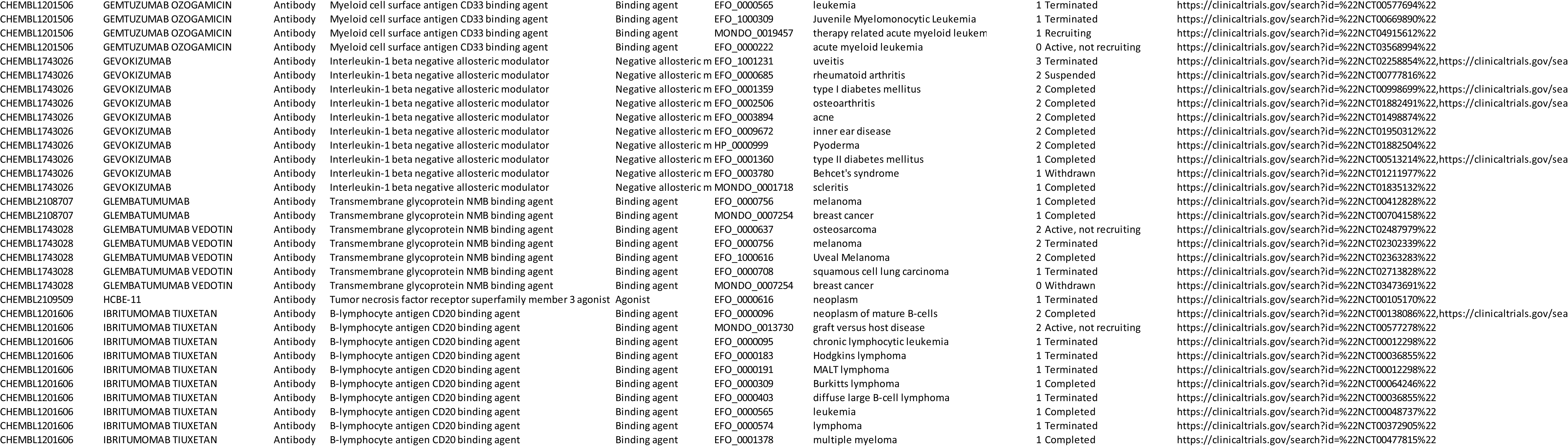
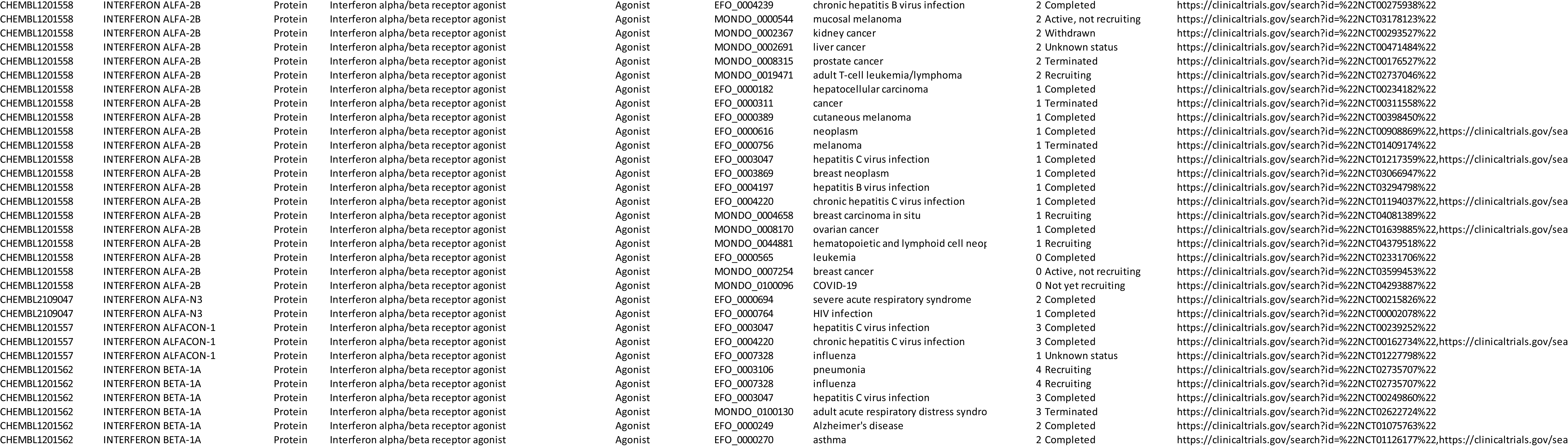
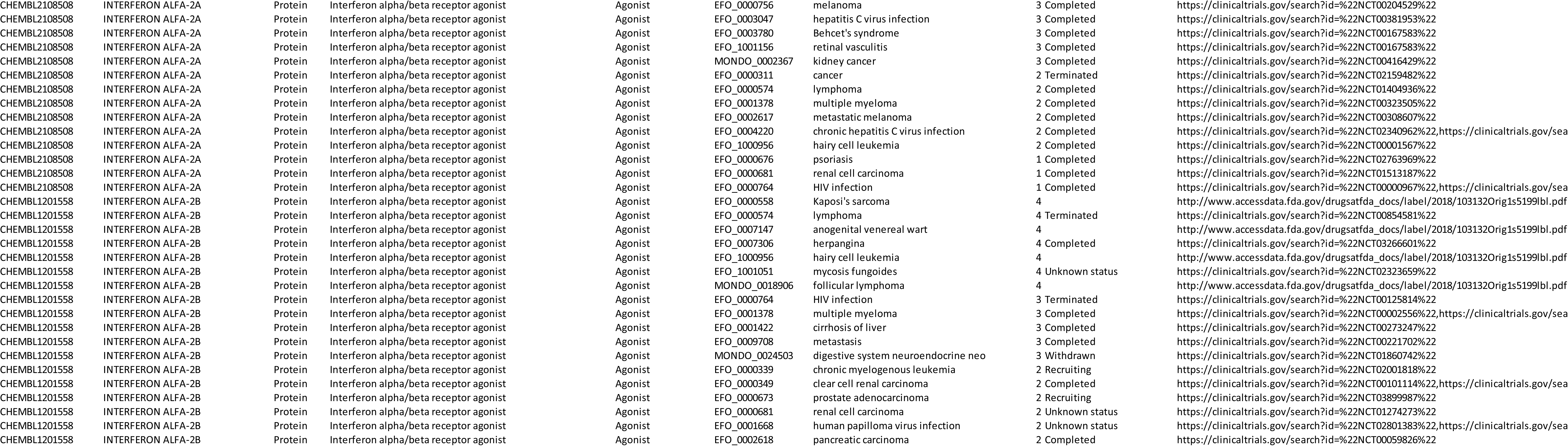
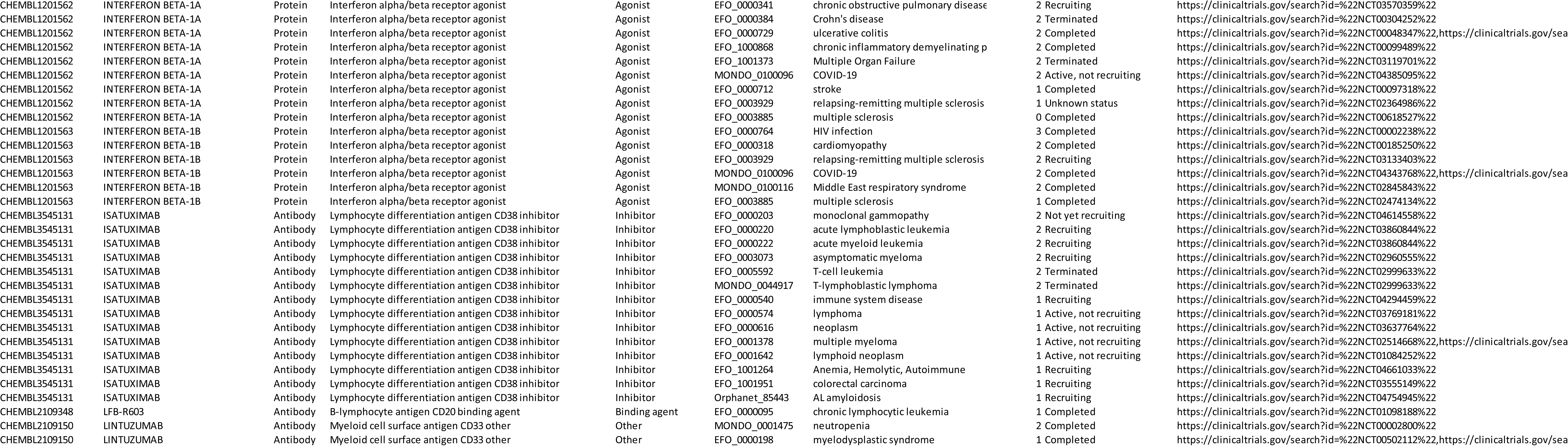
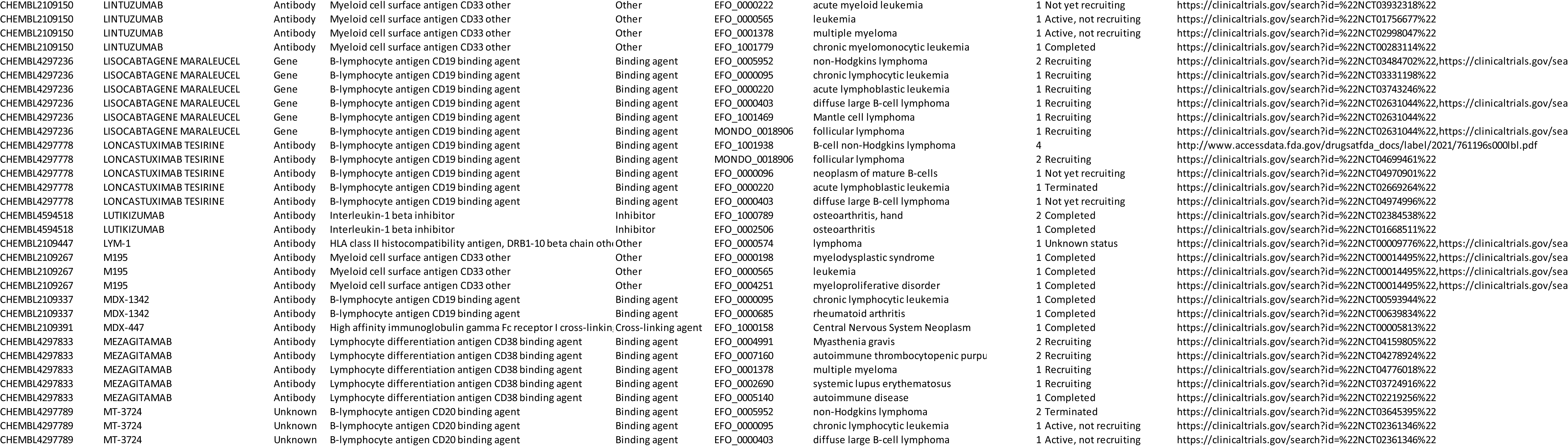
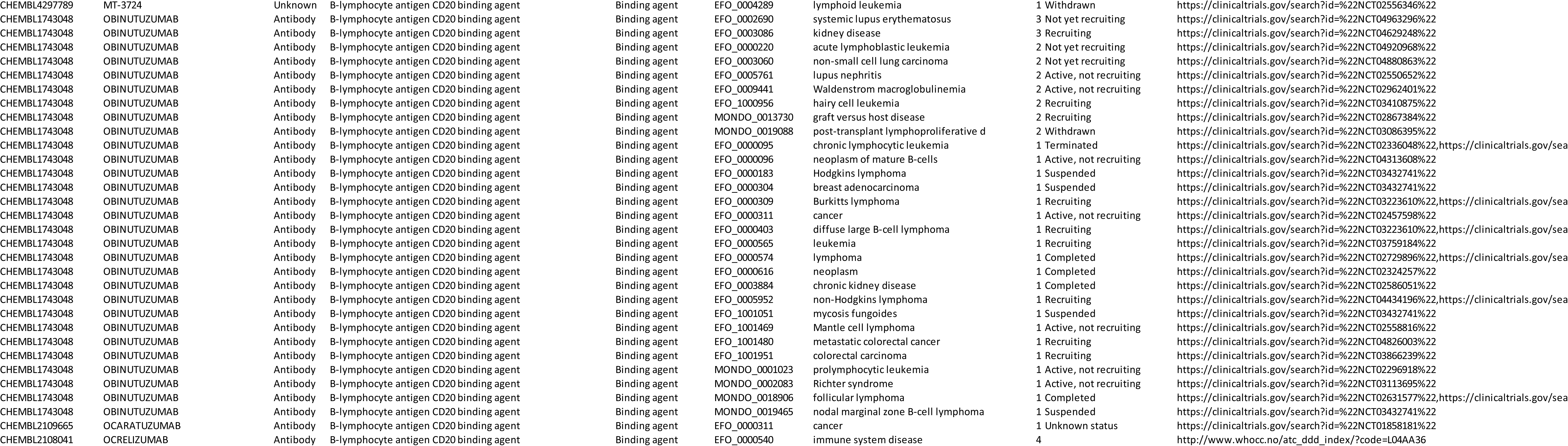
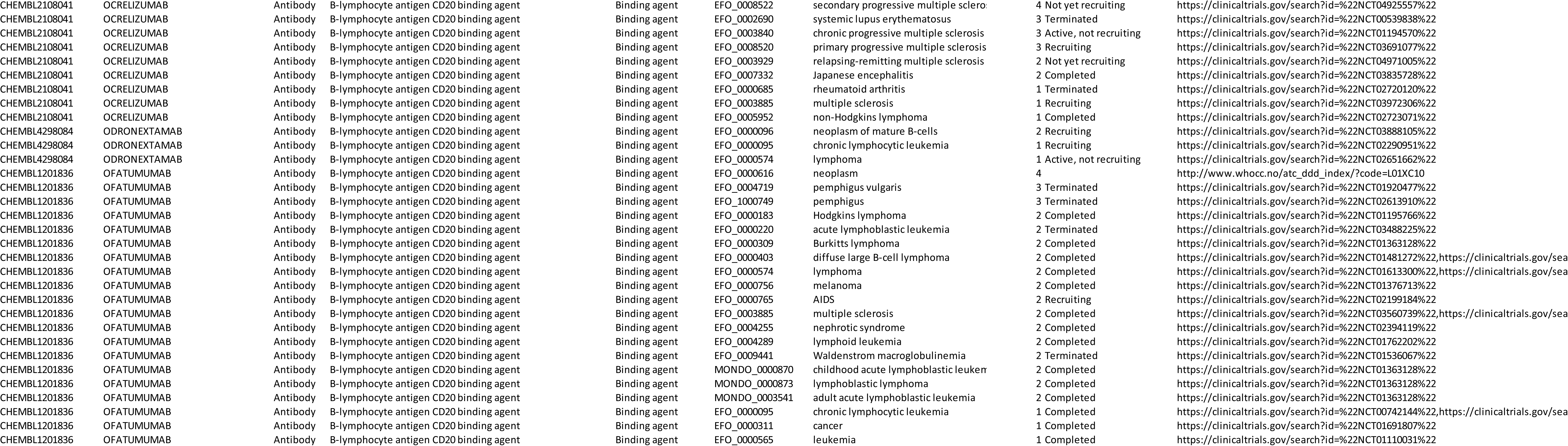
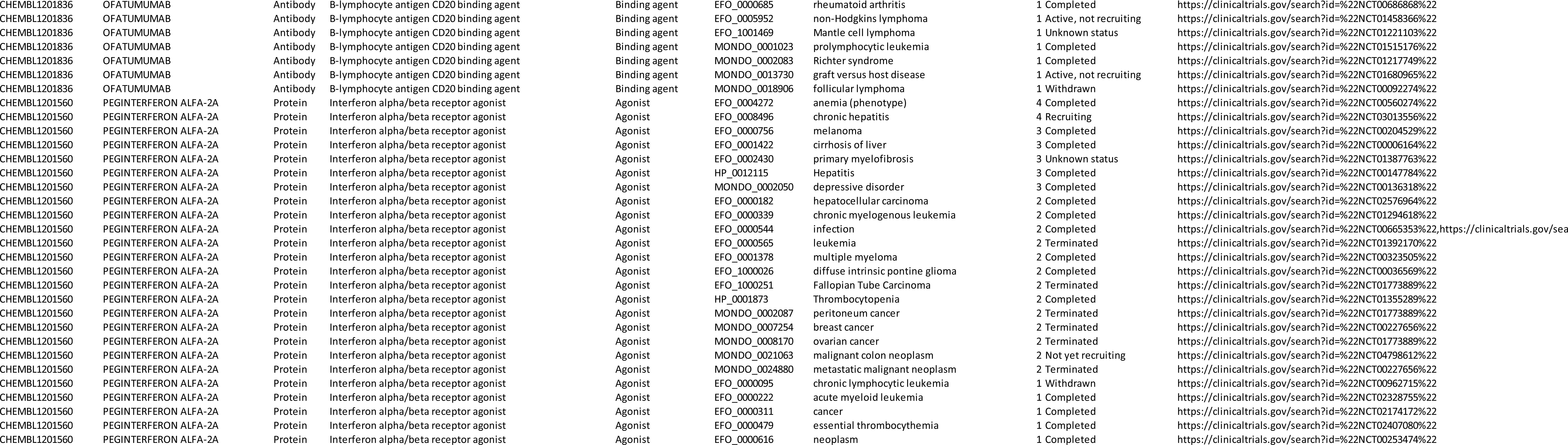
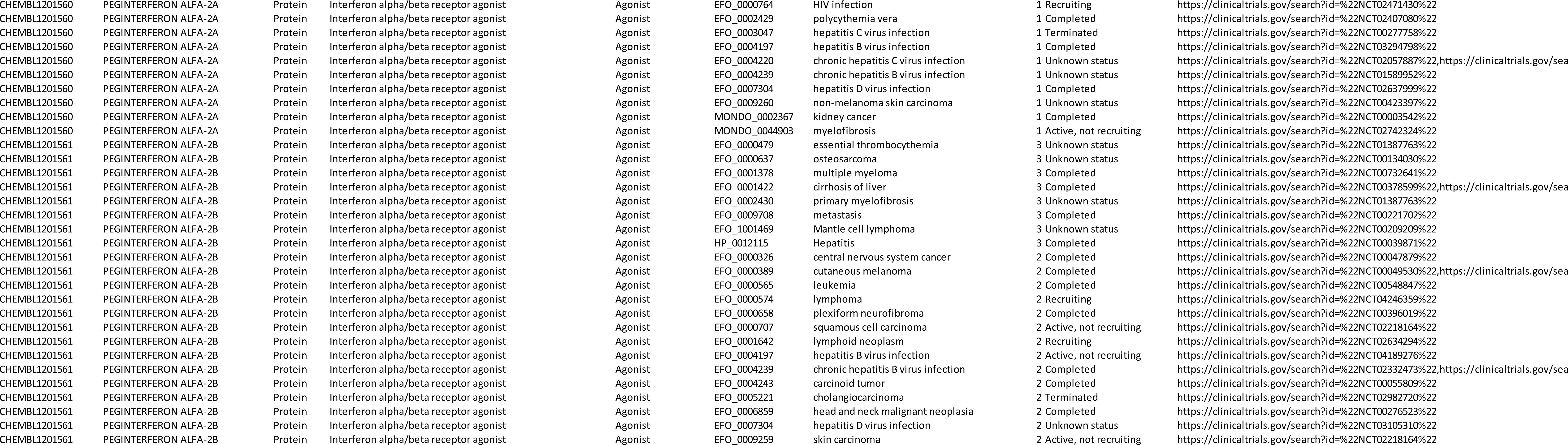
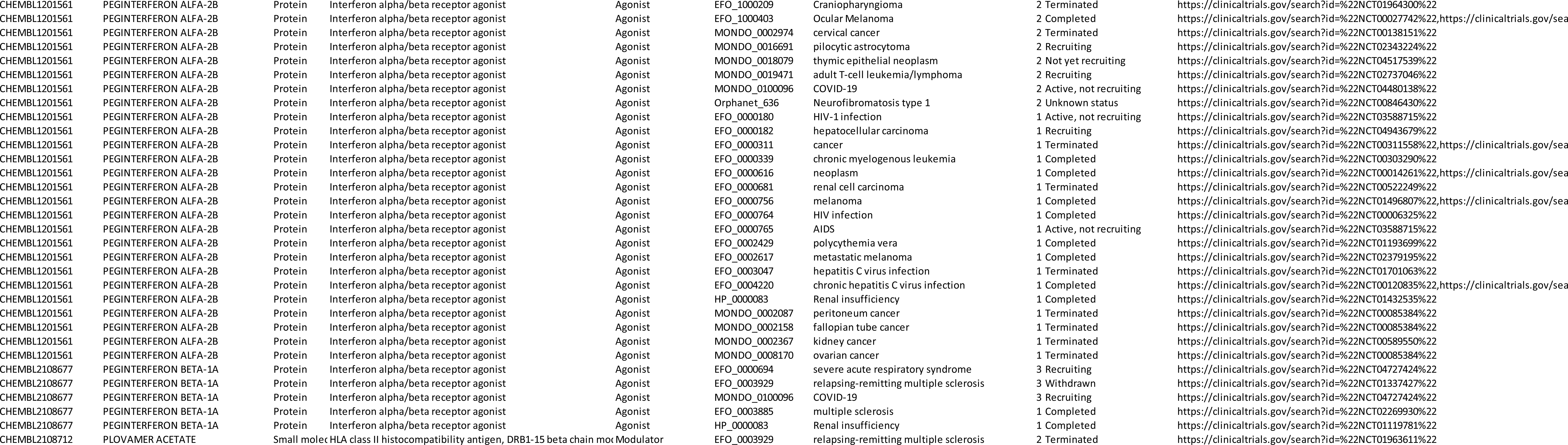
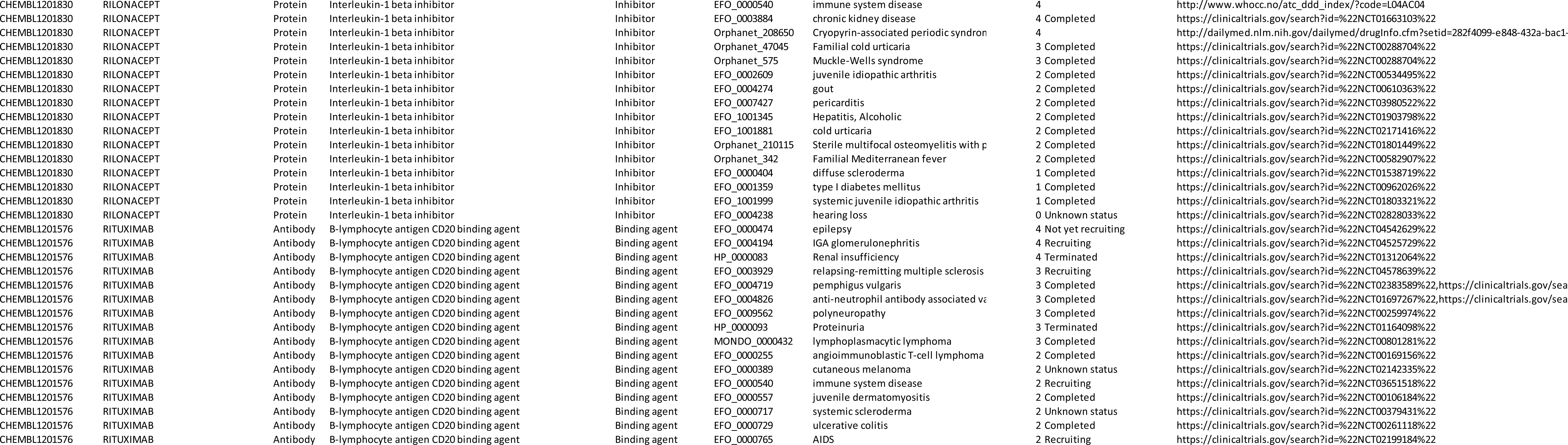
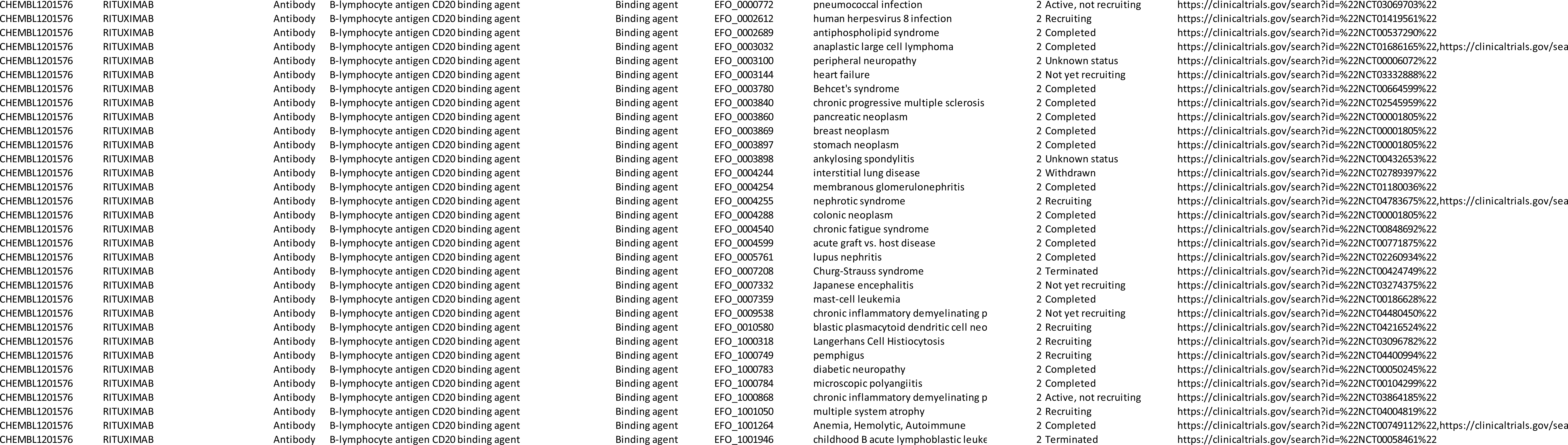
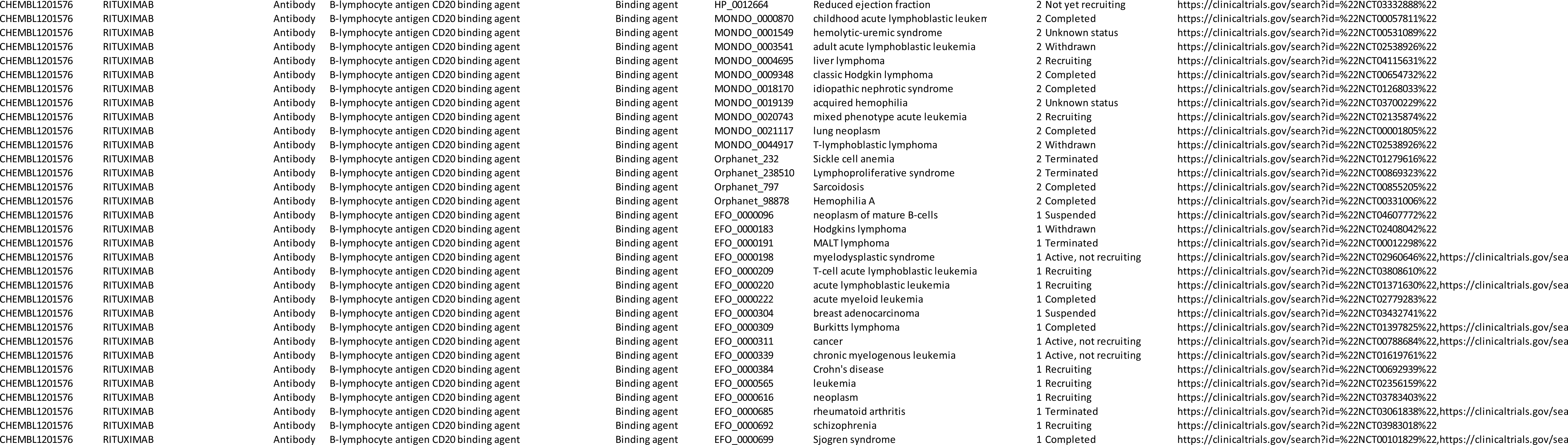
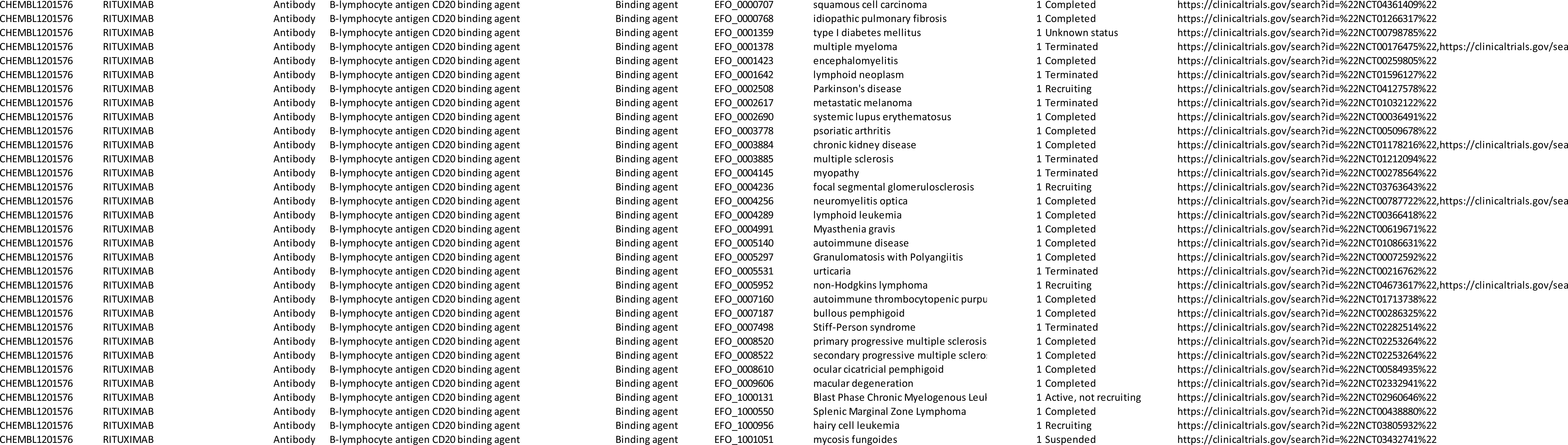
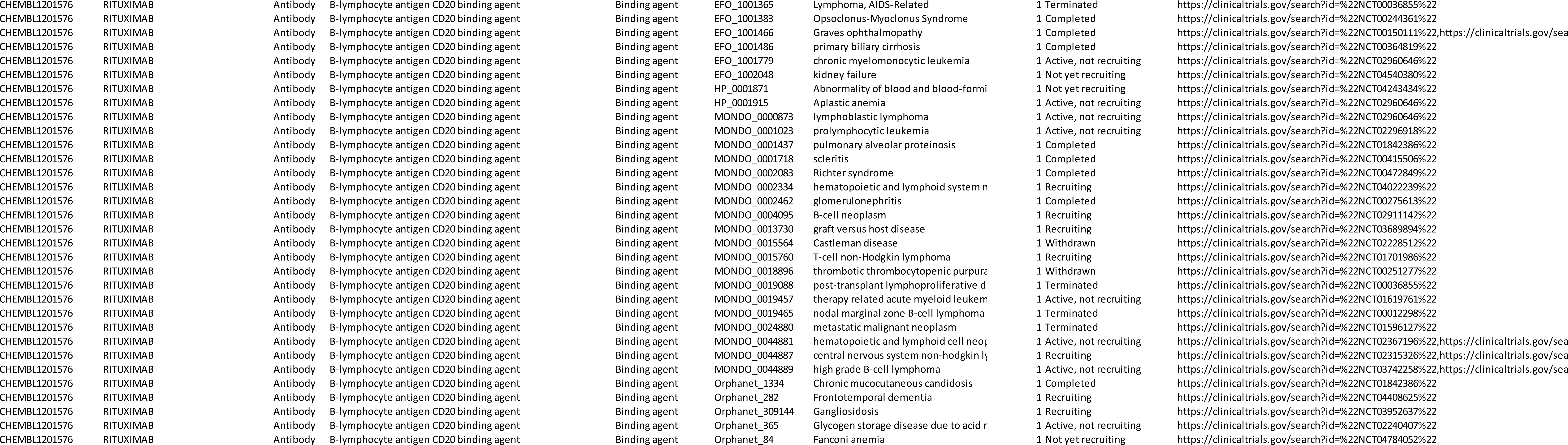
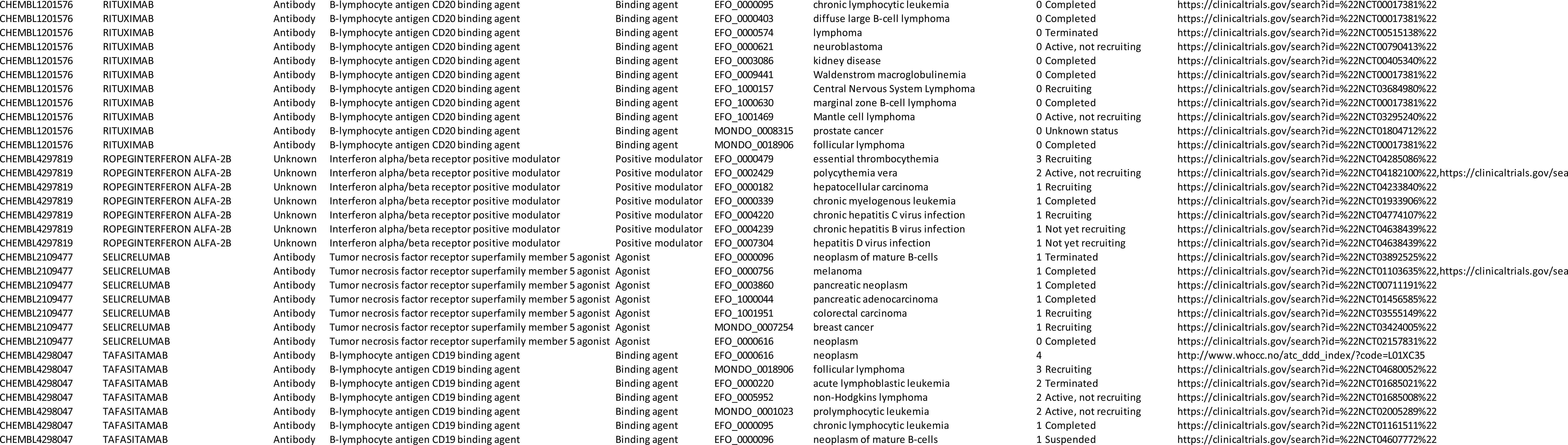
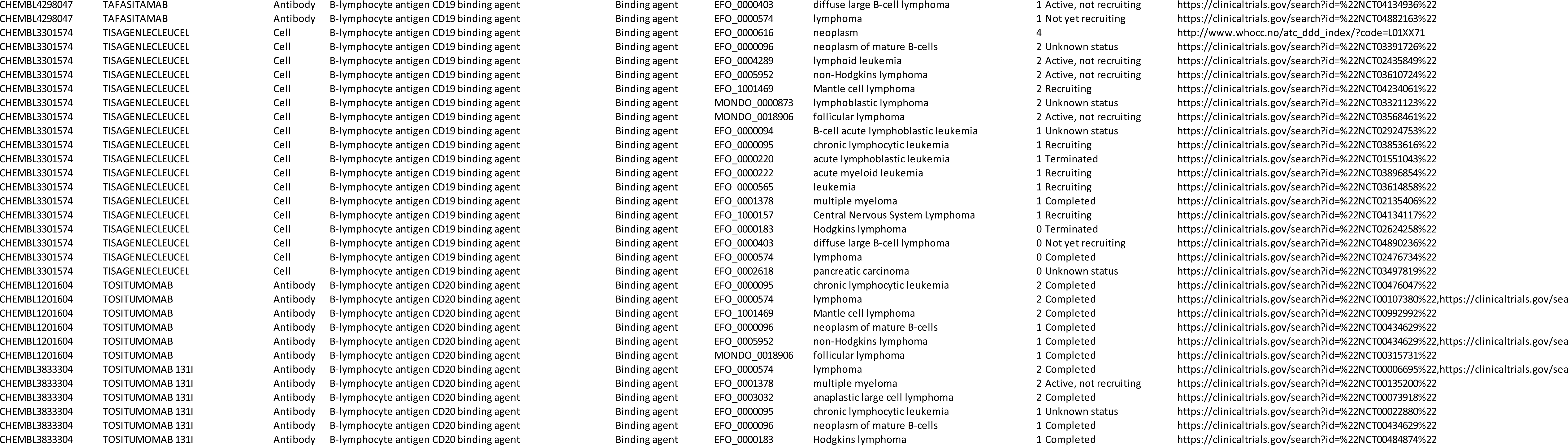
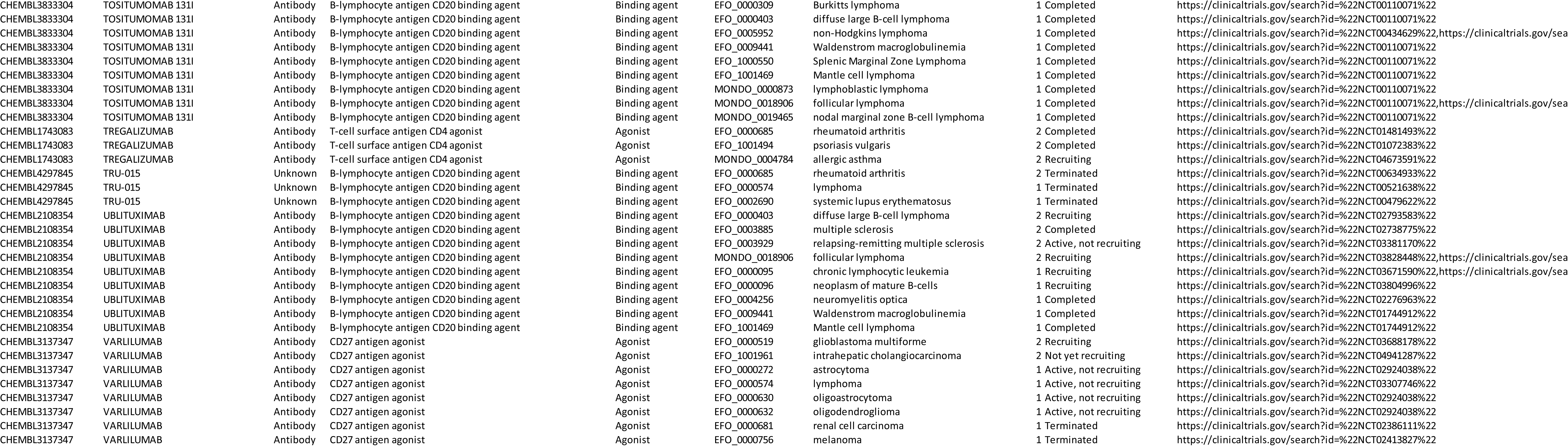
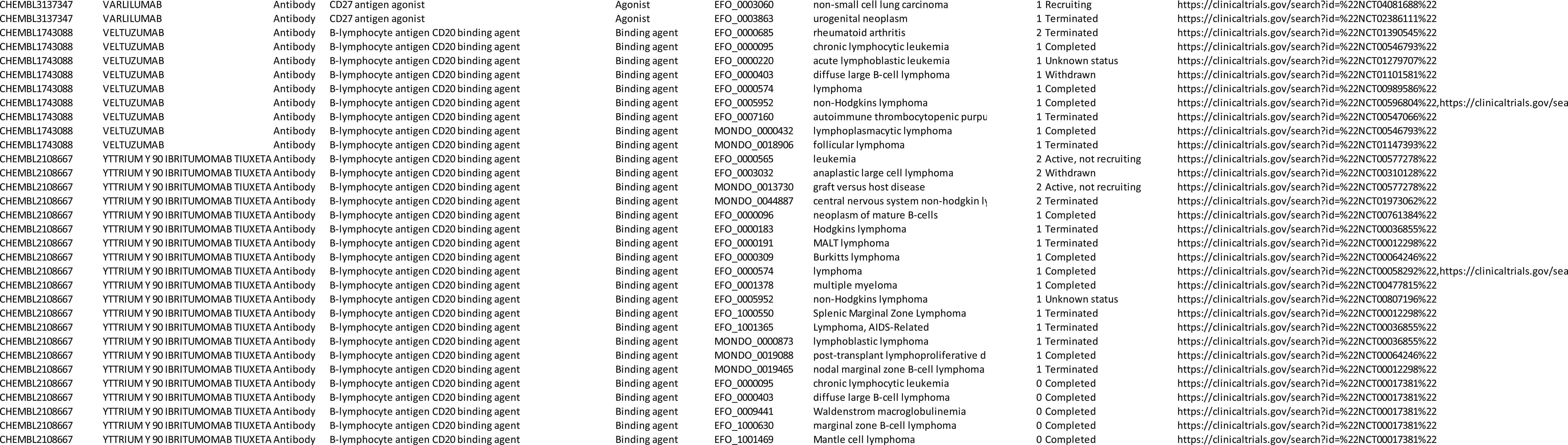
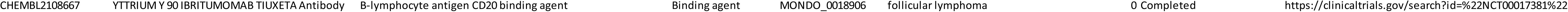
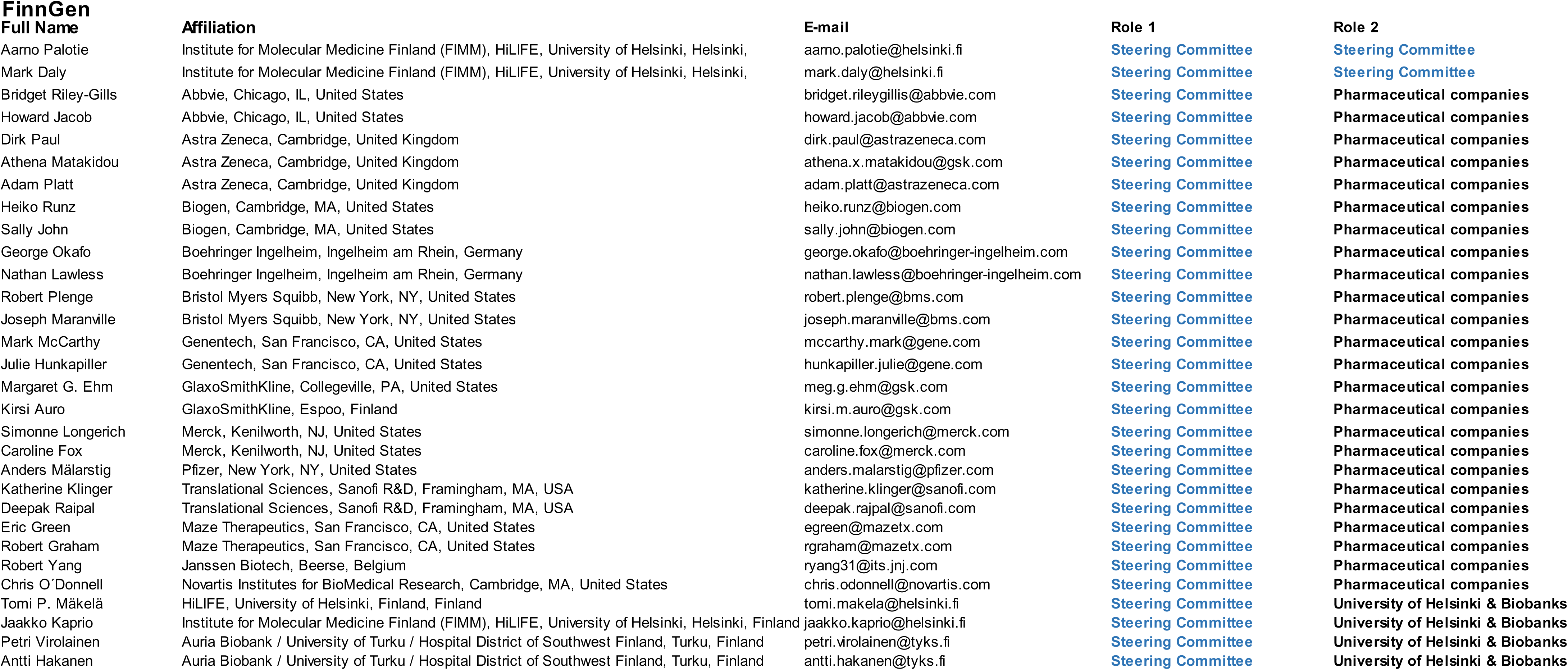
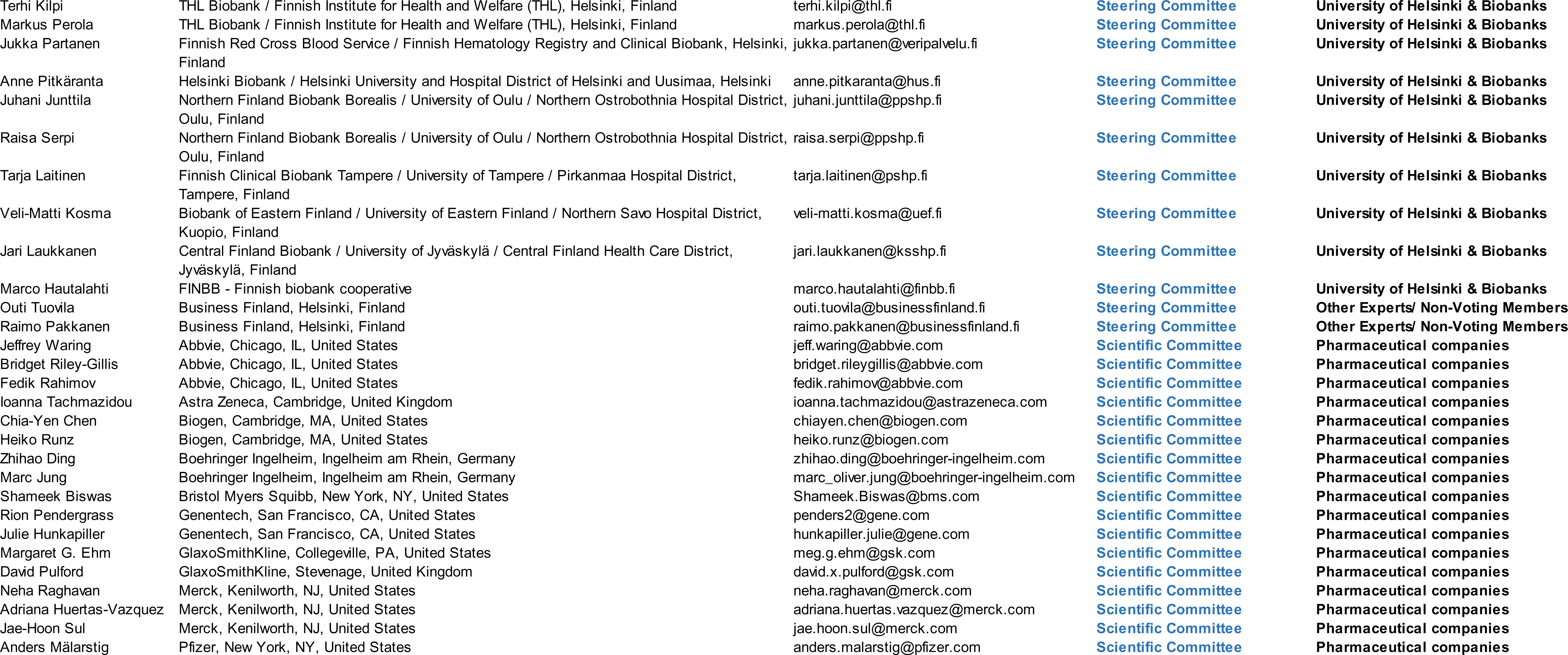
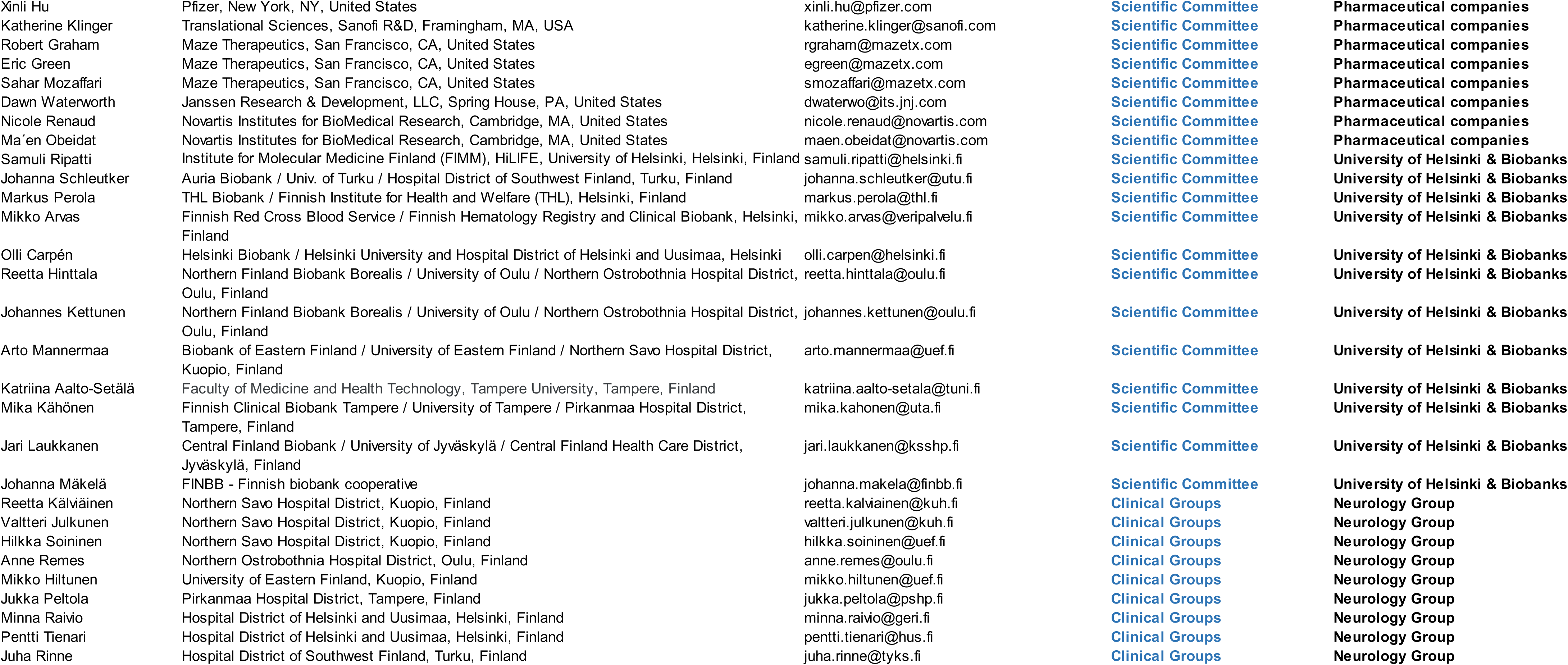
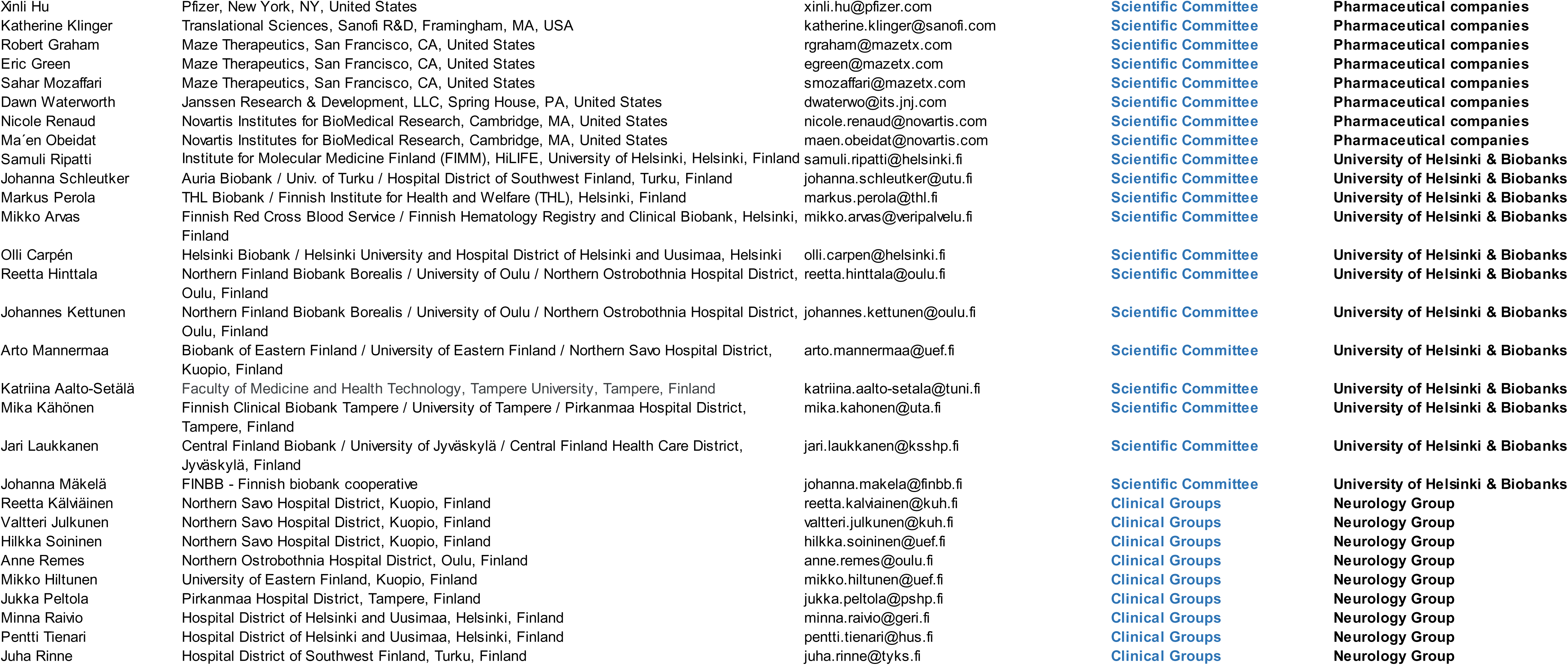
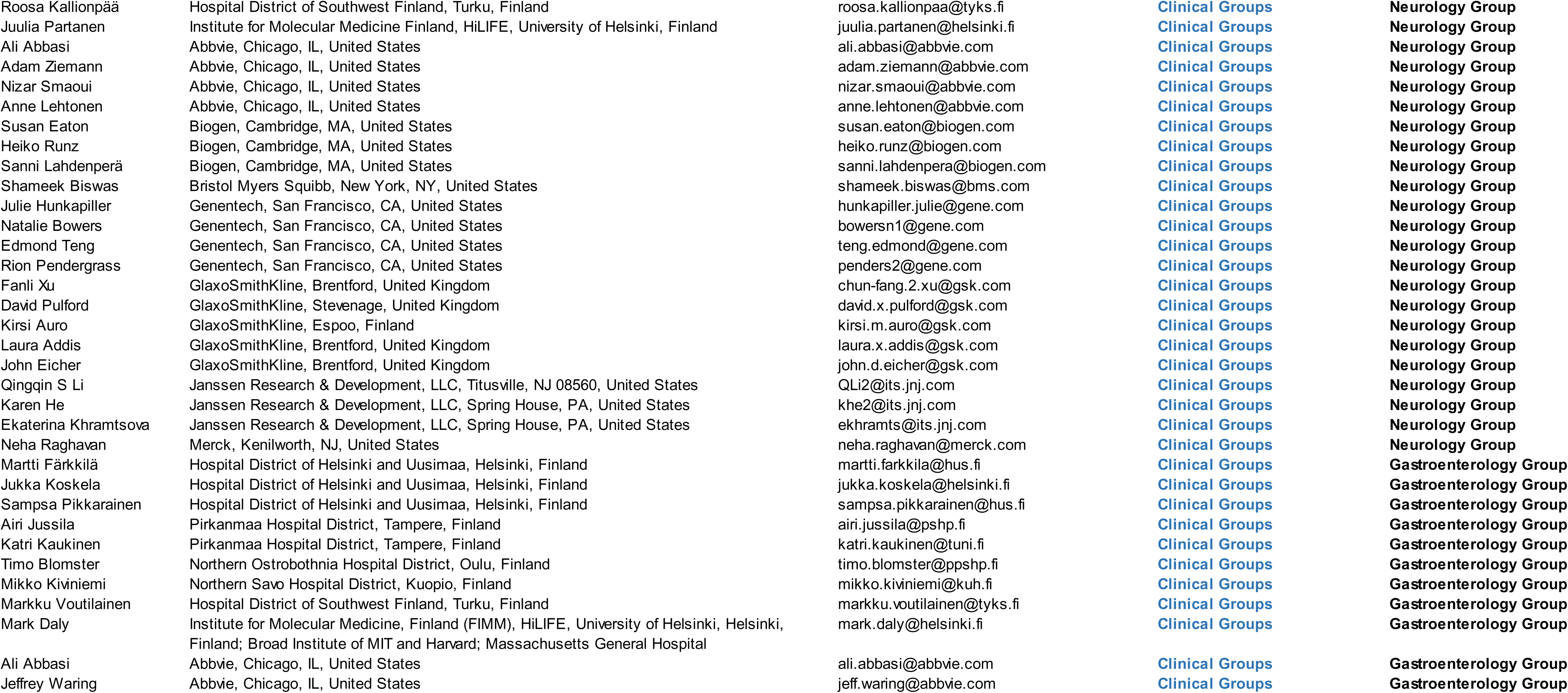
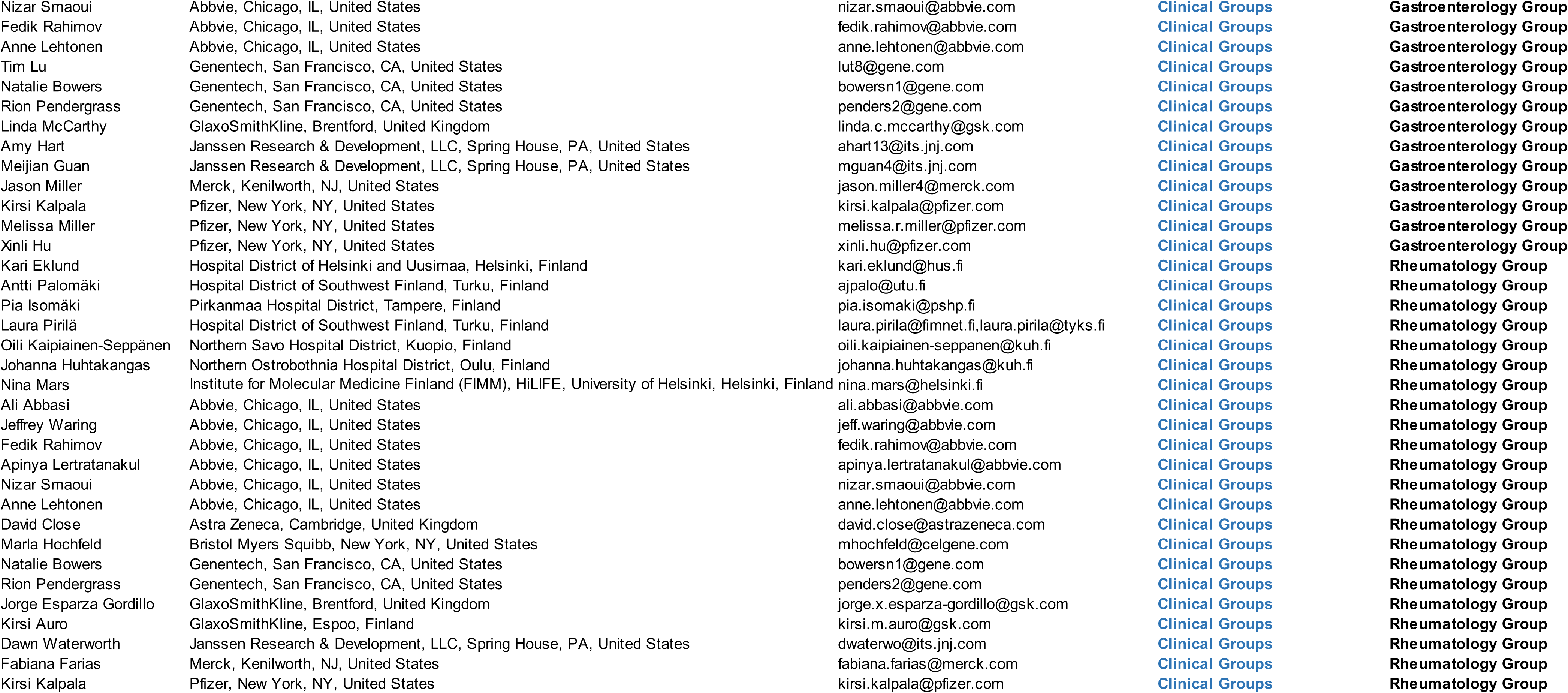
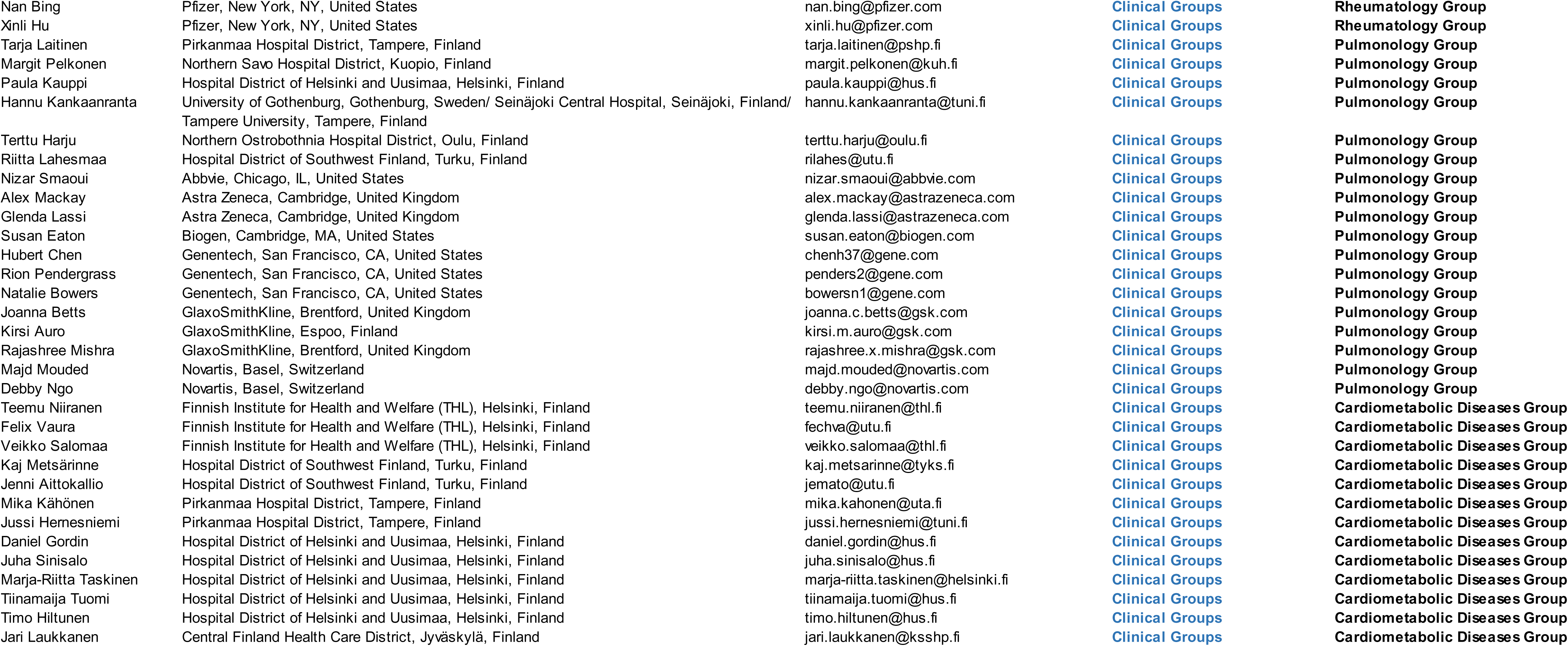
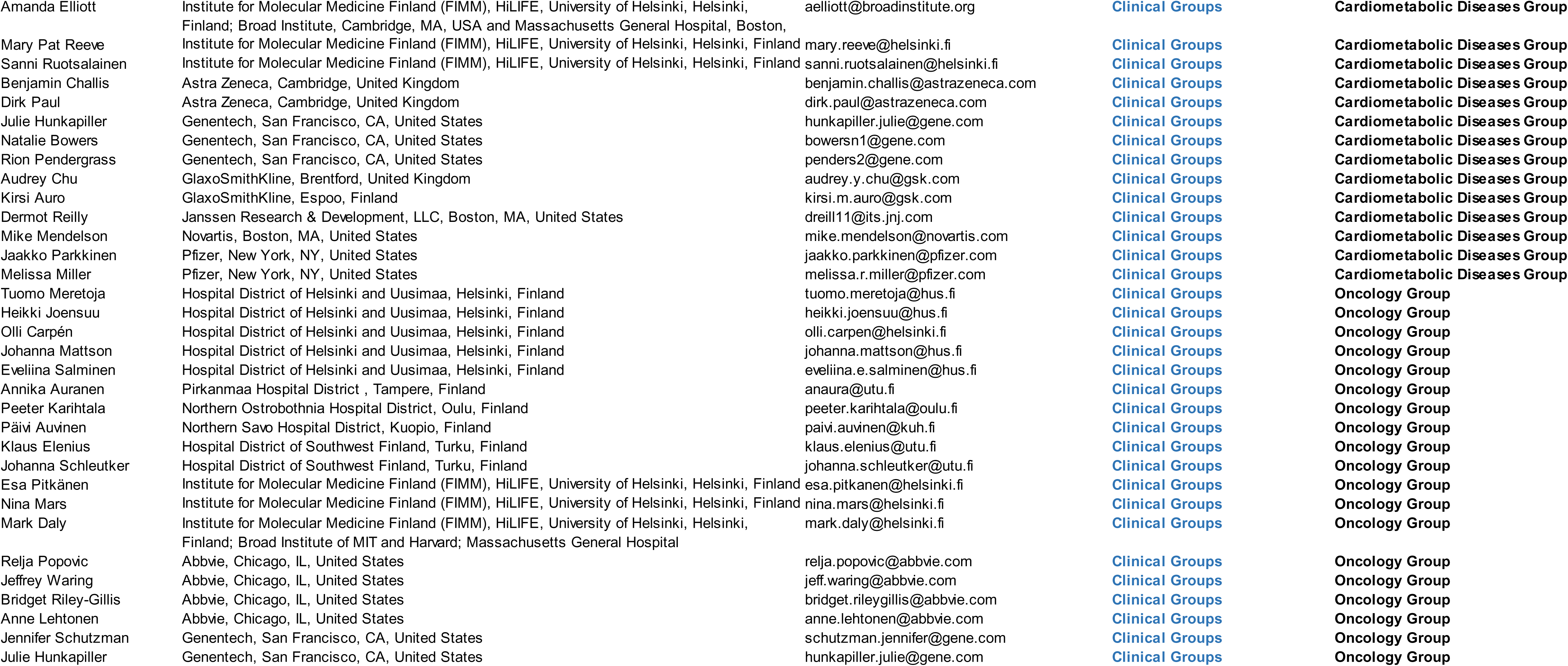
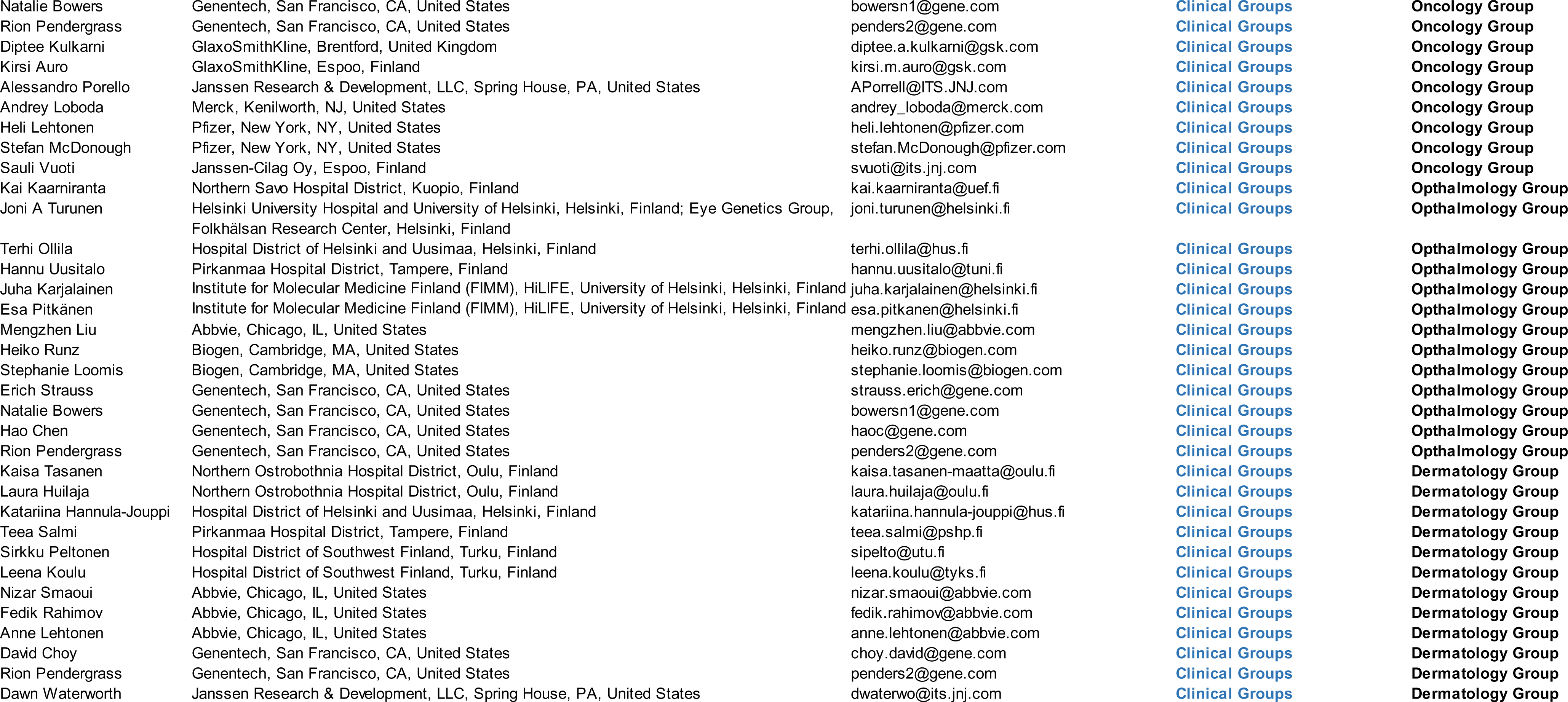
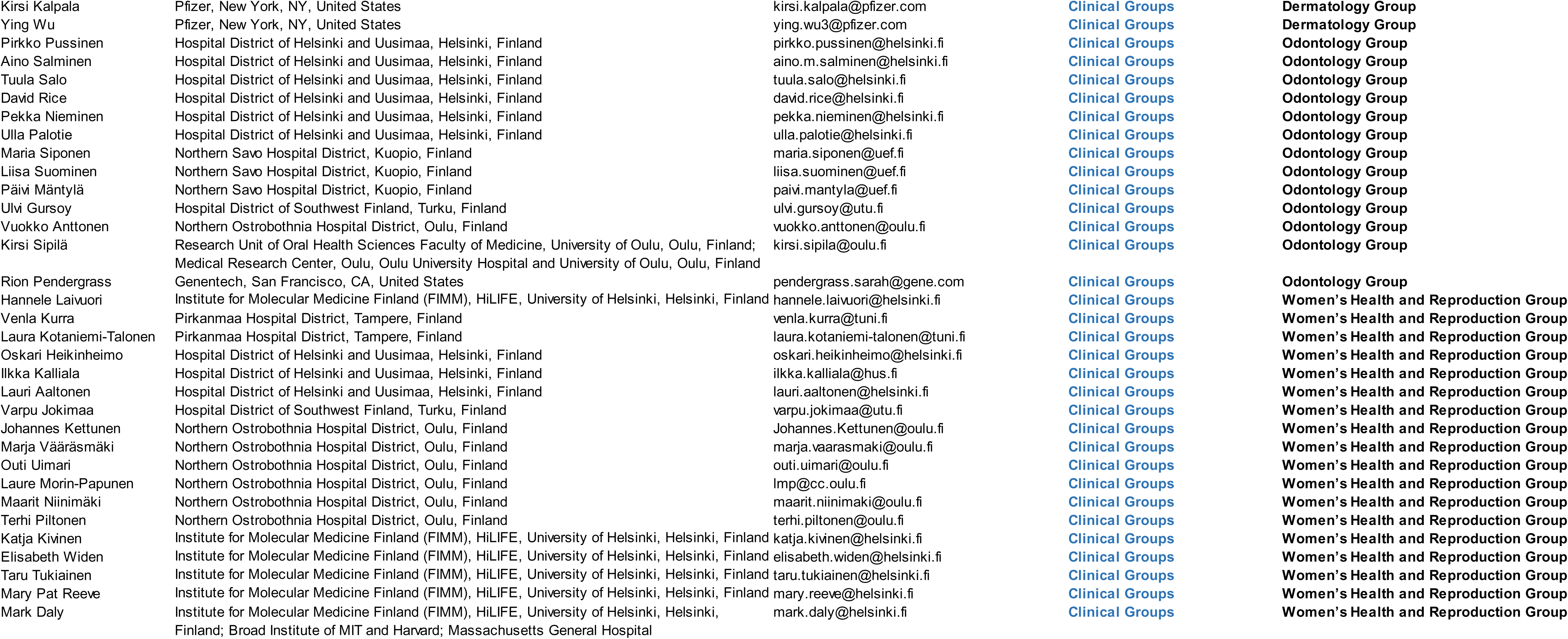
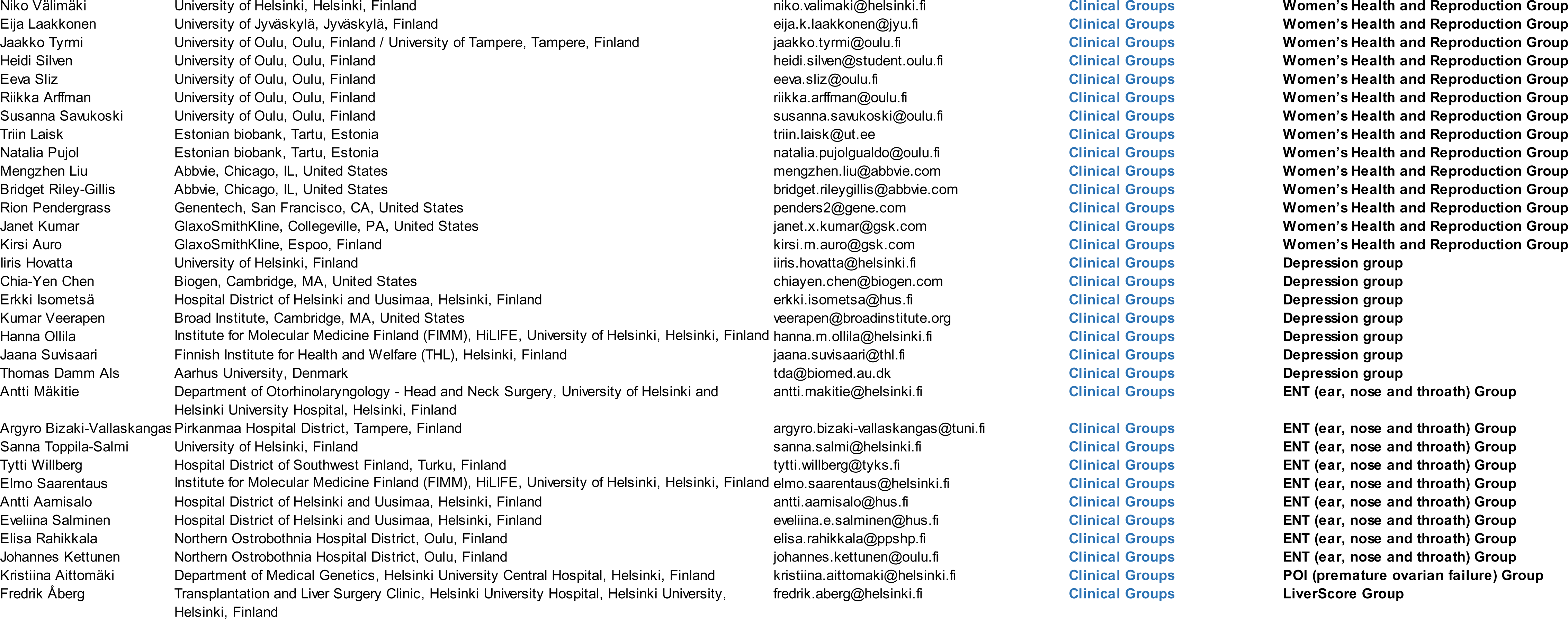
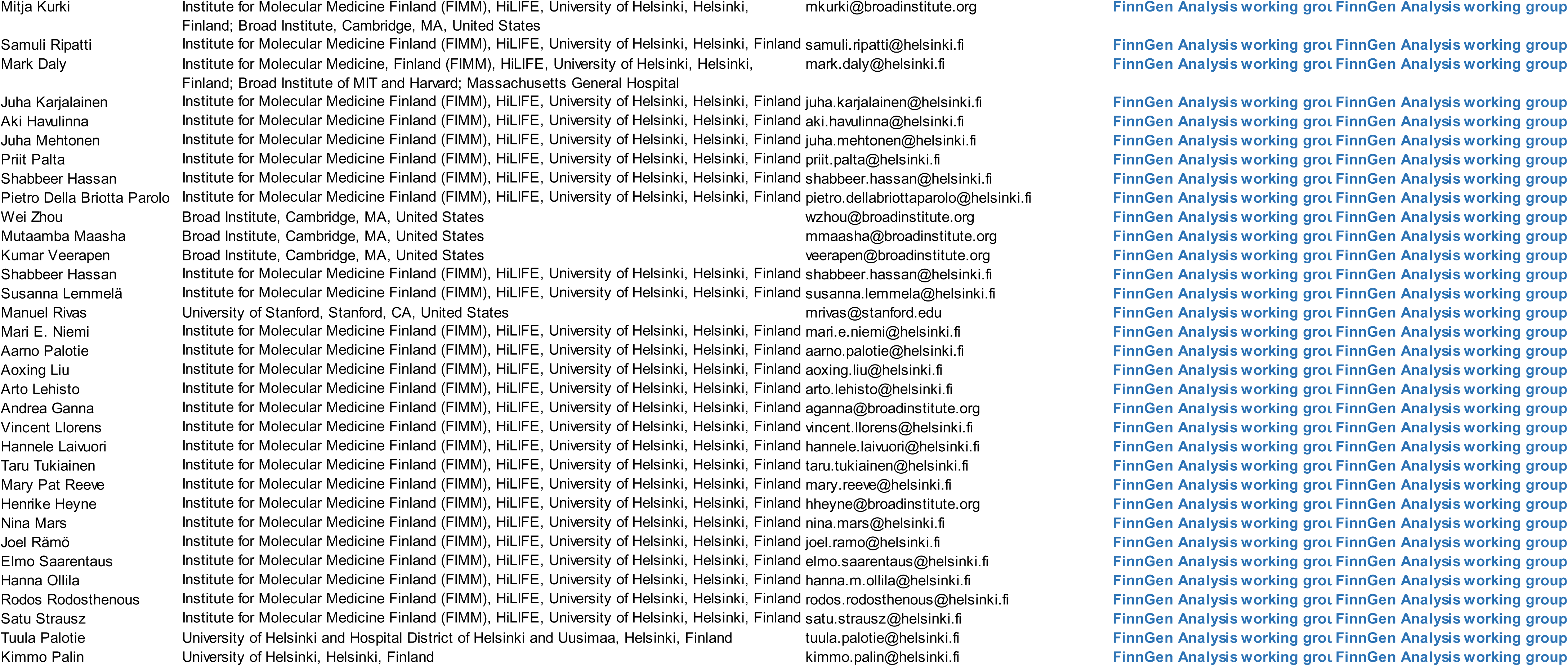
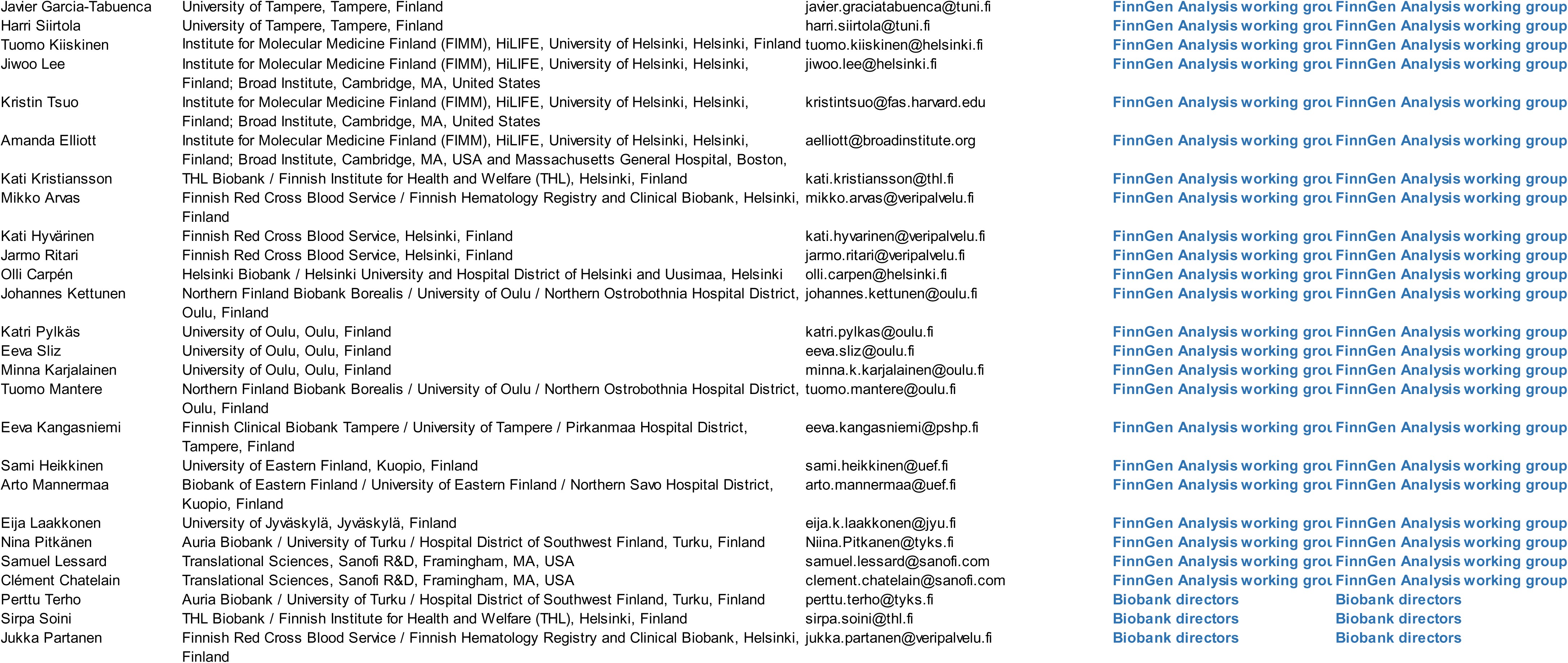
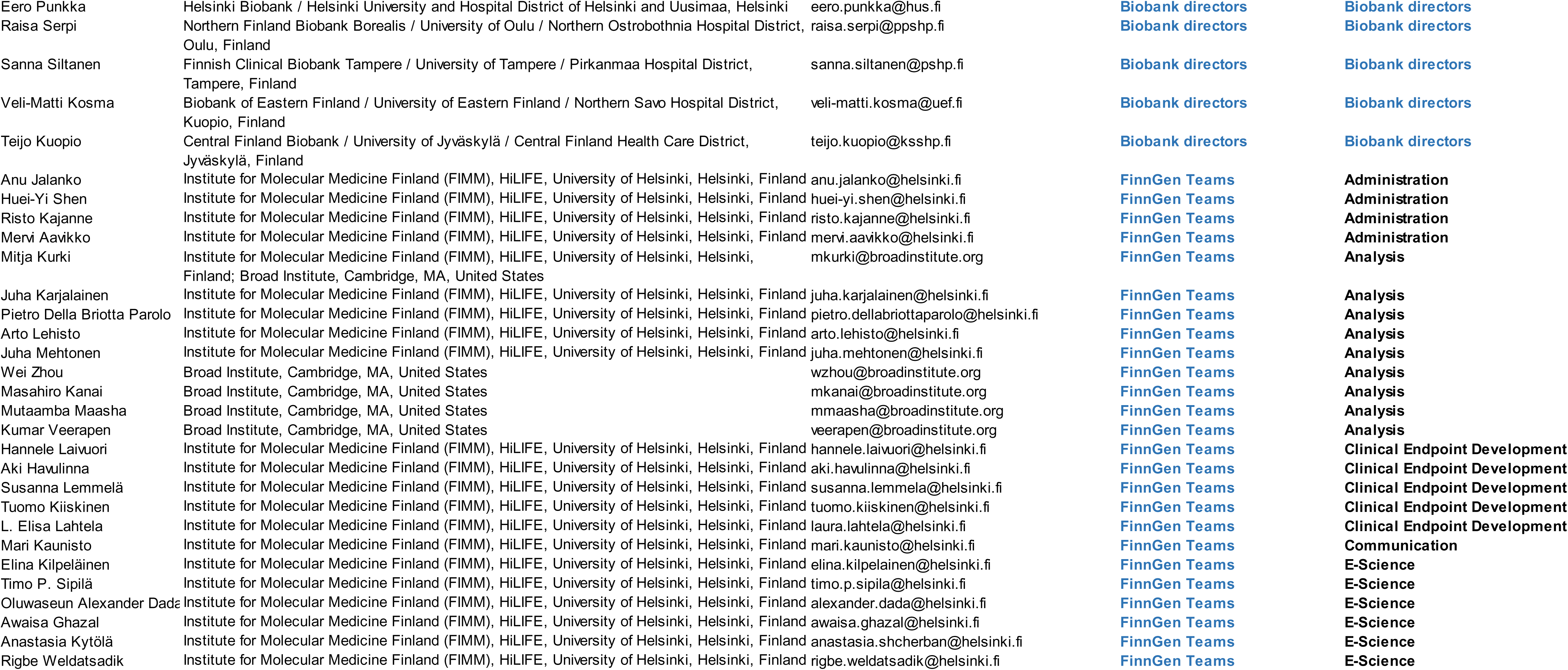
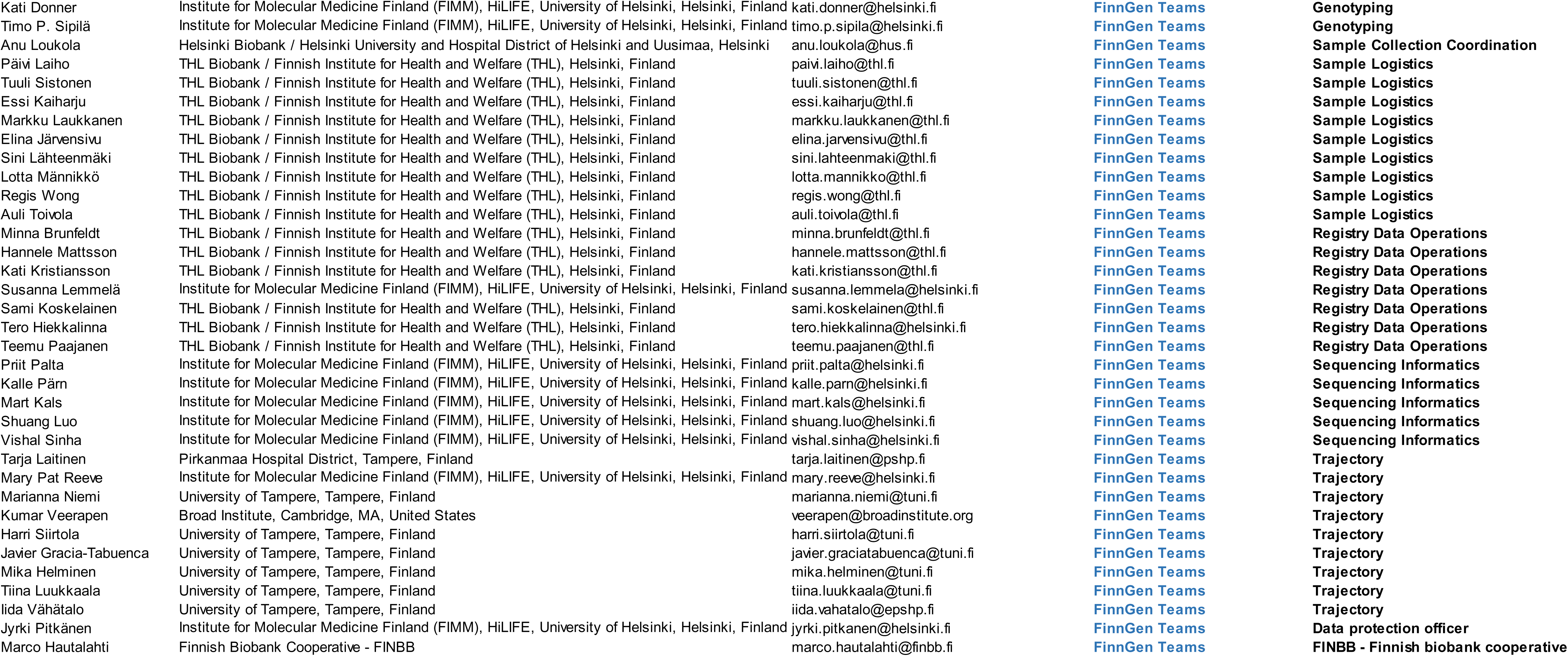
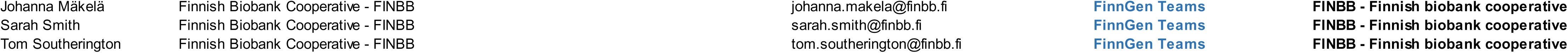
Drugs available in the Open Targets database that act on the biomarkers that associated with dementias and are hypothized to reduce the risk. In total 64 different drugs were identified.

## Notes

### Author Declarations

In the Whitehall II study, research ethics approvals were renewed at each wave; the most recent approval was obtained from the University College London Hospital Committee on the Ethics of Human Research (reference number 85/0938). Written, informed consent from participants was obtained at each contact. Patients and control subjects in FinnGen provided informed consent for biobank research, based on the Finnish Biobank Act. Alternatively, separate research cohorts, collected prior the Finnish Biobank Act came into effect (in September 2013) and start of FinnGen (August 2017), were collected based on study-specific consents and later transferred to the Finnish biobanks after approval by Fimea (Finnish Medicines Agency), the National Supervisory Authority for Welfare and Health. Recruitment protocols followed the biobank protocols approved by Fimea. The Coordinating Ethics Committee of the Hospital District of Helsinki and Uusimaa (HUS) statement number for the FinnGen study is Nr HUS/990/2017. The FinnGen study is approved by Finnish Institute for Health and Welfare (permit numbers: THL/2031/6.02.00/2017, THL/1101/5.05.00/2017, THL/341/6.02.00/2018, THL/2222/6.02.00/2018, THL/283/6.02.00/2019, THL/1721/5.05.00/2019 and THL/1524/5.05.00/2020), Digital and population data service agency (permit numbers: VRK43431/2017-3, VRK/6909/2018-3, VRK/4415/2019-3), the Social Insurance Institution (permit numbers: KELA 58/522/2017, KELA 131/522/2018, KELA 70/522/2019, KELA 98/522/2019, KELA 134/522/2019, KELA 138/522/2019, KELA 2/522/2020, KELA 16/522/2020), Findata permit numbers THL/2364/14.02/2020, THL/4055/14.06.00/2020,,THL/3433/14.06.00/2020, THL/4432/14.06/2020, THL/5189/14.06/2020, THL/5894/14.06.00/2020, THL/6619/14.06.00/2020, THL/209/14.06.00/2021, THL/688/14.06.00/2021, THL/1284/14.06.00/2021, THL/1965/14.06.00/2021, THL/5546/14.02.00/2020, THL/2658/14.06.00/2021, THL/4235/14.06.00/2021 and Statistics Finland (permit numbers: TK-53-1041-17 and TK/143/07.03.00/2020 (earlier TK-53-90-20) TK/1735/07.03.00/2021). The Biobank Access Decisions for FinnGen samples and data utilized in FinnGen Data Freeze 8 include: THL Biobank BB2017_55, BB2017_111, BB2018_19, BB_2018_34, BB_2018_67, BB2018_71, BB2019_7, BB2019_8, BB2019_26, BB2020_1, Finnish Red Cross Blood Service Biobank 7.12.2017, Helsinki Biobank HUS/359/2017, Auria Biobank AB17-5154 and amendment #1 (August 17 2020), AB20-5926 and amendment #1 (April 23 2020), Biobank Borealis of Northern Finland_2017_1013, Biobank of Eastern Finland 1186/2018 and amendment 22 /2020, Finnish Clinical Biobank Tampere MH0004 and amendments (21.02.2020 & 06.10.2020), Central Finland Biobank 1-2017, and Terveystalo Biobank STB 2018001.

